# Antidepressant Exposure and DNA Methylation: Insights from a Methylome-Wide Association Study

**DOI:** 10.1101/2024.05.01.24306640

**Authors:** E Davyson, X Shen, F Huider, M Adams, K Borges, D McCartney, L Barker, J Van Dongen, D Boomsma, A Weihs, H Grabe, L Kühn, A Teumer, H Völzke, T Zhu, J Kaprio, M Ollikainen, FS David, S Meinert, F Stein, AJ Forstner, U Dannlowski, T Kircher, A Tapuc, D Czamara, EB Binder, T Brückl, A Kwong, P Yousefi, CCY Wong, L Arseneault, HL Fisher, J Mill, S Cox, P Redmond, TC Russ, E van den Oord, KA Aberg, B Penninx, RE Marioni, NR Wray, AM McIntosh

## Abstract

**Importance:** Understanding antidepressant mechanisms could help design more effective and tolerated treatments.

**Objective:** Identify DNA methylation (DNAm) changes associated with antidepressant exposure.

**Design:** Case-control methylome-wide association studies (MWAS) of antidepressant exposure were performed from blood samples collected between 2006-2011 in Generation Scotland (GS). The summary statistics were tested for enrichment in specific tissues, gene ontologies and an independent MWAS in the Netherlands Study of Depression and Anxiety (NESDA). A methylation profile score (MPS) was derived and tested for its association with antidepressant exposure in eight independent cohorts, alongside prospective data from GS.

**Setting:** Cohorts; GS, NESDA, FTC, SHIP-Trend, FOR2107, LBC1936, MARS-UniDep, ALSPAC, E-Risk, and NTR.

**Participants:** Participants with DNAm data and self-report/prescription derived antidepressant exposure.

**Main Outcome(s) and Measure(s):** Whole-blood DNAm levels were assayed by the EPIC/450K Illumina array (9 studies, N_exposed_ = 661, N_unexposed_= 9,575) alongside MBD-Seq in NESDA (N_exposed_= 398, N_unexposed_= 414). Antidepressant exposure was measured by self-report and/or antidepressant prescriptions.

**Results:** The self-report MWAS (N = 16,536, N_exposed_ = 1,508, mean age = 48, 59% female) and the prescription-derived MWAS (N = 7,951, N_exposed_ = 861, mean age = 47, 59% female), found hypermethylation at seven and four DNAm sites (p < 9.42x10^-8^), respectively. The top locus was cg26277237 (*KANK1,* p_self-report_*=* 9.3x10^-13^, p_prescription_ = 6.1x10^-3^). The self-report MWAS found a differentially methylated region, mapping to *DGUOK-AS1 (*p_adj_ = 5.0x10^-3^) alongside significant enrichment for genes expressed in the amygdala, the “synaptic vesicle membrane” gene ontology and the top 1% of CpGs from the NESDA MWAS (OR = 1.39, p < 0.042). The MPS was associated with antidepressant exposure in meta-analysed data from external cohorts (N_studies_= 9, N = 10,236, N_exposed_ = 661, f3 = 0.196, p < 1x10^-4^).

**Conclusions and Relevance:** Antidepressant exposure is associated with changes in DNAm across different cohorts. Further investigation into these changes could inform on new targets for antidepressant treatments.

**3 Key Points:** *Question:* Is antidepressant exposure associated with differential whole blood DNA methylation?

*Findings:* In this methylome-wide association study of 16,536 adults across Scotland, antidepressant exposure was significantly associated with hypermethylation at CpGs mapping to *KANK1* and *DGUOK-AS1.* A methylation profile score trained on this sample was significantly associated with antidepressant exposure (pooled f3 [95%CI]=0.196 [0.105, 0.288], p < 1x10^-4^) in a meta-analysis of external datasets.

*Meaning:* Antidepressant exposure is associated with hypermethylation at *KANK1* and *DGUOK-AS1*, which have roles in mitochondrial metabolism and neurite outgrowth. If replicated in future studies, targeting these genes could inform the design of more effective and better tolerated treatments for depression.

## Introduction

Major Depressive Disorder (MDD) is predicted to become the leading cause of disability worldwide by 2030^1^, partly due to the limitations of current treatments^2^. Although antidepressants are commonly prescribed effective treatments^3^, they prove to be ineffective in a high proportion of cases, with an estimated 40% of those presenting with MDD developing treatment-resistant depression^4,5^. Furthermore, many treatments are commonly accompanied by side effects, including weight changes, fatigue and sexual dysfunction^2^. There is a need for more effective and better-tolerated antidepressant treatments and to target existing treatments to those most likely to respond. Advances are hampered by poor mechanistic understanding of both MDD itself^1,16,17^ and how currently prescribed antidepressants lead to therapeutic effects^9^.

The mechanism of currently prescribed antidepressants is incompletely understood. Initial theories surmised that their therapeutic effects were due to an increase in monoamine brain synaptic concentrations^10^. However, antidepressant treatment has a delayed onset for symptomatic improvement, which does not reflect the immediate effect on monoamine levels^7^. This casts doubt on the simple role of monoamines as a causal factor in MDD^7–9^, although other experimental paradigms continue to suggest their importance^11^. Another prominent theory of antidepressant action suggests that their therapeutic mechanism involves increasing synaptic remodelling^12^ and neuronal plasticity^9,13^. The evidence for the effect of antidepressants on DNA methylation (DNAm) is growing^14,15^. In vitro studies found that the antidepressant paroxetine interacted with DNA methyltransferase (DNMT), a key enzyme involved in DNAm^16^. Furthermore, studies of chronically stressed rodent models have found that stress-induced DNAm and behavioural changes are reversed through both antidepressant treatment^17^ and DNMT inhibitors^18^.

DNAm, the addition of a methyl group at a cytosine-phosphate-guanine (CpG) site, regulates gene expression and impacts cellular function^19,20^. In 2022, Barbu *et al.*^21^ performed a methylome-wide association study (MWAS) of self-reported antidepressant exposure in a subset of participants in Generation Scotland (GS, N = 6,428) and the Netherlands Twin Register (NTR, N = 2,449)^21^, identifying altered DNAm near to genes involved in the innate immune response in those exposed to antidepressants^21^. As self-report measures may be unreliable due to volunteer recall bias, a poor understanding of the medication nosology, and non-disclosure^22–24^, Barbu *et al.*^21^ also performed an MWAS of antidepressant exposure based on recorded antidepressant prescriptions in the last 12 months. However, this assumes continuous treatment, potentially overestimating exposure due to general low adherence to antidepressant medication^25^. Calculation of active treatment periods from consecutive prescribing events provides a potentially more reliable identification of antidepressant exposure^26^.

In our study, we build upon previous analyses by Barbu *et al.*^21^ by analysing a larger sample of GS (N = 16,536), and by estimating active treatment periods from prescribing records to identify those exposed to antidepressants at DNAm measurement. First, an MWAS was performed on both the self-report and prescription-derived measures of antidepressant exposure. Second, to assess the potential confounding by MDD, the MWAS analyses were restricted to MDD cases only. Third, functional follow-up analysis of differentially methylated CpG sites was performed. Fourth, we investigated the enrichment of top CpGs in GS and an independent MWAS conducted in the Netherlands Study of Depression and Anxiety (NESDA). Fifth, the relationship between time in treatment and DNAm at significant CpG sites was investigated. Finally, a methylation profile score (MPS) for self-report antidepressant exposure was trained in GS and tested for an association with antidepressant exposure in eight independent external datasets: Finn Twin Cohort (FTC), Study of Health in Pomerania (SHIP-Trend), Lothian Birth Cohort 1936 (LBC1936), FOR2107, NTR, Avon Longitudinal Study of Parents and Children (ALSPAC), Munich Antidepressant Response Study/Unipolar Depression Study (MARS-UniDep) and the Environmental Risk (E-Risk) Longitudinal Twin Study, alongside a prospective sample of GS: Stratifying Depression and Resilience Longitudinally (STRADL) (Figure 1).

**Figure 1:**
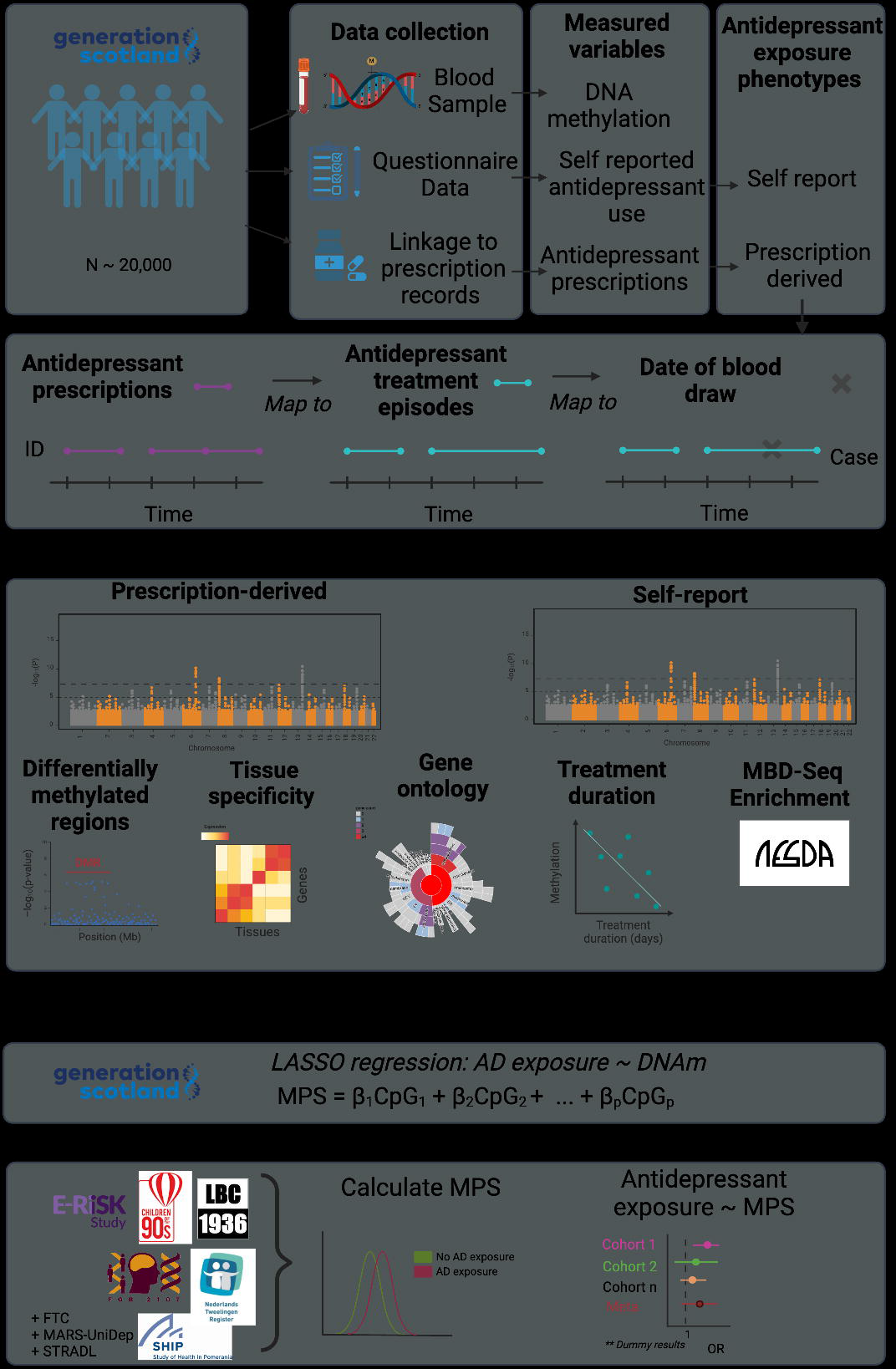
Schematic of study design.

## Methods and Materials

### Generation Scotland

Generation Scotland: The Scottish Family Health Study (GS) is a family-based cohort study (N ∼ 24,000), investigating the genetic and environmental factors influencing health within Scotland (eMethods)^27,28^. Data collection including biological sampling occurred between February 2006 and March 2011. Written consent was provided before any assessment and/or sampling took place. Ethical approval was provided by the Tayside Research Ethics Committee (REC reference 05/S1401/89).

### Methylation data

DNAm was profiled from baseline blood samples using the Illumina MethylationEPIC array for 19,390 individuals. Methylation typing was performed in 4 distinct sets between 2017 and 2021. Sets 1, 2 and 4 included related individuals within and between each set while all individuals in set 3 were unrelated to each other and to individuals in set 1 (genetic relationship matrix (GRM) threshold <iJ0.05). Following quality control (QC) (eMethods), the sample sizes were Set 1: 5,087, Set 2: 459, Set 3: 4,450 and Set 4: 8,873 individuals. The sets were combined and dasen normalisation was performed across all individuals^29^. In total, 752,741 probes and 18,869 individuals passed QC. Beta-values were transformed to M-values using the ‘*beta2M()*’ function in the *lumi* R package^30^, and standardised using the ‘std- probe’ flag in the OSCA software^31^.

### Antidepressant exposure phenotypes

Prescription-derived antidepressant exposure was measured using antidepressant prescriptions from Public Health Scotland (eMethods), according to the British National Formulary (BNF) paragraph code ‘040303’, which largely corresponds to Anatomical Therapeutic Chemical (ATC) subclass ‘N06A’ (eTable 1) (N_prescriptions_ = 174,454, N_people_= 7,544) (eTable 2, eFigures 1-8). After removing ambiguous prescriptions (N_prescriptions_ = 5,484, N_people_ = 171) (eMethods, eTable 3), active treatment periods were defined by consecutive dispensing of antidepressant medications (eMethods, eFigures 9-11). Those who had their blood-draw appointment ≥ 7 days after treatment initiation or < 7 days after the end of a treatment period were defined as exposed (N_exposed_ = 861) (eFigure 12). Those with no antidepressant prescriptions on record were defined as unexposed (N_unexposed_ = 7,090).

Self-reported antidepressant exposure was derived from questionnaires sent 1–2 weeks before venepuncture (eMethods, eTable 4, eFigure 13). Those who did/did not self-report antidepressant use were defined as exposed and unexposed respectively (N_exposed_ = 1,508, N_unexposed_ = 15,028). Out of 6,473 individuals with both self-report and prescription-derived phenotypes, 6,355 exhibited concordant classification of antidepressant exposure (eFigure 14). The MDD-only phenotypes were derived by stratifying the samples to those with a lifetime MDD diagnosis, ascertained by the Structured Clinical Interview of the Diagnostic and Statistical Manual, version IV (SCID)^32^ (prescription-derived: N_exposed_= 380, N_unexposed_ = 412, self-report: N_exposed_= 766, N_unexposed_ = 1,502) (eMethods, eFigure 15).

### Methylome-wide association study

The MWAS were performed using a Mixed-linear-model Omics-based Analysis (MOA) in the OSCA software^31^. To account for relatedness within GS, each phenotype was regressed on a GRM^33^ using the Best Linear Unbiased Prediction (BLUP) tool in GCTA software^34^. The residuals were entered into a MOA model, which included a methylation omics- relatedness matrix as a random effect and age, sex, AHRR probe (cg05575921) M-values to proxy for smoking status, and predicted monocyte and lymphocyte cell proportions as fixed effects. Statistical significance was assessed using the p-value threshold 9.42x10^-8^, as recommended for case-control MWAS analyses^35^. Effect sizes represent a per-1 standard-deviation increase in CpG methylation M-values.

Differentially methylated regions (DMRs) were identified using the *dmrff* R package^36^, which performs an inverse-variance-weighted meta-analysis of MWAS beta and standard-error estimates per region, adjusting for estimate uncertainty and the correlational structure between probes. Candidate DMRs are identified as sets (>2) of CpGs <= 500bp apart with nominal significance (P < 0.05) and consistent effect direction. DMRs achieving Bonferroni-corrected p-value < 0.05 were considered statistically significant.

### Functional annotation

Gene-sets for both MWAS were collated by annotating the top 100 CpGs (by p-value) by the Infinium MethylationEPIC BeadChip database^37^. Hypergeometric tests, using ‘*phyper()*’, were used to assess the overlap of CpGs and gene-sets from both analyses. The background set consisted of CpGs and genes on or annotated to the EPIC array (eTable 5). The ‘GENE2FUNC’ analysis in functional mapping and annotation web-tool (FUMA) was used to assess enrichment of both gene-sets across 54 specific tissues in the Genotype-Tissue Expression (GTEx)^38,39^ Project. Both gene-sets were tested for enrichment in synapse-related GO terms using the SynGo web tool^40^ and for enrichment in GO: Biological Processes gene- sets (20 < ngenes < 500) in the msigdbr database^41^, using the *‘gsameth()*’ function from *missMethyl* R package^42^ (eTable 6).

### Methylation profile score

A least absolute shrinkage and selection operator (LASSO) penalised regression model was applied using *‘cv.biglasso()’* from the *glmnet* R package^43^ on the GS sample. First, the self-report phenotype was regressed against the GRM (using BLUP), age, sex, AHRR probe M-values as a proxy for smoking status, batch, and white blood cell (monocyte and lymphocyte) proportions. The residuals were used as the dependent variable and the standardised CpG sites on both EPIC and 450K Illumina arrays (N = 365,912) were included as independent variables. Ten-fold cross-validation was applied and the mixing parameter was set to 1. The non-zero weighted CpGs identified in the LASSO model were used to calculate a weighted-sum MPS in external datasets (FTC, SHIP-Trend, FOR2107, NTR, LBC1936, ALSPAC, MARS-UniDep and E-Risk, N_total_ = 9,578, N_exposed_ = 619) and a prospective sample of GS (STRADL, N_total_ = 658, N_exposed_ = 42) (eMethods). The association between antidepressant exposure and the MPS was assessed using; generalised linear mixed models, generalised linear models and generalised estimation equations, depending on the cohort’s population structure (i.e., twin studies vs unrelated participants) and DNAm pre-processing (eMethods, eTables 7-9). Age, sex, batch (where applicable), white blood cell proportions/counts and M-values at the AHRR probe were included as covariates. A random-effects meta-analysis using a DerSimonian-Laird estimator was performed to assess the overall association between the MPS and antidepressant exposure, using the *meta* R package^44^.

## Results

The demographic features of both antidepressant exposure phenotypes in the GS are shown in Table 1.

Table 1: Demographics of antidepressant exposed and unexposed individuals using the prescription-derived and self-reported measures in Generation Scotland. M = Mean, SD = Standard Deviation.

### Methylome-wide association studies

The self-report MWAS (Figure 2A, Table 2) and prescription-derived MWAS (Figure 2B, eTable 10) identified seven and four hypermethylated CpGs respectively, in those exposed to antidepressants (eFigure 16, eFigure 17). The effect estimates from all CpGs in both analyses were significantly correlated (R = 0.54, p < 2.2x10^-16^) (eFigure 18).

**Figure 2:**
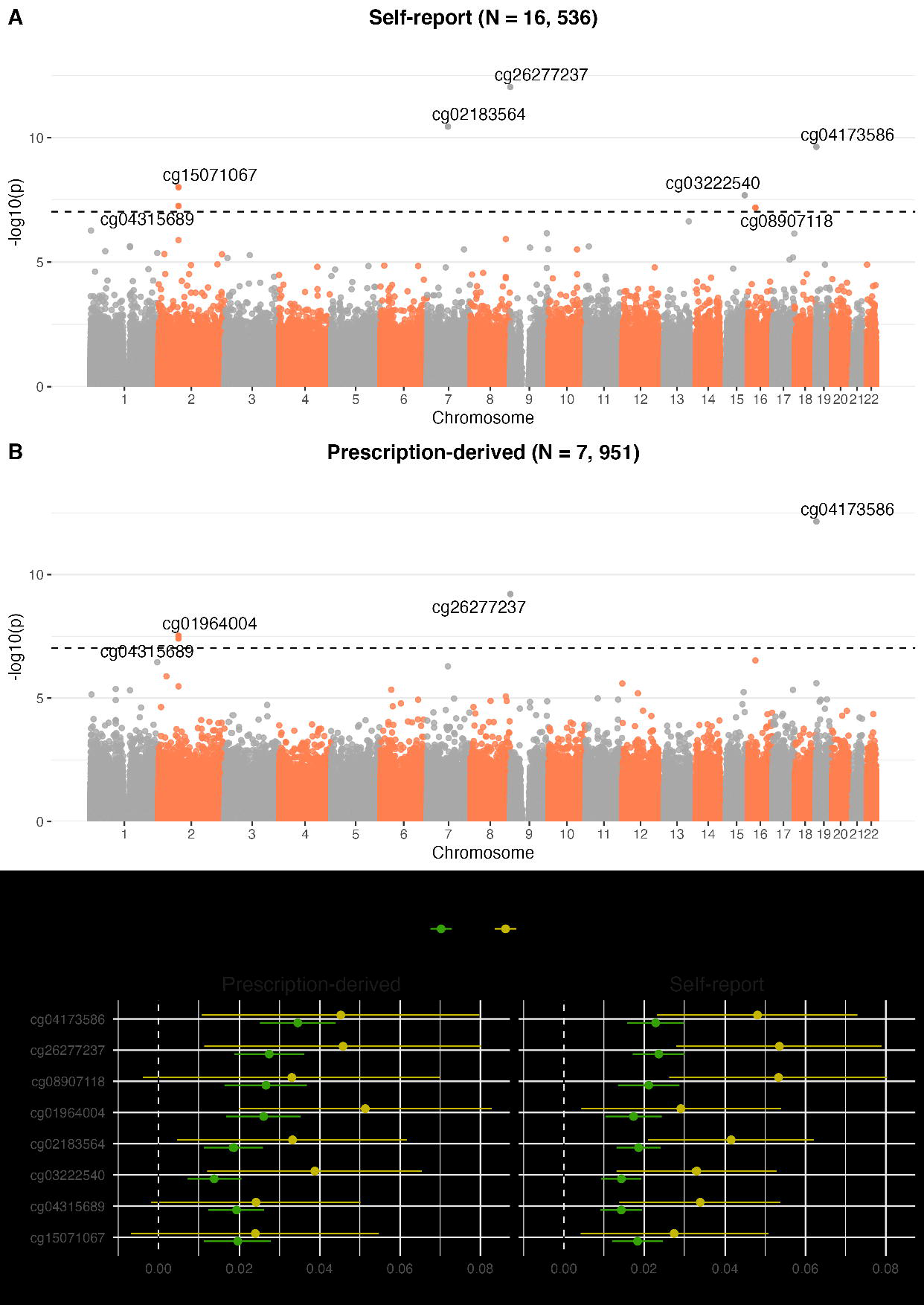
Methylome-wide association study of self-reported (A) and prescription-derived (B) antidepressant exposure. (C) The standardised effect sizes and 95% confidence-intervals for associated CpGs (p < 9.42x10^-8^) for the full sample (dark-green) and MDD-only sample (light-green).

Table 2: **Eight CpGs associated with self-reported and/or prescription-derived antidepressant use.** The EWAS catalog was searched using the *ewascatalog* R package^45^ for other studies (n > 1000) which report a significant CpG-trait association, accessed on 17/03/2024.

In the MDD-only self-report MWAS, only cg08527546 exhibited a significant association with antidepressant exposure (β = 0.050, p = 3.57x10-8); no CpGs were significantly associated in the MDD-only prescription-derived MWAS (eTable 11, eFigure 19). For both phenotypes, there was a significant correlation between CpG effect estimates in the full and MDD-only analyses (Rself-report = 0.57, pself-report < 2.2x10^-16^, Rprescription= 0.43, pprescription< 2.2x10^-16^) (eFigure 20). Notably, restricting the analyses to MDD cases did not attenuate the effect sizes of the significant CpGs (Figure 2C).

### Differentially methylated regions

There were 719,506 candidate regions considered in the dmrff analysis. The self-report MWAS had one significant DMR (f3 = 0.096, p_adj_ = 4.98x10^-3^) (eTable 12), consisting of two CpGs (cg01964004 and cg15071067), which maps to deoxyguanosine kinase antisense RNA1 (*DGUOK-AS1*) (eFigure 21). The prescription-derived MWAS identified no significant DMRs (eTable 13).

### Functional Annotation

The most significant 100 CpGs from the self-report and prescription-derived MWAS were annotated to 77 and 83 genes respectively (eTables 14-17). There was a significant overlap between both the CpG-sets (N_overlap_ = 17, p = 1.95x10^-48^, eFigure 22) and the gene-sets (N_overlap_ = 16, p =1.3x10^-25^, eFigure 23). The self-report gene-set was significantly enriched for the genes expressed in the amygdala (eTable 18, eFigure 24), whilst the prescription-derived gene-set had no significant enrichment (eTable 19, eFigure 25). There was significant enrichment of the self-report gene-set in GO:0008021;” synaptic vesicle membrane” (p_FDR_ = 0.030, SF26-28, ST20-23). There was no significant enrichment for any GO:Biological Processes pathways tested for either gene set (eFigure 29-30, eTable 24-25)

### Enrichment analysis: Netherlands Study of Depression and Anxiety

An MWAS of self-reported antidepressant exposure was performed in the NESDA cohort^46^ (N = 812, N_exposed_ = 398), with all participants having a recent (∼ 6 months) MDD DSM-IV diagnosis (eResults). The DNAm assays were obtained from whole blood samples using methyl-CG binding domain sequencing (MBD-Seq)^47–49^ (eResults). Enrichment tests were performed to assess the overlap of top findings from GS and NESDA, using the *shiftR* R package^50^. For both MWAS’, CpGs were filtered to those which showed a concordant direction of effect and three thresholds (0.1, 0.5 and 1%) were used to define top (by p-value) findings. Results suggested a small (OR: 1.39) but significant (P < 0.042) enrichment between the top 1% of results from both MWAS’.

### Methylation differences by time in treatment

To investigate the relationship of DNAm at significantly associated CpGs (n = 8) with the length of antidepressant exposure, a Spearman correlation test was performed between the DNAm levels and time in current treatment for those within a treatment period (N = 863). Two probes, cg15071067 (*DGUOK-AS1)* and cg26277237 (*KANK1)*, showed a significant correlation between methylation and time in treatment (cg15071067:p= 0.085, p = 0.012, cg26277237: p= 0.087, p = 0.011) (eFigures 31-33, eTable 26), with the same direction of effect found in the MWAS.

### Methylation Profile Score

There were 212 CpGs identified by the LASSO regression model (eFigures 34-36, eTable 27), used to calculate the MPS in external cohorts (Figure 3A, eFigures 37-46). All cohorts bar one (NTR) showed a positive relationship between antidepressant MPS and antidepressant exposure (f3_FTC_ = 0.156, f3_SHIP-Trend_ = 0.134, f3_STRADL_ = 0.149, f3_LBC1936_ = 0.228, f3_FOR2107_ = 0.349, f3_MARS_= 0.263, f3_ALSPAC_= 0.170, f3_ERISK_= 0.342, f3_NTR_= -0.031) (eFigure 47, eTable 28).

**Figure 3:**
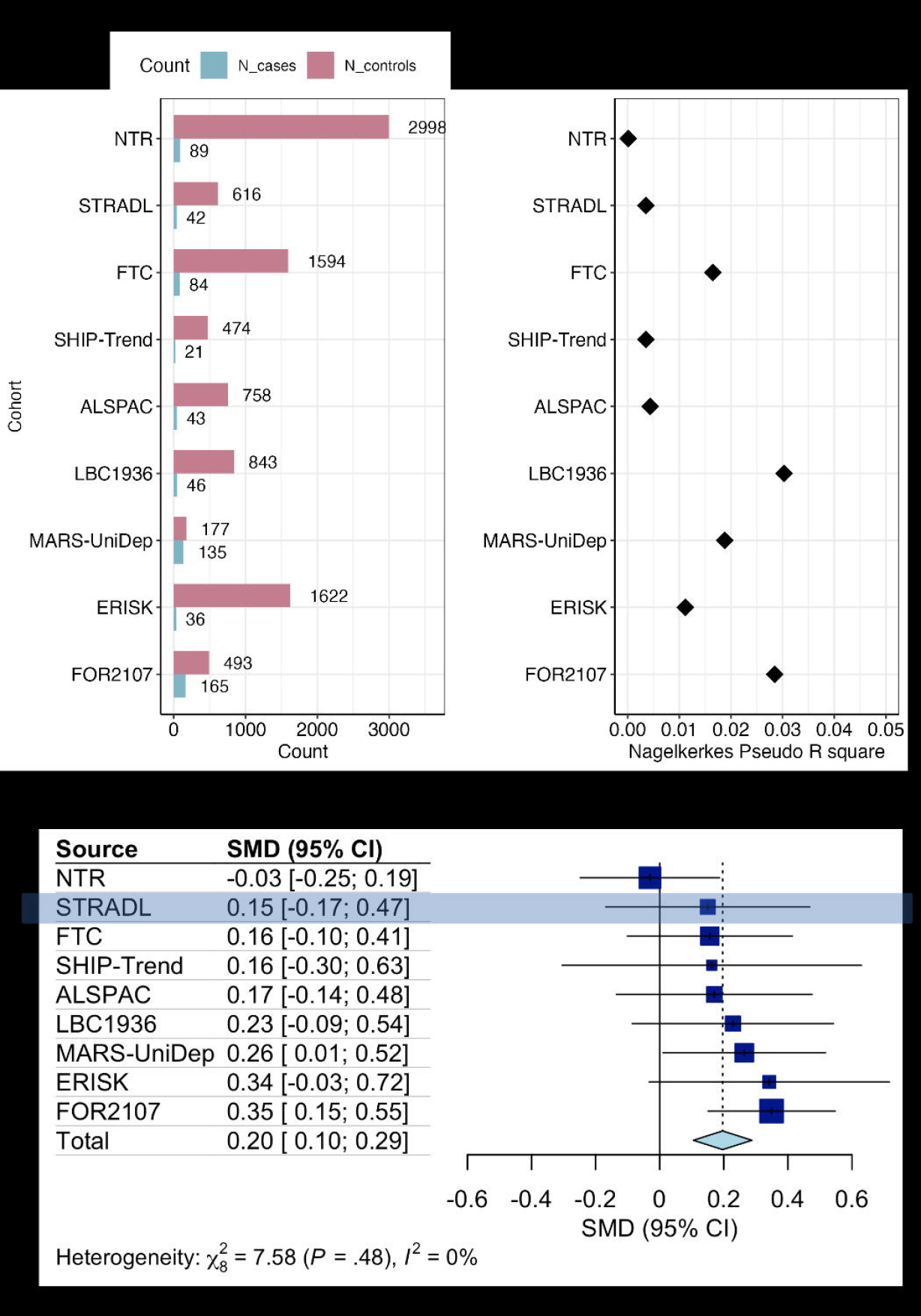
Antidepressant exposure ∼ MPS in external cohorts. A) The sample sizes of each dataset, **B)** Nagelkerke’s pseudo R^2^, **C)** The effect sizes and confidence intervals of MPS ∼ antidepressant exposure analysis in each cohort. Square size = study weight, diamond = meta-analysed effect.

Nagelkerke’s pseudo R^2^ estimates ranged from 1.11x10^-3^ (NTR) to 0.03 (LBC1936) (Figure 3B). The random-effects meta-analysis found a significant association between antidepressant exposure and the MPS (pooled f3 [95%CI]: 0.196 [0.105, 0.288], p < 1x10^-4^), with low heterogeneity between studies (I^2^ [95%CI] = 0% [0, 64.8%]) (eTable 29).

## Discussion

This study presents the largest investigation of the impact of antidepressant exposure on the methylome^51^. The results from self-report or prescription-derived antidepressant exposure were broadly consistent, corroborating previous findings^26^. There was evidence of hypermethylation at eight CpGs and a region on Chromosome 2 (BP: 74196550-74196572) in those exposed to antidepressants. The CpG with the highest significance and the largest effect size, cg26277237, mapped to KN motif and ankyrin repeat domains 1 (*KANK1*), was previously reported by Barbu *et al.*^21^ on a smaller sample of GS. The DMR analysis indicated antidepressant exposure is also significantly associated with hypermethylation near *DGUOK- AS1*, a long non-coding RNA (lncRNA). *KANK1* facilitates the formation of the actin cytoskeleton and has an active role in neurite outgrowth and neurodevelopment^52^. A meta-analysis of copy-number variant association studies found a significant duplication in *KANK1* in those with five different neurodevelopmental disorders, including MDD^53^. *DGUOK-AS1* has an inhibitory role on the expression of a nearby gene *DGUOK*^54^, which encodes a mitochondrial enzyme involved in the production of mitochondrial DNA^54^, and has previously been implicated as a risk gene in schizophrenia^55^ and Alzheimer disease^56^. A recent review reported evidence that antidepressants do influence mitochondrial function, although the effects are heterogeneous between different types of antidepressants, independent of their current classification^57,58^.

Seven of the CpGs significantly hypermethylated with antidepressant use have been reported previously to also be significantly hypermethylated with incident and/or prevalent type 2 diabetes (T2D)^59^ in GS. Previous epidemiological studies have indicated that antidepressant use leads to an increased risk of T2D onset in a time- and dose-dependent manner^60,61^. Future prospective and longitudinal research into the link between antidepressant use, DNAm and T2D, alongside the use of other independent datasets is required.

The performance of the GS-trained MPS in discriminating antidepressant exposure across eight external datasets, demonstrates that this may be a generalizable biomarker indicative of antidepressant exposure and adds to a growing set of MPS that could potentially provide clinically relevant phenotypic information^62–65^. Accurate estimation of this exposure history could be highly valuable for epidemiological studies where prescribing data may not be available. Taken in combination with MPS for other risk factors, an MPS for antidepressant exposure may help provide a robust characterisation of an individual’s medical history.

There are several strengths of this study. The comparison of self-report and prescription-derived measures is valuable, as the former is often cheaper and easier to obtain in large-scale cohort studies^25^. Furthermore, the MDD-only analysis indicates that the hypermethylation associated with antidepressant use is not driven by MDD indication. Additionally, the significant association of an MPS trained in GS with antidepressant exposure in external datasets, and the significant enrichment with an independent MWAS consolidates our findings.

This study has various limitations. Both measures of antidepressant exposure do not discriminate between antidepressant drugs, classes, or dosages. However, we anticipate the opportunity to investigate more medication-specific effects on the methylome using prescription-linkage data as biobanks increase in size. Additionally, all the cohorts used primarily consist of European ancestry. It is paramount that this analysis is conducted in non-European ancestral groups to further verify our findings and disentangle any ancestry-specific effects^66–68^. Finally, by design, this epidemiological study cannot directly address causality between antidepressant exposure and DNAm. The integration of DNAm analysis into randomised controlled trials of antidepressants is important to establish the exact nature of the association and to inform potential new targets for antidepressant therapy.

This study indicates that antidepressant exposure is associated with hypermethylation at *DGUOK-AS1* and *KANK1*, which have roles in mitochondrial metabolism and neurite outgrowth respectively. Future research should include more cohorts of non-European ancestry, alongside the incorporation of DNAm in randomised trials of antidepressants to further consolidate findings and establish causality. If replicated, targeting of these genes could inform the design of more effective and better tolerated treatments for depression.

## Data Availability

Summary statistics from the MWAS analyses are being submitted for open access on the EWAS catalog. All code and analysis scripts are available on GitHub (https://github.com/Elladavyson).

https://github.com/Elladavyson

https://www.ewascatalog.org/

## Acknowledgments

This work has made use of the resources provided by the Edinburgh Compute and Data Facility (ECDF, 2024) (http://www.ecdf.ed.ac.uk/).

## Generation Scotland (GS) & Stratifying Resilience and Depression Longitudinally (STRADL)

We thank the participants in Generation Scotland for making this research possible. We also acknowledge the team at Generation Scotland for collecting and preparing the data for analyses.

## FOR2107

We are deeply indebted to all study participants and staff. A list of acknowledgments can be found here: www.for2107.de/acknowledgements.

## Netherlands Twin Register (NTR)

We warmly thank all twin families of the Netherlands Twin Register who make this research possible.

## Munich Antidepressant Response Study / UniPolar Depression Study (MARS-UniDep)

We would like to thank all contributors to the research project including physicians, psychologists, study nurses, researchers and research assistants, and of course patients of the hospital of the Max Planck Institute of Psychiatry in Munich and psychiatric hospitals in Augsburg and Ingolstadt.

## Avon Longitudinal Study of Parents and Children (ALSPAC)

We thank all the families who took part in this study, the midwives for their help in recruiting them, and the whole ALSPAC team, which includes interviewers, computer and laboratory technicians, clerical workers, research scientists, volunteers, managers, receptionists and nurses.

## E-risk Longitudinal Twin Study (E-risk)

We are grateful to the E-Risk study mothers and fathers, the twins, and the twins’ teachers for their participation. Our thanks to Professors Terrie Moffitt and Avshalom Caspi, the founders of the E-Risk study, and to the E-Risk team for their dedication, hard work, and insights.

## GS & STRADL

This work was supported by 3 Wellcome Trust grants (220857/Z/20/Z, 226770/Z/22/Z and 104036/Z/14/Z [AMM]). Funding from the Biotechnology and Biological Sciences Research Council and Medical Research Council (Grant Nos. MR/X003434/1 and MR/W014386/1) and the European Union (Grant agreement 847776) is also gratefully acknowledged. The Psychiatric Genomics Consortium (including AMM) has received major funding from the U.S. National Institute of Mental Health (Grant No. 5 U01MH109528-03).

## Finnish Twin Cohort (FTC)

Phenotype and genotype data collection in FT12 and FT16 studies of the Finnish twin cohort has been supported by the Wellcome Trust Sanger Institute, the Broad Institute, ENGAGE – European Network for Genetic and Genomic Epidemiology, FP7-HEALTH-F4-2007, grant agreement number 201413, National Institute of Alcohol Abuse and Alcoholism (grants AA- 12502, AA-00145, and AA-09203 to R J Rose; AA15416 and K02AA018755 to D M Dick; R01AA015416 to J Salvatore) and the Academy of Finland (grants 100499, 205585, 118555, 141054, 264146, 308248, 265240, 263278 to J Kaprio, 328685, 307339, 297908 and 251316 to M Ollikainen, and Centre of Excellence in Complex Disease Genetics grants 312073, 336823, and 352792 to J Kaprio), NIH/NHLBI (grant HL104125 to X Wang), and Sigrid Juselius Foundation to M Ollikainen and J Kaprio.

## Study of Health in Pomerania (SHIP-Trend)

SHIP is part of the Community Medicine Research net of the University of Greifswald which is funded by the Federal Ministry of Education and Research (01ZZ9603, 01ZZ0103, and 01ZZ0403), the Ministry of Cultural Affairs and the Social Ministry of the Federal State of Mecklenburg-West Pomerania. DNA methylation data have been supported by the DZHK (grant 81X3400104). The University of Greifswald is a member of the Caché Campus program of the InterSystems GmbH.

## FOR2107

The German multicenter consortium “Neurobiology of Affective Disorders. A translational perspective on brain structure and function” is funded by the German Research Foundation (Research Unit FOR2107). Principal investigators are Tilo Kircher (speaker FOR2107, DFG grant numbers KI588/14-1, KI588/14-2, KI588/20-1, KI588/22-1, KI 588/15-1, KI 588/17-1), Udo Dannlowski (co-speaker FOR2107; DA1151/5-1, DA1151/5-2, DA1151/6-1), Axel Krug (KR3822/5-1, KR3822/7-2), Igor Nenadic (NE2254/1-2, NE2254/2-1, NE2254/3-1, NE2254/4-1), Carsten Konrad (KO4291/3-1), Marcella Rietschel (RI 908/11-1, RI 908/11-2), Markus Nöthen (NO 246/10-1, NO 246/10-2), Stephanie Witt (WI 3439/3-1, WI 3439/3-2). Tilo Kircher received unrestricted educational grants from Servier, Janssen, Recordati, Aristo, Otsuka, neuraxpharm.

## NTR

This work was supported by the Royal Dutch Academy for Arts and Science (KNAW) Academy Professor Award (PAH/6635) to DIB; the Netherlands Organization for Scientific Research (NWO 480-15-001/674) and Biobanking and Biomolecular Resources Research Infrastructure (BBMRI-NL: 184.021.007; 184.033.111).

## MARS-UniDep

The MARS cohort was sponsored by the Max Planck Society. The UniDep cohort was funded by the Bavarian Ministry of Commerce and by the Federal Ministry of Education and Research in the framework of the National Genome Research Network, Foerderkennzeichen 01GS0481 and the Bavarian Ministry of Commerce. DNA methylation analysis of a subset of both cohorts was financed by ERA-NET NEURON.

## Lothian Birth Cohort 1936 (LBC1936)

The LBC1936 is supported by the Biotechnology and Biological Sciences Research Council, and the Economic and Social Research Council [BB/W008793/1] (which supports SEH), Age UK (Disconnected Mind project), the Medical Research Council [G0701120, G1001245, MR/M013111/1, MR/R024065/1], the Milton Damerel Trust, and the University of Edinburgh. SRC is supported by a Sir Henry Dale Fellowship jointly funded by the Wellcome Trust and the Royal Society (221890/Z/20/Z). Methylation typing of was supported by Centre for Cognitive Ageing and Cognitive Epidemiology (Pilot Fund award), Age UK, The Wellcome Trust Institutional Strategic Support Fund, The University of Edinburgh, and The University of Queensland.

## ALSPAC

The UK Medical Research Council and Wellcome (Grant Ref: 217065/Z/19/Z) and the University of Bristol provide core support for ALSPAC. ASFK is funded by a Wellcome Early Career Award (Grant ref: 227063/Z/23/Z). A comprehensive list of grants funding is available on the ALSPAC website (http://www.bristol.ac.uk/alspac/external/documents/grant-acknowledgements.pdf). This publication is the work of the authors and ED, PY & ASFK will serve as guarantors for the ALSPAC contents of this paper. PY’s work is supported by the National Institute for Health and Care Research Bristol Biomedical Research Centre and the Medical Research Council Integrative Epidemiology Unit at the University of Bristol (MC_UU_00032/4, MC_UU_00032/6). The views expressed are those of the authors and not necessarily those of the NIHR or the Department of Health and Social Care.

## E-Risk

The E-Risk Study is funded by grants from the UK Medical Research Council [G1002190; MR/X010791/1]. Additional support was provided by the US National Institute of Child Health and Human Development [HD077482] and the Jacobs Foundation. Chloe C. Y. Wong was supported by the National Institute for Health and Care Research (NIHR) Maudsley Biomedical Research Centre at South London and Maudsley NHS Foundation Trust and King’s College London [NIHR203318]. Helen L. Fisher was supported by the Economic and Social Research Council (ESRC) Centre for Society and Mental Health at King’s College London [ES/S012567/1]. The views expressed are those of the authors and not necessarily those of the NIHR, the Department of Health and Social Care, the ESRC, or King’s College London.

## Netherlands Study of Depression and Anxiety

The NESDA study is supported by the Geestkracht program of the Netherlands Organization for Health Research and Development (Zon-Mw, grant number 10–000-1002) and the participating institutions (VU University Medical Center, Leiden University Medical Center, University Medical Center Groningen. The current methylation project was supported by grant R01MH099110 from the National Institute of Mental Health. The sponsors had no role in the design and conduct of the study; collection, management, analysis, and interpretation of the data; preparation, review, or approval of the manuscript; or decision to submit the manuscript for publication.

