## Supplementary Information for "Antidepressant Exposure and DNA Methylation: Insights from a Methylome-Wide Association Study"

Davyson et al.

### Methods

### Study Population: Generation Scotland

Protocol Paper: <https://doi.org/10.1093/ije/dys084>^1^

In depth information regarding the recruitment and assessment of participants in Generation Scotland (GS) can be found elsewhere^2^. In brief, participants over the age of 18 years were randomly recruited through general practice NHS surgeries within Scotland from 2006 to 2011. Participants filled out a pre-clinical questionnaire, and subsequently attended an extensive in-person clinic where physical measurements, biological sampling, and a psychiatric assessment was performed. Participants were informed that the nature of the study was to investigate the health of the Scottish population and gave consent before any sampling took place.

### Methylation data: DNAm quality control

DNAm profiling of the GS samples was carried out by the Genetics Core Laboratory at the Edinburgh Clinical Research Facility, Edinburgh, Scotland. Quality control and normalisation steps were applied to each wave separately. Quality control in the first wave was performed using *shinyMethyl* R package^3^. Outliers were removed based on visual inspection of the log median intensity of methylated vs unmethylated signals. Samples were removed for which DNAm-predicted sex did not match recorded sex. The following were removed using the *wateRmelon* R package^4^: i) samples where >1% of CpGs have detection p-value > 0.05, ii) probes with a beadcount < 3 in > 5% of samples; and iii) probes for which >0.5% of samples had a detection p value > 0.05. Proportion of white blood cells were estimated using the houseman method implemented by the *minfi* R package^5^, using the reference data from Reinius et al, (2012)^6^. Quality control steps on wave 2-4 were performed by the R package *meffil*^7^ and *shinyMethyl*. Dye bias and background correction was performed using *meffil* and the *‘noob’* method^8^ to exclude: samples with strong dye bias and/or issues with bisulphite conversion, samples with outlying median methylated signal intensity and samples where DNAm predicted sex did not match recorded sex. *Meffil* was used to remove samples with > 0.5% of CpG sites with detection p value > 0.01, alongside probes which had a beadcount < 3 in > 5% of samples and/or had a detection p-value >0.01 in > 1% of samples. Proportion of white blood cells were estimated using the houseman method implemented by *minfi*, using the reference data from Reinius et al, (2012)^6^. Multi-dimensional scaling (MDS) was performed, and visual inspection identified any outlying samples which were removed. Following quality control, probes on the Y and X chromosome were removed, alongside those which are predicted to bind poorly according to either Zhou et al, (2017)^9^ or McCartney et al, (2016)^10^. Following quality control, there were 5,087, 459, 4,450 and 8,873 individuals within set 1, 2, 3, and 4 respectively. The sets were combined and dasen normalisation was performed across all individuals, using *wateRmelon*. The dasen-normalised beta-values were converted to M-values using the *‘beta2M()’* function in the *lumi* R package^11^.

### Antidepressant exposure phenotypes

#### Prescription-derived antidepressant exposure

Consent was obtained from 23,603 participants for their data in GS to be linked to the National Health Service (NHS) records, through their Community Health Index (CHI) number. An individual’s CHI number is routinely used by NHS Scotland for various procedures, including prescription events^12^. Prescription data for these individuals was obtained from the Scottish National Prescribing Information System (PIS)^13^, which gives access to individual prescribing and dispensing events in a community setting since 2009. Further information about the PIS database can be found elsewhere^13^. There was a total of 2,841,726 prescription events for 21,318 individuals, with dispensing dates spanning April 2009 to January 2017. A prescription data-entry had extensive information including the medication name, formulation, quantity, strength, BNF codes, dispense data, paid date, and free-text prescriber instructions.

Antidepressant prescriptions selected included serotonin reuptake inhibitors (SSRIs) (Fluoxetine, Sertraline, Paroxetine, Citalopram, Escitalopram and Fluvoxamine Maleate), monoamine oxidase inhibitors (MAOs) (Moclobemide, Phenelzine, Tranylcypromine), tricyclic antidepressants (TCAs) (Amitriptyline, Clomipramine, Dosulepin, Doxepin, Imipramine, Lofepramine, Mianserin, Nortriptyline, Trazodone, Trimipramine) and other antidepressants (Agomelatine, Mirtazapine, Reboxetine, Tryptophan, Venlafaxine). Exact numbers of each antidepressant are shown in Figure **1**. The vast majority (n = 173,818) of the prescriptions were for tablets or capsules, and the rest were in liquid forms (n = 636).

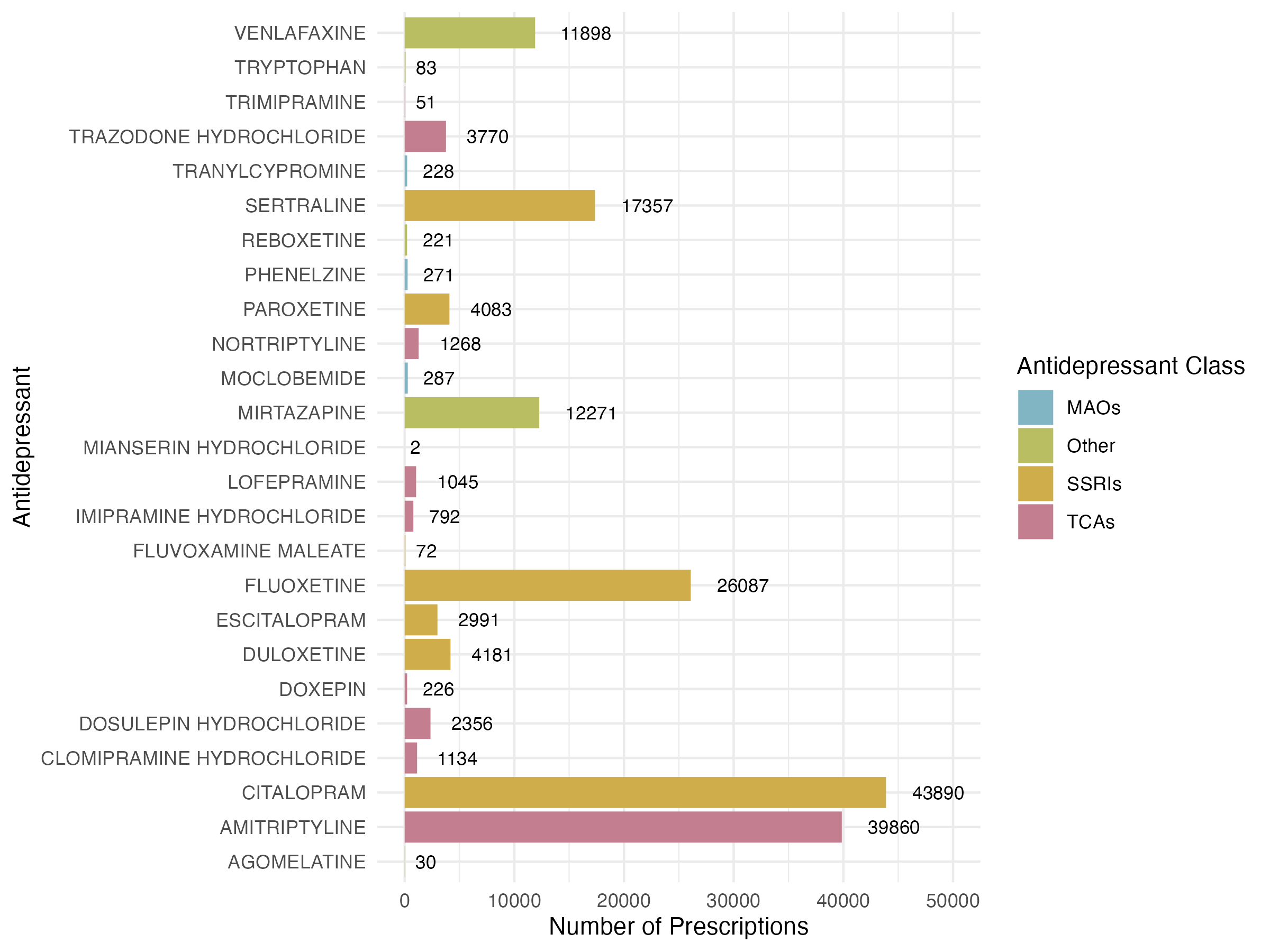

**Figure** **1:** All antidepressant prescriptions for individuals in Generation Scotland between 2009 and 2017.

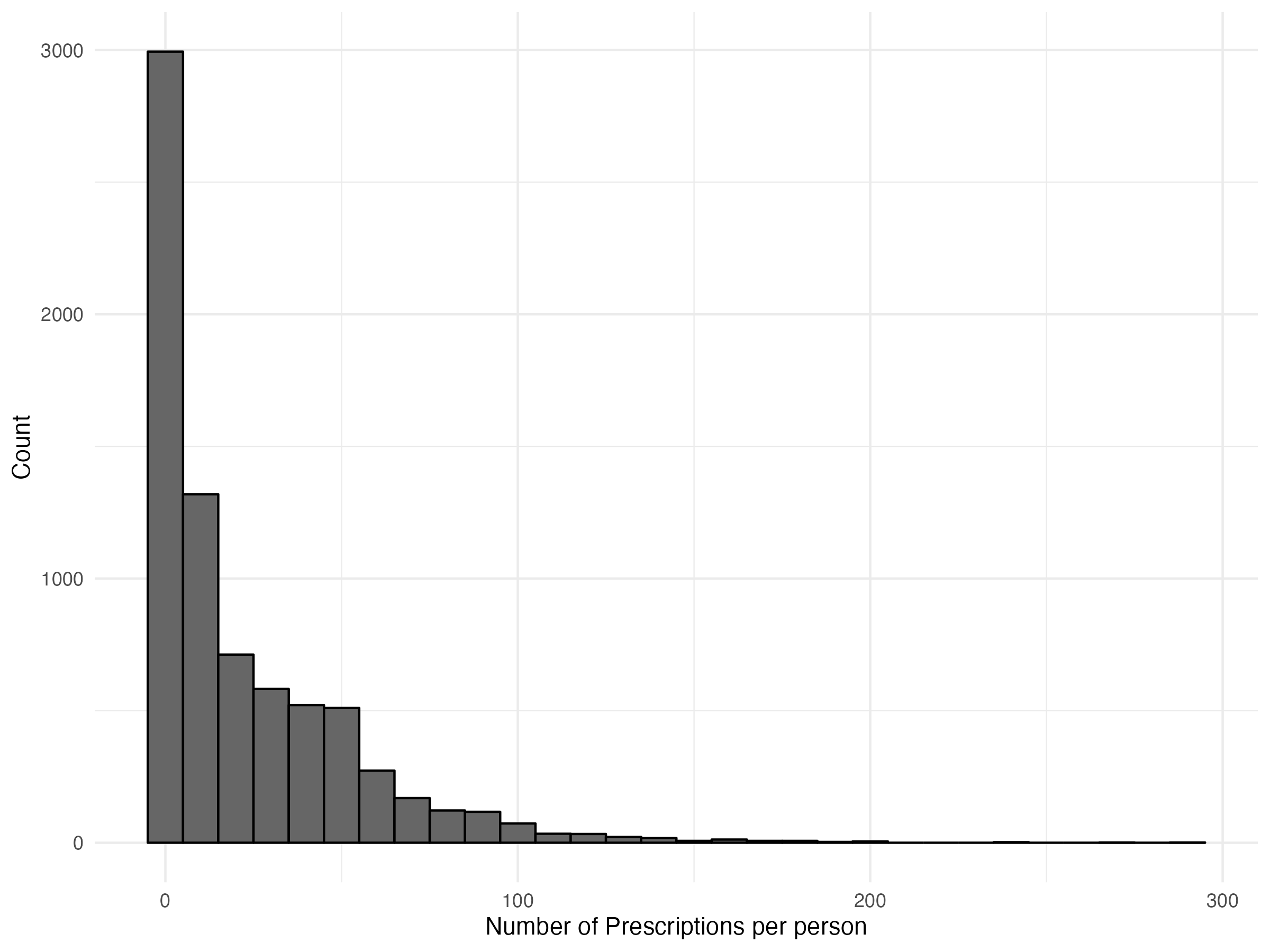

**Figure** **2:** Histogram of the number of antidepressant prescriptions per person between 2009-2017.

##### Drug Dosages

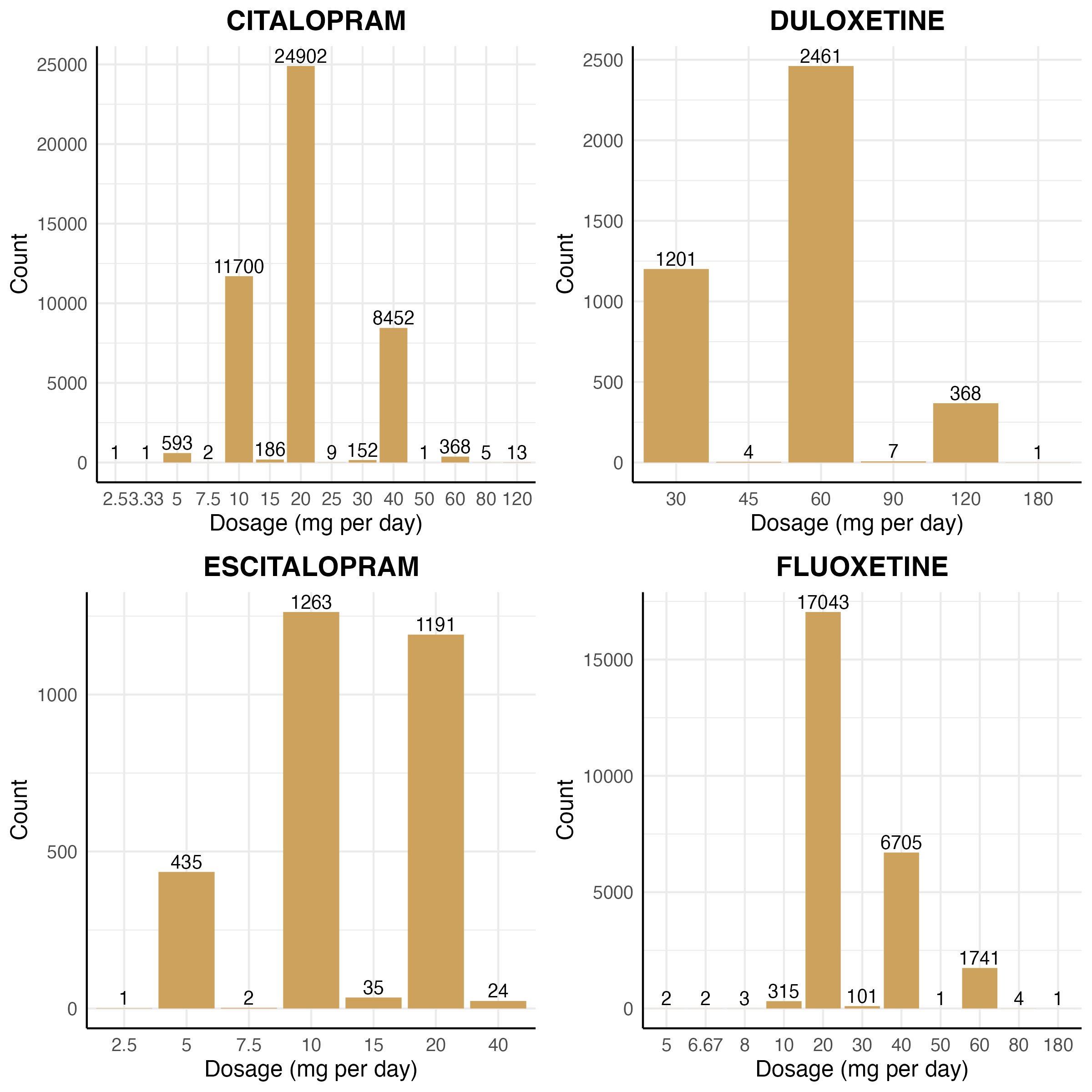

**Figure** **3:** Prescription dosages for each antidepressant medication after parsing prescription instructions.

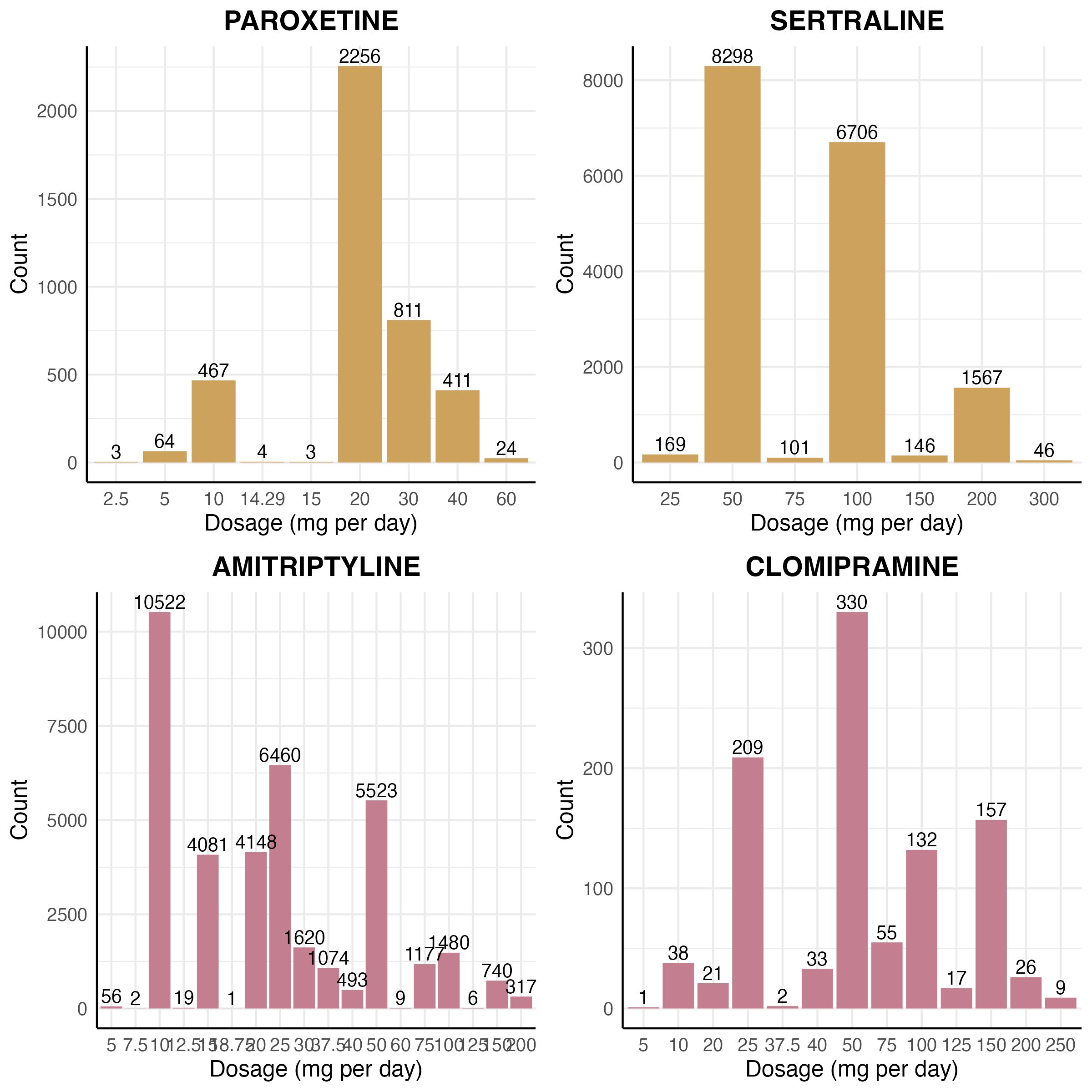

**Figure** **4:** Prescription dosages for each antidepressant medication after parsing prescription instructions.

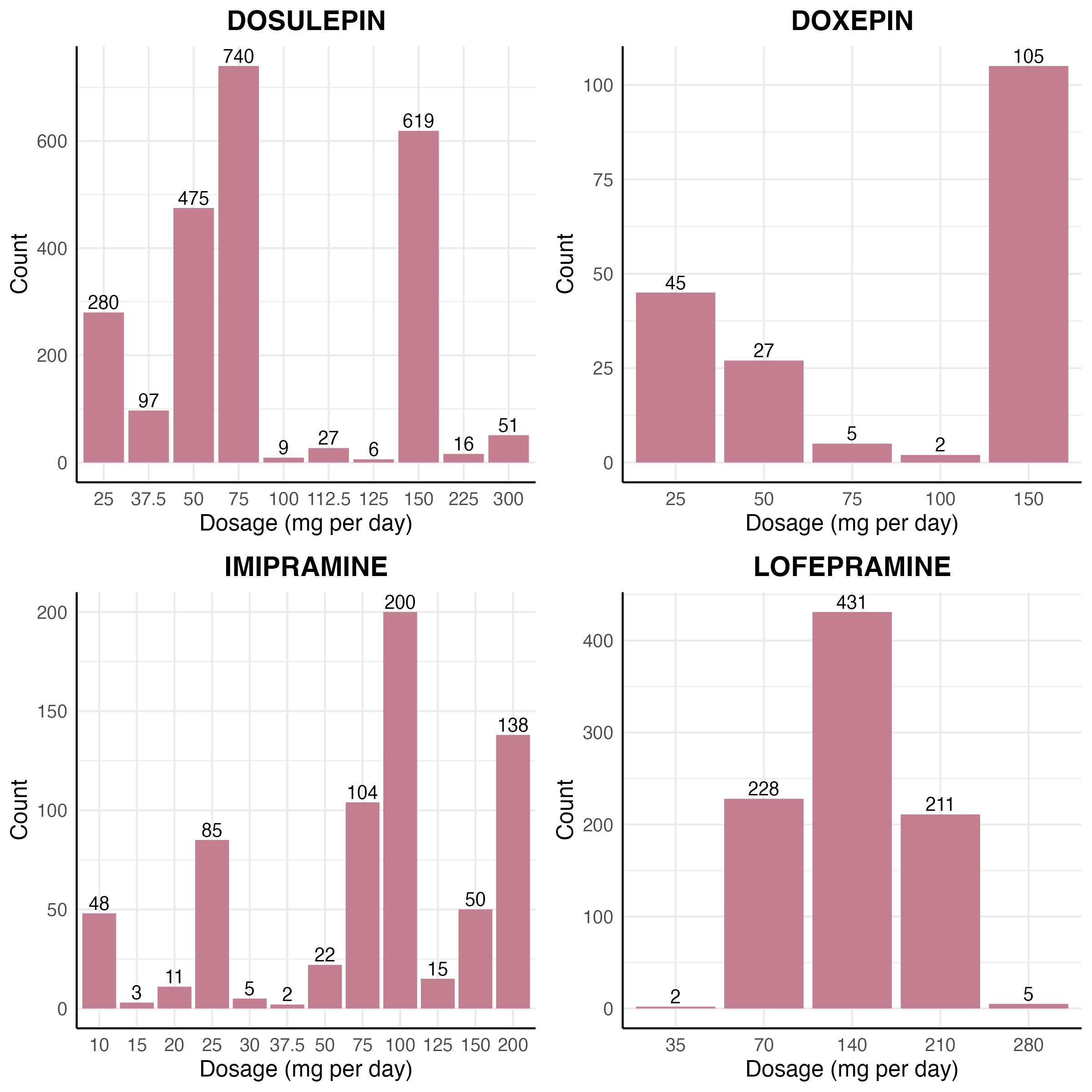

**Figure** **5:** Prescription dosages for each antidepressant medication after parsing prescription instructions.

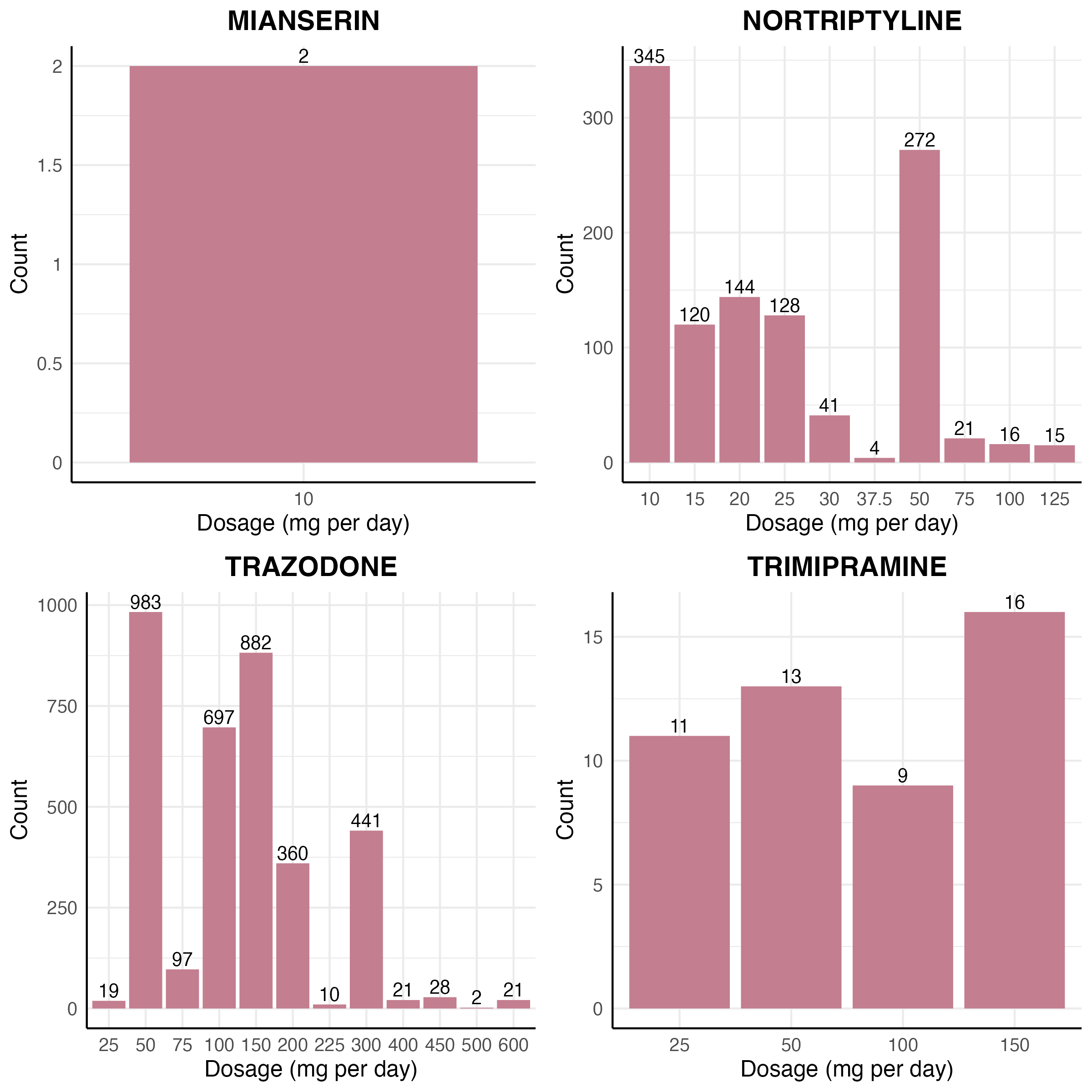

**Figure** **6:** Prescription dosages for each antidepressant medication after parsing prescription instructions.

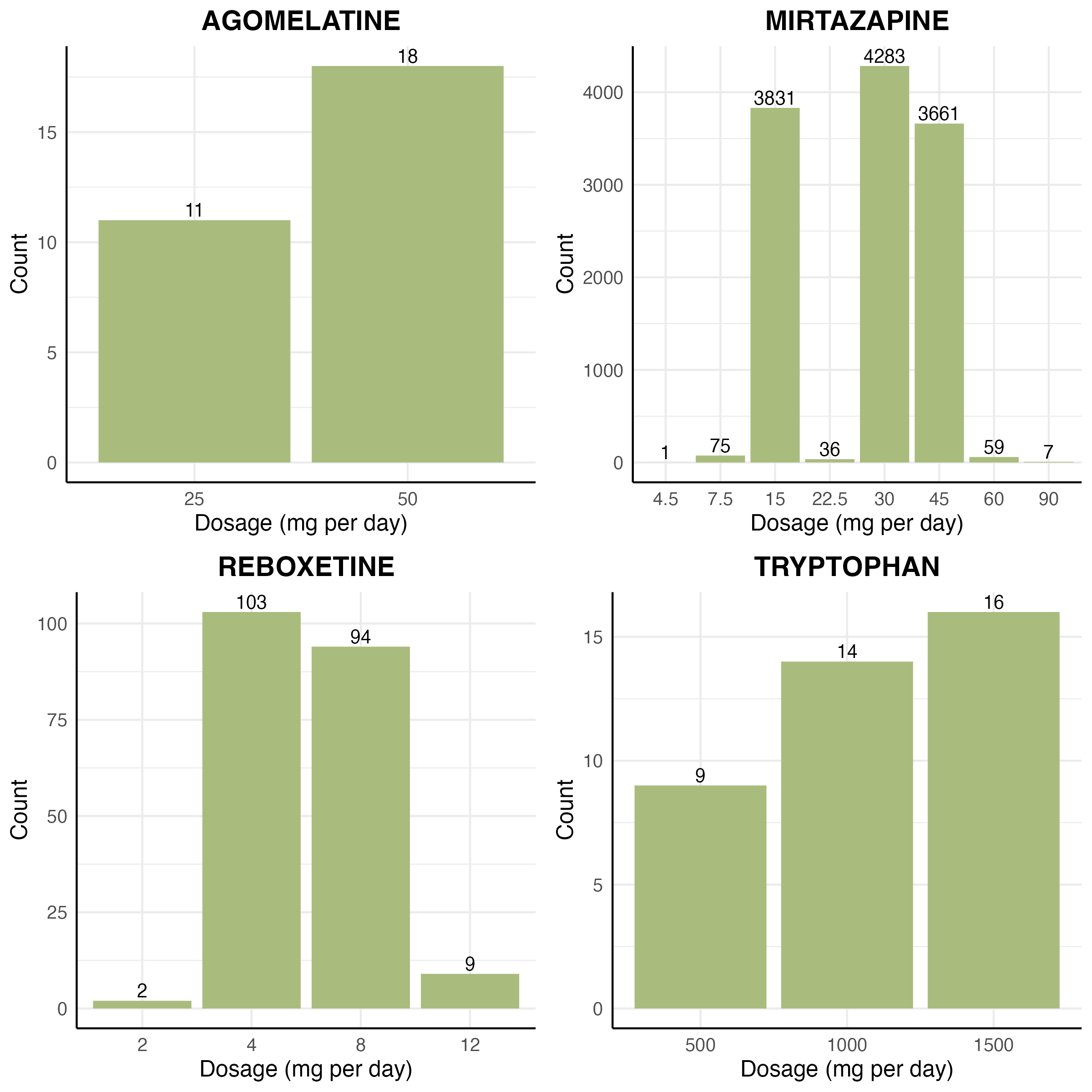

**Figure** **7:** Prescription dosages for each antidepressant medication after parsing prescription instructions.

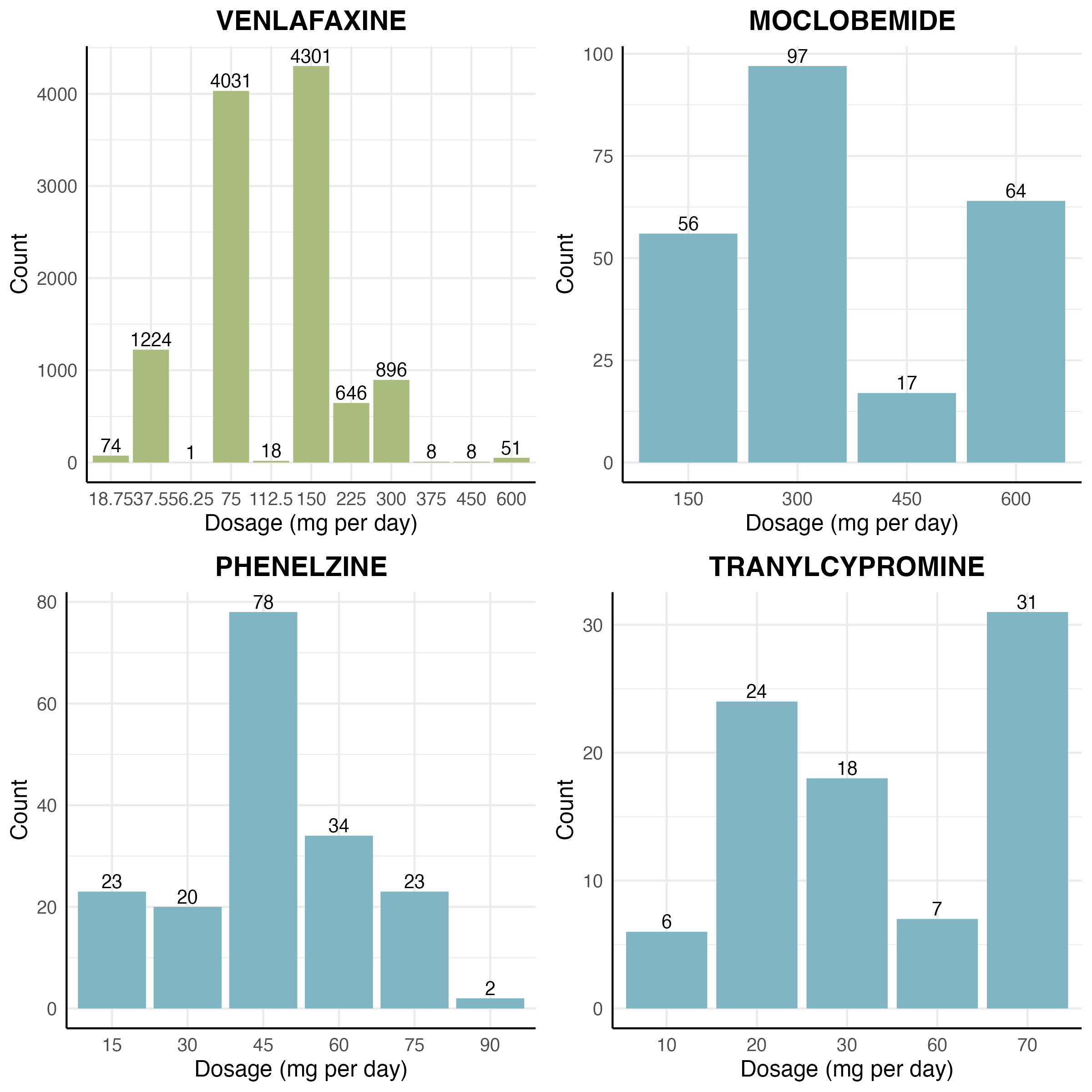

**Figure** **8:** Prescription dosages for each antidepressant medication after parsing prescription instructions.

##### Ambiguous prescription instructions

Prescriptions with ambiguous or dosage-irrelevant instructions, e.g ‘as directed’, ‘as instructed’, ‘taken in the morning’ (n = 9216) were identified by the regex expressions: *^$_, _^(to be taken\s*|(for\s*)?use\s*|take\s*)?as (dir|directed|advised|discussed).*(to be taken\s*|(for\s*)?use\s*|take\s*)?$* and *^non formulary please choose citalopram instead$*. For the prescriptions in capsule and tablet form (N_prescriptions_ =9187), the instruction was assumed to be a default of 1 tablet a day. For the solutions (n = 636), ambiguous prescriptions (n = 48) were imputed to be the same as the most common prescription instruction given to the individual in the previous year, if there were no previous prescriptions (n = 6), then the prescription was discarded from the dataset.

Additionally, prescriptions which had instructions for titrating or reducing dosages e.g ‘take 2 a day for a week, then 1, then stop’ (n_tablets_ = 5430, 3.12% n_liquids_ = 48, 7.55%) were discarded from the analysis.

##### Multiple prescriptions on the same day insert

2, 604 people received multiple (1+) prescriptions on the same dispense date (N_prescriptions_ = 37, 817). We theorised that this could be due to two possibilities:

- **Scenario 1) Same continuous treatment**: each prescription is for the continuation of the same treatment and are being collected ahead of time.
- **Scenario 2) Different simultaneous treatment**: each prescription aligns to a separate treatment and are to be taken simultaneously.

We distinguish between each scenario using a decision tree, illustrated in Supplementary Figure **9**. In brief, prescriptions assumed to be for the **scenario 1** (same continuous treatment) if a) prescriptions are for the same medication and the same dosage, or b) medications for the same medication and different dosages but the combined dosage is over the maximum recommended dosage as we theorise that this would not be prescribed to be taken simultaneously. If **scenario 1** is met, then the multiple prescriptions are merged together to one prescription event (i.e 2 prescriptions, each with 28 tablets, would be merged to be one prescription with 56 tablets) (N_prescriptions_= 10, 016). Prescriptions are assumed to be for **scenario 2** (separate simultaneous treatments) if a) the prescriptions are for different medications, or b) the prescriptions are for the same medication and different dosages, but the combined dosage is under the maximum recommended dosage. If it is determined to be **scenario 2)**, then we treat each prescription as independent and do not merge the prescription events (N_prescriptions_= 27, 801).

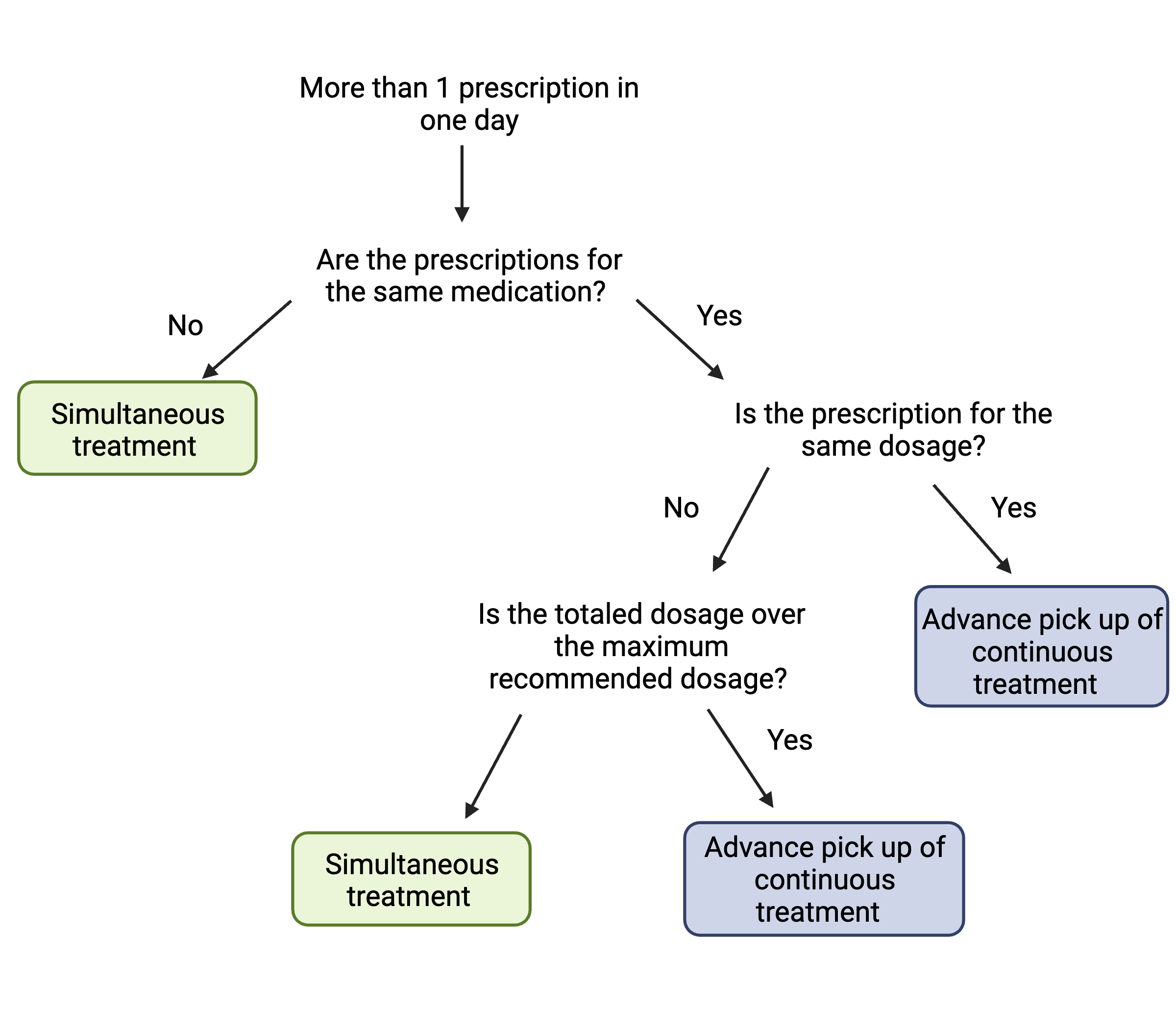

**Figure** **9:** Decision tree of marking multiple prescriptions dispensed on the same day as simultaneous treatment or continuous treatment collected in advance, which are treated as independent and merged prescription events respectively.

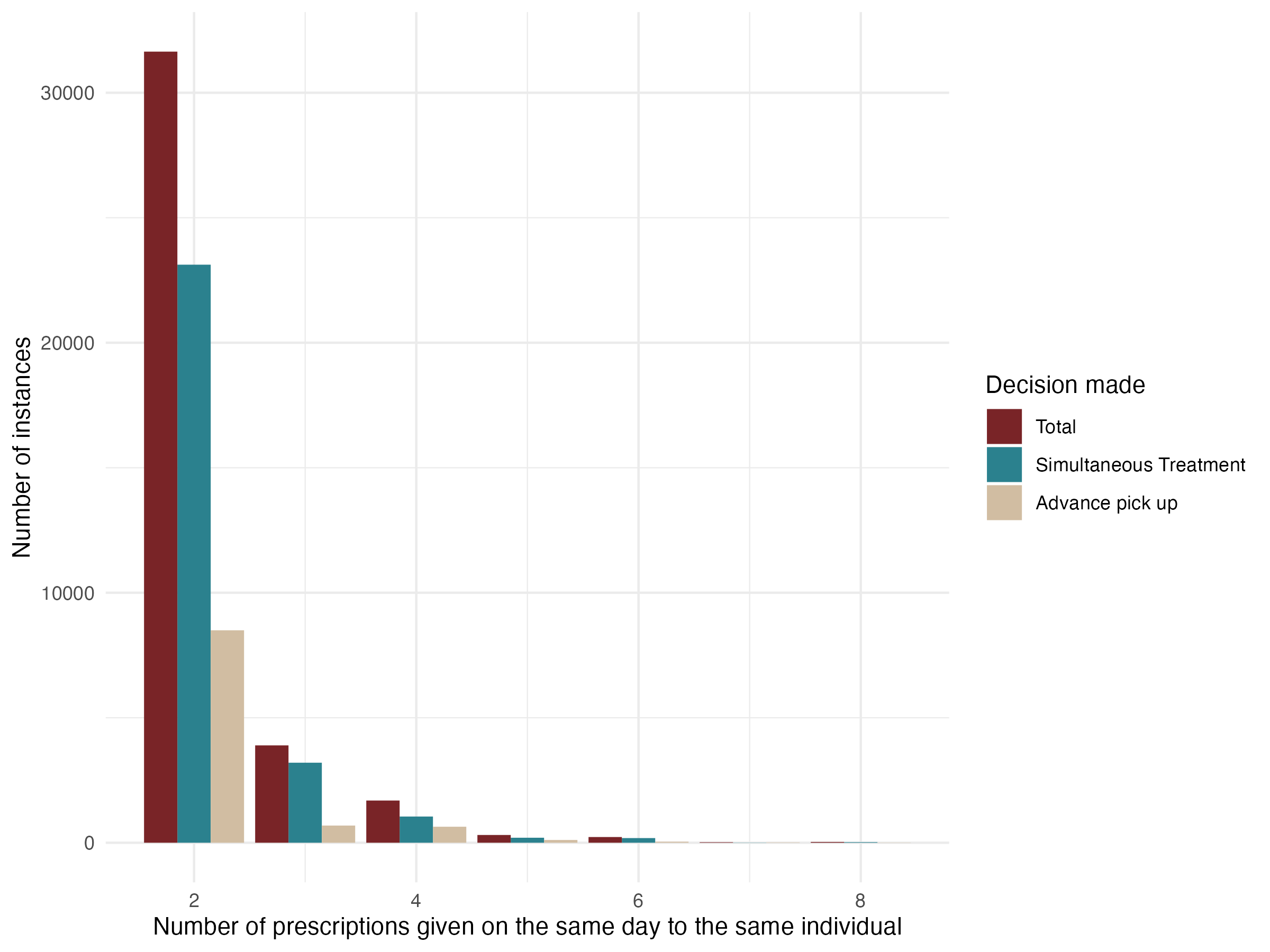

**Figure** **10:** Bar plot of the number of instances in which multiple prescriptions were dispensed to the same individual on the same day (> 1), and the proportion of these events which were assigned to simultaneous treatment and advance pick up from our decision tree process.

A summary of processing of the prescription data is shown in Supplementary Figure **11**

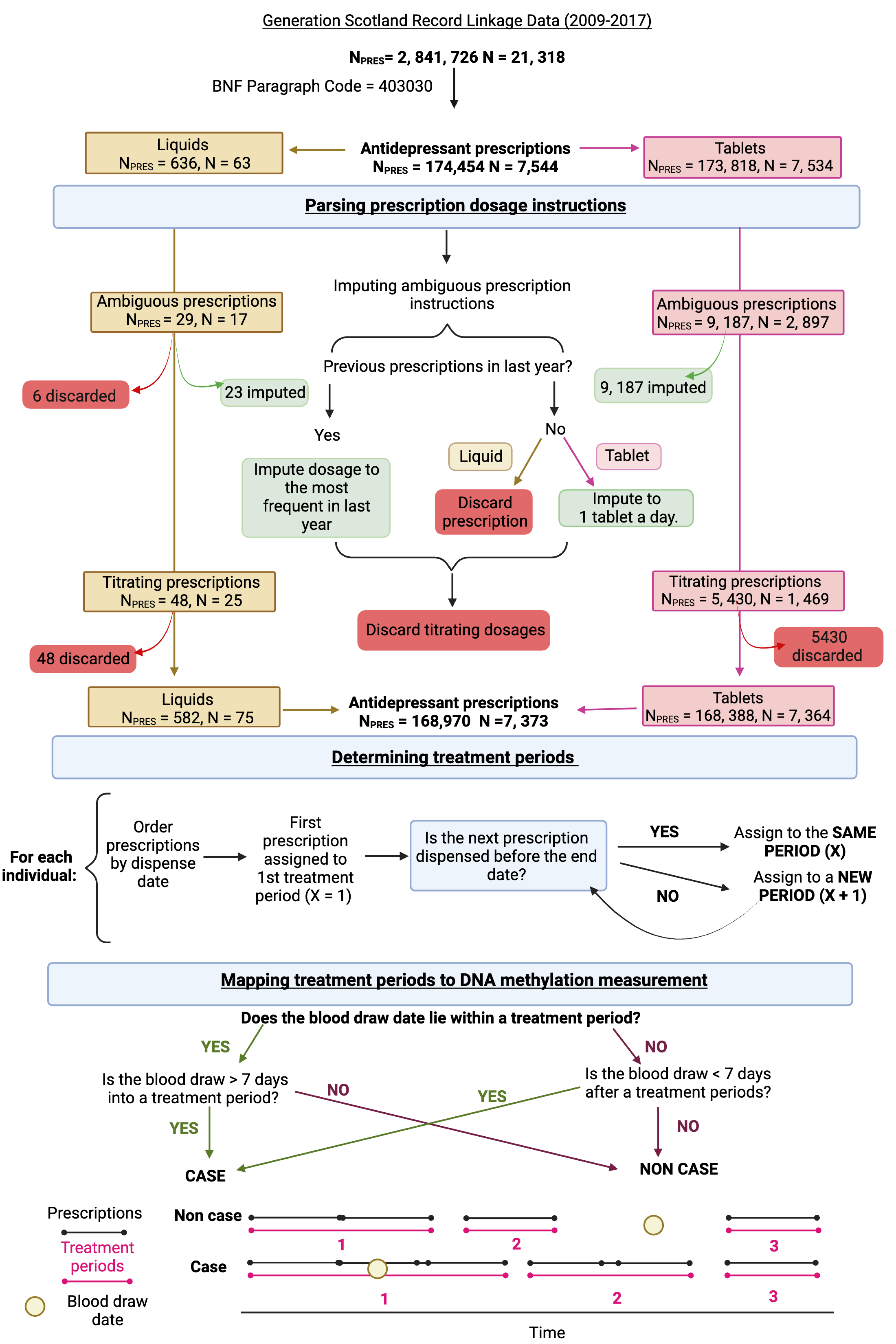

**Figure** **11:** Flow chart demonstrating the processing of prescription data, the derivation of antidepressant treatment periods and the mapping of treatment periods to the DNA methylation measurement.

##### Treatment periods

For each prescription, the prescription duration ($PD$) would be calculated as the fraction of the total dosage by the daily dosage (imputed from the prescription instructions). This is then extended by 10% to account for non-perfect adherance to medication.

$$PD=1.1\left( \frac{Dose_{Total}}{Dose_{Daily}} \right) \text{(1)}$$

The expected end date ($E\left( ED \right)$) of a prescription is then calculated by adding the estimated prescription duration ($PD$) onto the Dispense Date ($DD$) of the prescription, assuming individuals take the first dosage on the day of dispensation.

$$E\left( ED \right)=DD+PD \text{(2)}$$

Each individual had all their antidepressant prescriptions assigned to a treatment ‘period’. The first prescription on record ($Pres_{1}$), was assigned to Period 1 ($Per_{1}$). For the remaining $n$ prescriptions, $Pres_{2}$ to $Pres_{n}$, the period assigned to each prescription was determined by the following criteria:

If the prescription’s dispense date was earlier than the expected end date of the previous prescription ($DD_{Pres_{n}}\leq E\left( ED_{n-1} \right)$), then it would be assigned to the same period as the previous prescription.

$$Period\left( Pres_{n} \right)=Period\left( Pres_{n-1} \right) \text{(3)}$$

The $E\left( ED \right)_{n}$ would then be calculated again according to Equation $\left( 2 \right)$.

Otherwise, in instances where the dispense date is after the expected end date of the previous prescription ($DD_{P_{n}}>E\left( ED_{n-1} \right)$), then it would be assigned to a new period of treatment.

$$Period\left( Pres_{n} \right)=Period\left( Pres_{n-1}+1 \right) \text{(4)}$$

Each treatment period is defined as starting at the dispense date of the first prescription within the treatment period and ending at the expected end date of the last prescription within the period.

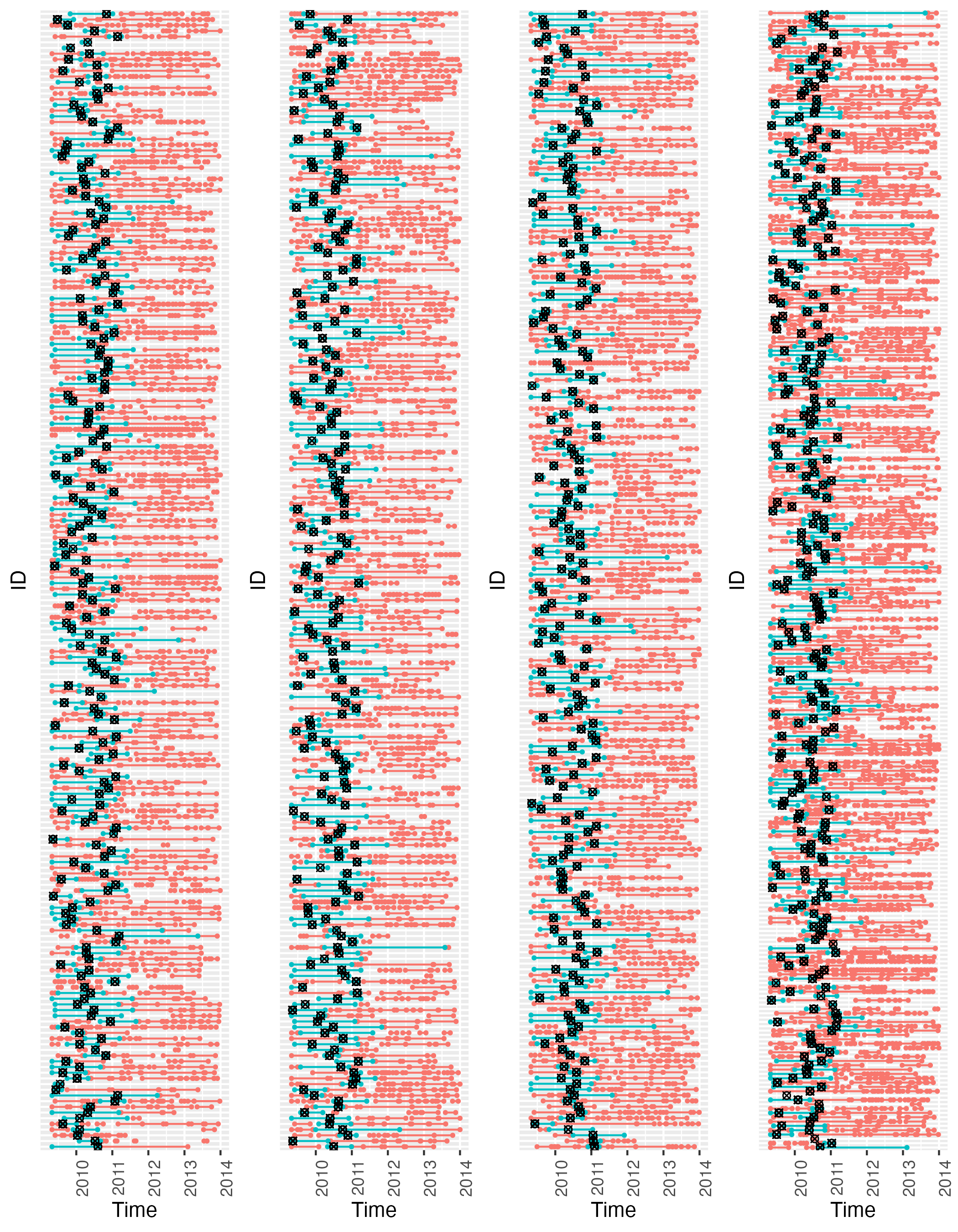

**Figure** **12:** Antidepressant treatment periods for all antidepressant-exposed individuals (n = 861) in the prescription-derived phenotype. Antidepressant exposure is defined as those > 7 days into or < 7 days out of an antidepressant treatment period at the time of the blood draw. The time point of the blood-draw appointment is denoted by the black symbol, the treatment periods are colored depending on whether it encompasses the blood draw appointment (blue) or not (red).

#### Self-report phenotypes

Self-reported antidepressant exposure was assessed using two different questionnaires. The first version (n = 9, 924) enabled participants to free text any medications they were taking. The second version (n = 13, 675) had the question “Are you regularly taking any of the following medications” with a yes/no checkbox next to a “Antidepressants” option. Classification of antidepressant exposure was done using both; those who named an antidepressant medication from the British National Forumulary Chapter “040303” (which roughly equates to Anatomical Therapeutic Chemical (ATC) subclass ‘NO6A’) and those who checked ‘yes’ for “Antidepressants” (N_exposed_ = 1, 508) were classed as antidepressant-exposed. Those who did not name any antidepressants or checked ‘no’ were classed as antidepressant-unexposed (N_unexposed_ = 15, 028). The medications which were selected as antidepressants in the 1st version of the self-report medications questionnaire included tricyclic antidepressants (Amitriptyline, Amoxapine, Clomipramine, Dosulepin, Doxepin, Imipramine, Lofepramine, Maprotiline, Mianserin, Nortriptyline, Trazodone, Trimipramine), monoamine-oxidase inhibitors (Isocarboxazid, Moclobemide, Phenelzine, Tranylcypromine), selective serotonin reuptake inhibitors (Citalopram, Duloxetine, Escitalopram, Fluoxetine, Fluvoxamine Maleate, Paroxetine, Sertraline), and other antidepressants (Agomelatine, Duloxetine, Flupentixol, Mirtazapine, Nefazodone, Oxitriptan, Reboxetine, Tryptophan, Venlafaxine, Vortioxetine).

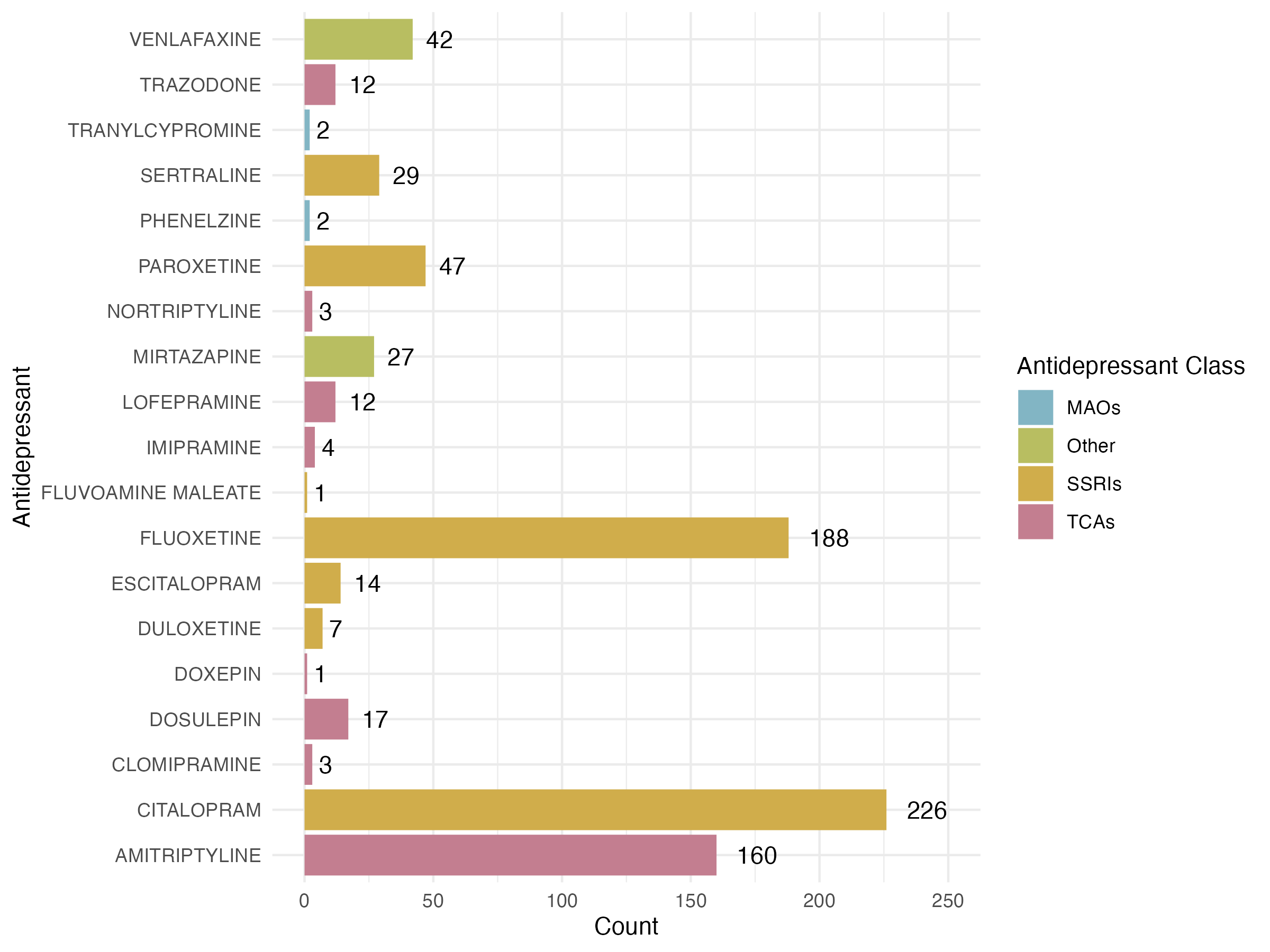

**Figure** **13:** Self-reported antidepressant medications named by participants using free-text medication reporting (n = 9127, n_exposed_ = 797).

#### Summary

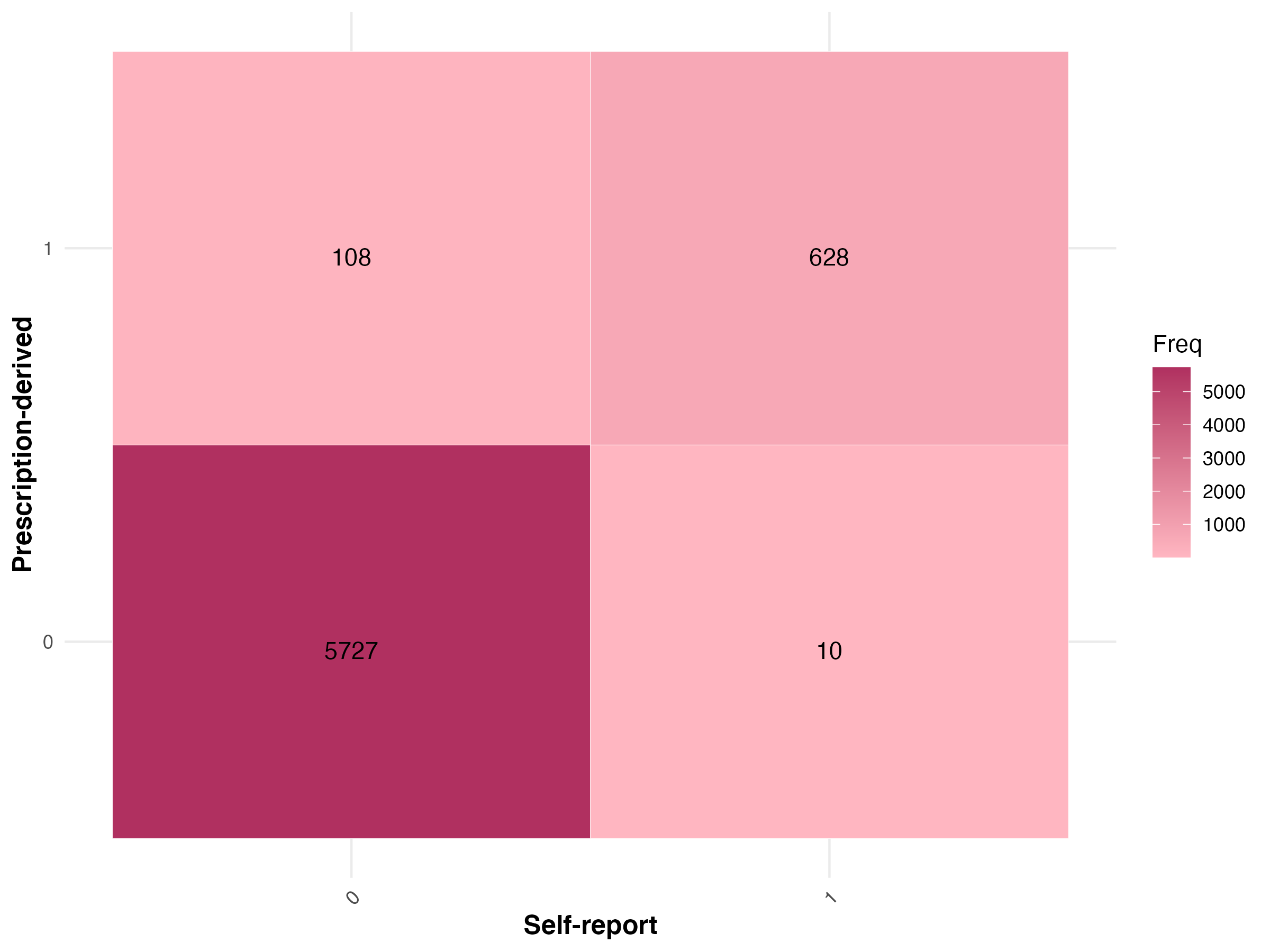

**Figure 14:** Comparison of self-report and prescription-derived measures of antidepressant exposure in those present in both samples (N = 6,473). Coding: 0 = Antidepressant-unexposed, 1 = Antidepressant-exposed.

### Major Depressive Disorder

Major Depressive Disorder (MDD) was confirmed using the Structured Clinical Interview for DSM-IV Non-Patient Version (SCID)^14^. Those who answered ’yes’ to an initial two screening questions, completed a more extensive interview about the nature of the MDD (i.e a current or historical episode of MDD, the age of onset and the number of episodes). However, for this analysis, the phenotype was binary-coded: participants who answered no for the initial screen, or those who did not meet the MDD criteria following the extensive interview were assigned as controls, and those who answered yes and met the criteria for either single episode or recurrent MDD were assigned as cases. Participants who fulfilled criteria for bipolar disorder in the SCID assessment were discarded (n = 75). All interviewers received multiple group training sessions in the administration of the SCID throughout the study and were blinded to the diagnostic status of any relatives in the study. Furthermore, the SCID phenotype within GS was found to have a high inter rater-reliability for the identification of MDD cases^15^.

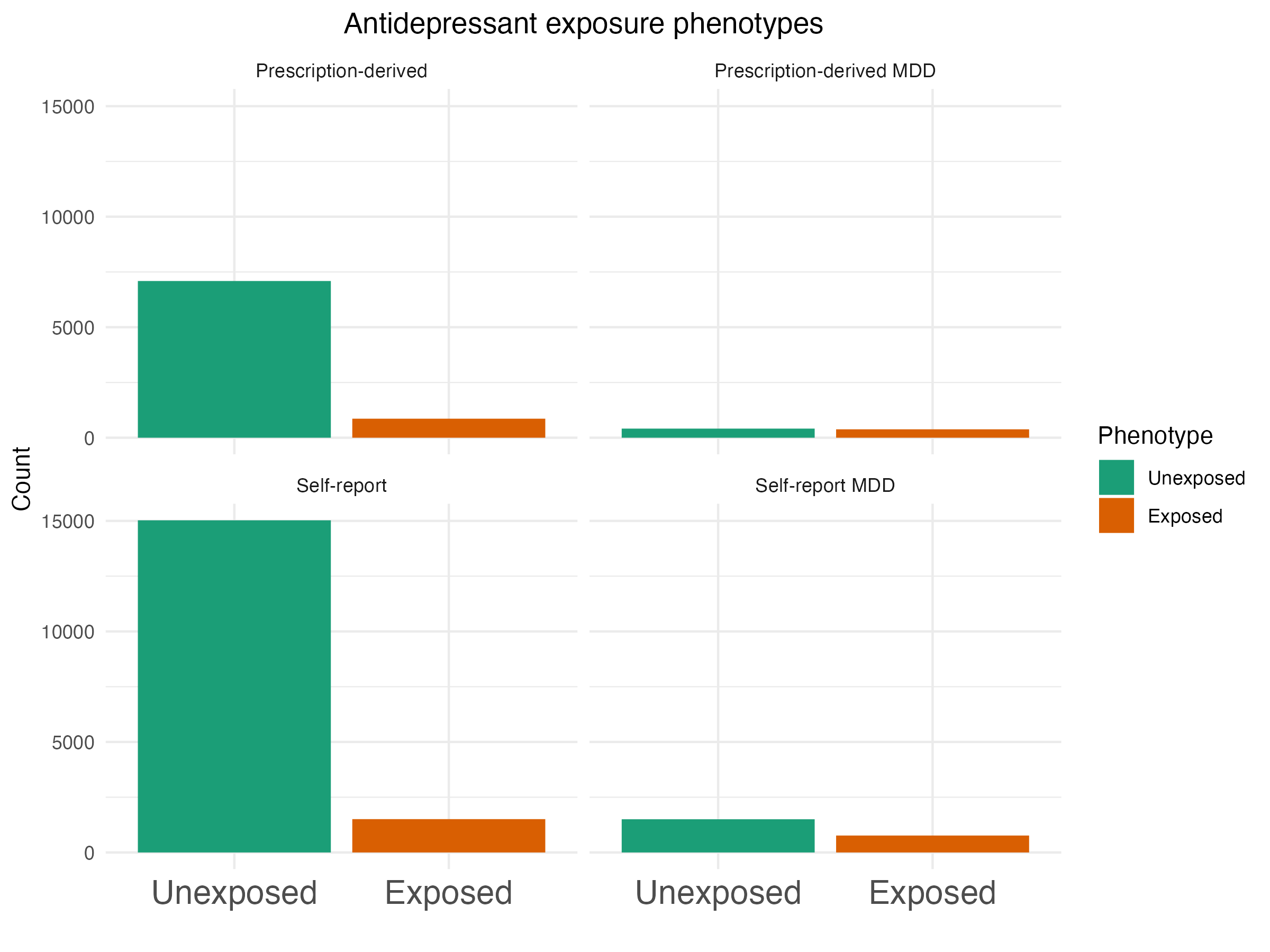

**Figure** **15:** Antidepressant exposure phenotypes used in the MWAS analysis. Left Panel: Sample used in the discovery MWAS analysis. Right Panel, Samples filtered to MDD cases only, used to investigate the potential confounding effect of MDD status.

### External Cohorts

The association between antidepressant exposure and antidepressant exposure MPS was assessed using generalised linear mixed models, generalised linear models and generalised estimation equations, depending on the cohort’s population structure (i.e., twin studies vs. unrelated participants) and DNAm pre-processing. In depth information about each cohort, the DNAm preprocessing, the calculation of the MPS and the association model (antidepressant-exposure ~ MPS) is detailed below, and summarised in Supplementary Tables 7-9.

#### Finnish Twin Cohort (FTC)

Protocol papers:

Older Finnish Cohort: <https://doi.org/10.1017/thg.2019.54>^16^

FinnTwin12: <https://doi.org/10.1017/thg.2019.83>^17^

FinnTwin16: <https://doi.org/10.1017/thg.2019.106>^18^

##### Cohort Description

The Finnish Twin Cohort comprises of three longitudinal sub cohorts: The Older Finnish Twin Cohort^16^ (Old), established in the 1970s, and the FinnTwin12 (FT12)^17^ and FinnTwin16 (FT16)^18^ cohorts established in the 1990s to identify genetic and environmental precursors of various health-related behaviours and diseases. The Old cohort consists of Finnish same-sex monozygotic and dizygotic twin pairs born before 1958 (n = 13, 888 pairs) with extensive data collection including behavioural and lifestyle traits over multiple waves in 1975, 1981, 1990 and 2011. The younger cohorts, FT12 and FT16 are longitudinal studies of five consecutive birth cohorts (born in 1983-1987, n = 2, 700 pairs and 1975-1979, n = 2, 800 pairs, respectively) of Finnish monozygotic and dizygotic twin pairs. Participants have completed multiple surveys on behavioural and lifestyle traits at five rounds of follow up beginning in adolescence through to adulthood (FT16 at age 16, 17, 18.5, 24, 34; FT12 at age 12, 14, 17.5, 22, 35). In addition to the 4-5 questionnaire waves, the twins have participated in several intensive on-site studies including interviews, clinical measures and tests, and blood sampling. FT16 intensive study in 2001-2006 included pairs discordant and concordant for alcohol use. These pairs were interviewed and sampled at the mean age of 26, with 599 twins providing DNA^19^. Wave 4 of FT12 included intensive study for randomly selected pairs during 2006-2009, with 1260 twins completing the interviews and clinical protocol, and providing DNA at the mean age of 222. Old cohort twin pairs discordant and concordant for blood pressure were included in a study on epigenetics of essential hypertension (EH-Epi) in 2012-2015^20^. Altogether 428 twins at the mean age of 62 participated in the clinical protocol and provided DNA samples.

**Ethical Approval**: Participants were given information on the study procedures and of freedom to participate or to decline at any point in both oral and written form. Informed consent was obtained upon the contact with the study subjects before new questionnaire information was collected, and when clinical investigations were undertaken with sampling of biological material. Ethics approvals have been granted for multiple studies concerning the FTC twins by the ethics committees of Helsinki University Central Hospital (113/E3/2001, 249/E5/2001, 346/E0/05, 270/13/03/01/2008, and 154/13/03/00/2011) with the last one on the transfer of biological samples to the THL Biobank in 2018 (HU51179912017).

##### Antidepressant exposure phenotype

In the younger cohorts, FT12 and FT16, antidepressant exposure was recorded using the psychiatric Semi-Structured Assessment for Genetics of Alcohol (SSAGA) psychiatric interviews, which included question on current medication use (“Have you used antidepressants within the last 30 days?”) in a selected subsample of participants, as described above. Antidepressant exposure in the Old cohort was ascertained in a subsample of 447 twins participating in the EH-Epi study using on-site interview, which asked, “What medications are you currently using?” For all the twin cohorts (FT12, FT16 and Old) the medications were coded by Anatomical Therapeutic Chemical (ATC codes), and subclasses of N06A were considered antidepressants. Those who named any antidepressant medication were defined as antidepressant-exposed and were defined as those who did not name a single antidepressant medication. In total, there were 84 exposed (41 FT12, 18 FT16 and 25 Old cohort) and 1594 unexposed (805 FT12, 478 FT16 and 311 Old cohort) with good quality DNAm data generated from the blood samples collected at the same visit with the interview on AD use.

##### DNAm preprocessing

DNAm was profiled from whole blood samples using the Infinium HumanMethylation450K (‘450K array’) or EPIC (‘EPIC array’) BeadChip Kit (Illumina, San Diego, CA, USA). Altogether 336 Old cohort (Average Age = 62.1), 764 FT12 (Average Age = 22.4), and 215 FT16 (Average Age = 26.1) cohort individuals were measured on the 450K array, and 82 FT12 (Average age = 22.3) and 281 FT16 (Average age = 26.3) cohort individuals were measured on the EPIC array. The following quality control pipeline was used for both the 450K and EPIC DNAm data. The DNAm data were pre-processed in the R package *meffil*^7^. Bad quality samples were excluded based on the following criteria: i) sex mismatch, ii) median methylation vs. unmethylated signal > 3 standard deviations (SD), iii) failed control probe metrics and if > 20% of probes per sample had iv) detection p value > 0.05 and v) bead number < 3. To remove technical variation between the samples, functional normalisation including the control probe principal components was performed, followed by bad quality probe removal: i) probes with detection p value > 0.05 in more than 20% samples, ii) bead number < 3 in more than 20% samples, iii) sex chromosome probes and iv) cross-reactive and ambiguously mapped probes as well as probes on polymorphic CpGs^9^. Beta Mixture quantile (BMIQ) normalisation implemented in the R package *wateRmelon*^4^ was then performed to adjust for type 2 probe bias. The data pre-processing resulted in 390,304 and 765,385 probes in 450K (n = 1315) and EPIC data (n = 363), respectively. Cell-type fractions were estimated using *EpiDISH* R package^22^, running Robust Partial Correlations with a reference for 7 main immune cell subtypes: neutrophils, basophils, monocytes, B-cells, NK-cells, CD4+ and CD8+ T-cells^23^. Lymphocyte cell proportions were then calculated as the aggregate of the estimated CD4+ T, CD8+ T, NK and B cell proportions. Methylation beta values were transformed to M-values using *‘logit2()’* function in the *minfi*^5^ R package. The M-values were then standardised using Z-score normalisation using the *scale()* function in R.

##### Methylation Profile Score

Following the quality control protocol, there were 183 CpGs and 211 CpGs present on the 450K (all the 3 cohorts) and EPIC array (FT12 & FT16) respectively, out of the 212 CpGs which make up the methylation profile score (MPS). There was no missingness in the CpGs deriving the MPS from either array. The MPS for each individual was derived as a weighted sum of the 183 or 211 available CpGs, depending on which array DNAm was profiled.

##### Antidepressant exposure ~ MPS association model

The association between antidepressant exposure and the antidepressant exposure MPS was tested under a generalised estimation equation (GEE) model, using the *‘glmgee()’* function from the *glmtoolbox* R package^24^, with antidepressant exposure as the outcome. The model included the following predictors: antidepressant exposure MPS, age at blood sampling, sex, the estimated monocyte cell proportions, estimated lymphocyte (aggregate of CD4T, CD8T, B cells and NK cells) cell proportions and M values at the AHRR probe (‘cg05575921’) to proxy for smoking status. The following settings were used: family=binomial, 50 maximum number of iterations, and the ‘exchangeable’ option to account for correlation structure within families and within persons.

```
assoc_mod <- glmgee(as.factor(antidep)~ scale(AD_MRS) + scale(age) + scale(Mono) +
                 scale(lymphocytes) + scale(cg05575921) + as.factor(sex_coded),
                 data = MRS_covs_pheno,
                 id = familyID,
                 family = binomial,
                 corstr = "Exchangeable")
```

##### Funding and Acknowledgment

Phenotype and genotype data collection in FT12 and FT16 studies of the Finnish twin cohort has been supported by the Wellcome Trust Sanger Institute, the Broad Institute, ENGAGE – European Network for Genetic and Genomic Epidemiology, FP7-HEALTH-F4-2007, grant agreement number 201413, National Institute of Alcohol Abuse and Alcoholism (grants AA-12502, AA-00145, and AA-09203 to R J Rose; AA15416 and K02AA018755 to D M Dick; R01AA015416 to J Salvatore) and the Academy of Finland (grants 100499, 205585, 118555, 141054, 264146, 308248, 265240, 263278 to J Kaprio, 328685, 307339, 297908 and 251316 to M Ollikainen, and Centre of Excellence in Complex Disease Genetics grants 312073, 336823, and 352792 to J Kaprio), NIH/NHLBI (grant HL104125 to X Wang), and Sigrid Juselius Foundation to M Ollikainen and J Kaprio.

#### Study of Health in Pomerania (SHIP-Trend)

Protocol paper:<https://doi.org/10.1093/ije/dyac034>^25^

##### Cohort Description

The Study of Health in Pomerania (SHIP) is a population-based cohort (N ~ 12,728, Trend = 4,420 and Start = 4,308, Next ~ 4,000) with the aims of assessing the prevalence and risk factors underlying several clinical and subclinical disorders within Pomerania, Western Germany^25,26^. For this analysis, data from the SHIP-Trend cohort was used. Comprehensive data collection of this cohort, including in-depth interviews, imaging, and biological sampling for 4,420 subjects was conducted between September 2008 and September 2012.

**Ethical Approval** Participants provided written informed consent before any assessment and/or sampling took place. The Ethics Committee of the University Medicine Greifswald, Germany provided ethical approval for the study (BB 39/08).

##### Antidepressant exposure phenotype

All participants were asked to bring any medications they were currently taking (in the last 7 days) to their in-clinic interview. Medications were coded using Anatomical Therapeutic Chemical (ATC) codes, subclasses of N06A were considered antidepressants. Those who brought any antidepressant medication were defined as antidepressant-exposed, and those were defined to those who did not bring any antidepressant medication were defined as antidepressant-unexposed. In total, there were 21 exposed and 474 unexposed with quality DNAm data.

##### DNAm preprocessing

DNAm was profiled from blood samples at baseline for a subsample of SHIP-Trend participants, randomly selected based on the availability of multi-OMICS data, using the Illumina HumanMethylation EPIC BeadChip array (n=508). Samples excluded type II diabetes and were enriched for prevalent myocardial infarction. The samples were taken between 07:00 AM and 04:00 PM. Serum aliquots were prepared for immediate analysis and for storage at -80 °C in the Integrated Research Biobank (Liconic, Liechtenstein) and the DNA samples were processed at the Helmholtz Zentrum München. The CPACOR workflow was followed for the preparation and normalization of the array data using the software package R (www.r-project.org), with the array idat files being processed using the *minfi* R package^5^. Probes that had a detection p-value above background (sum of per-array methylated and unmethylated intensity values, based p-value ≥ 1x10-16) were set to missing. Methylation beta values were calculated as proportion of methylated intensity value on the sum of methylated+unmethylated+100 intensities. During steps such as bisulfite conversion, hybridization and extension, arrays were removed following observation of technical problems (±4 SD outside control probe intensity mean). Additionally arrays with a mismatch of recorded sex and methylation-predicted sex were excluded. Additionally, arrays with a call rate < 5% were removed from the analysis. A total of 495 samples with methylation data on 865,859 sites was available for subsequent analyses following quality control. Methylation β values were transformed into M-values by first setting values of zero to half the value of the smallest non-zero β of the CpG site over all participants, and then using Mi=log2(βi/(1-βi)). The M-values were then standardised using Z-score normalisation using the *‘scale()’* function in R.

##### Methylation Profile Score

After adhering to the quality control protocols, all 212 CpGs were available for the calculation of the MPS, which was derived as a weighted sum of the CpGs. There were 145 individuals with data available for all 212 CpGs, with others having varying degrees of missingness in the data. This was mainly confined to 3 CpGs (cg13569486, cg26192826 and cg15897613). The MPS for each individual was derived as a weighted sum of the CpGs. In the case that an individual had a missing value for a CpG(s), the probe(s) were excluded from the MPS (i.e., given a weight of 0).

##### Antidepressant exposure ~ MPS association model

The association between the antidepressant exposure and antidepressant exposure MPS was tested using a generalised linear mixed effects model with a logistic link function, using the *‘glmer()’* function from the *lme4* R package^27^, with antidepressant exposure as the outcome. The model included the following predictors with a fixed effect: antidepressant exposure MPS, age at blood sampling, sex, white blood cell count, percentage of monocytes and lymphocytes, M values at the AHRR probe (‘cg05575921’) to proxy for smoking status, and the top 3 principal components of the control probe intensities (obtained by the CPACOR workflow). Each of those variables (except sex) were standardised using Z-score normalisation. Additionally, array processing batch was included as a random effect.

```
Generalized linear mixed model fit by maximum likelihood (Laplace Approximation) ['glmerMod']
Family: binomial  ( logit )
Formula: as.factor(antidep) ~ scale(AD_MRS) + scale(AGE_SHIP_T0) + scale(mo_pct_e) + scale(ly_pct_e) + scale(AHRR) + as.factor(SEX_SHIP_T0) + scale(PC1_cp) + scale(PC2_cp) + scale(PC3_cp)  + (1 | Batch) + scale(wbc)
```

##### Funding and Acknowledgement

SHIP is part of the Community Medicine Research net of the University of Greifswald which is funded by the Federal Ministry of Education and Research (01ZZ9603, 01ZZ0103, and 01ZZ0403), the Ministry of Cultural Affairs and the Social Ministry of the Federal State of Mecklenburg-West Pomerania. DNA methylation data have been supported by the DZHK (grant 81X3400104). The University of Greifswald is a member of the Caché Campus program of the InterSystems GmbH.

#### FOR2107

##### Study Population

Protocol paper: <https://doi.org/10.1007/s00406-018-0943-x>^28^

FOR2107 is a research programme including both a human cohort (target n = 2500) and animal models, which broadly aims to establish the neurobiological mechanisms of genetic and environmental risk factors for affective disorders^28^. The human cohort consists of individuals from various groups, those diagnosed with MDD, those diagnosed with bipolar disorder, those diagnosed with schizophrenia, those diagnosed with schizoaffective disorder, and healthy controls (HC) with and without genetic/environmental risk factors. Participants undergo an in-person extensive deep-phenotyping (MR imaging, clinical course, neuropsychology, personality, risk/protective factors, biomaterial: blood, stool, urine, hair, saliva) at baseline and again at a follow-up appointment two years later. Baseline data collection started in November 2014 and is ongoing.

**Ethical Approval** Participants provided written consent before any assessment and/or sampling took place. The ethics committees of the Medical Faculties, University of Marburg (AZ: 07/14) and University of Münster (2014-422-b-S) provided ethical approval for the study.

##### Antidepressant exposure phenotype

During the in-person assessment participants were asked about their current medication. The assignment of drugs to the antidepressant treatment group largely aligns with the ATC coding and BNF coding (Supplementary Table 1). Individuals diagnosed with bipolar or schizophrenia, or schizoaffective disorder were excluded from the analysis. Those who named any single antidepressant medication were defined as antidepressant-exposed and those who did not name a single antidepressant medication nor any single neuroleptic medication were defined as antidepressant-unexposed, based on the described classification. In total, there were 165 exposed (all having a lifetime diagnosis of MDD) and 493 unexposed (thereof, 167 with a lifetime diagnosis of MDD and 326 HC) with high-quality DNAm data.

##### DNAm preprocessing

DNAm profiling and preprocessing were conducted within the larger FOR2107 DNAm sample (n=881), of which the sample eligible for the current study constitutes a subset. DNAm was profiled from peripheral whole blood samples using the Illumina Infinium MethylationEPIC BeadChip v1.0. Quality control was performed using the *minfi*^5^ and *ewastools*^29^ R packages. Samples which had a call rate < 98 %, sex mismatch, or mismatch in SNP fingerprints and those with technical issues detected from the Illumina control probes were excluded from the analysis. After stratified quantile normalisation, methylation probes which had a call rate < 98%, SNPs inside the probe body, a non-CpG type or a non-autosomal genomic position were excluded. Cross-reactive or polymorphic probes with an allele frequency of >5 % in the European population (n = 52, 541) were excluded based on procedure outlined by McCartney et al (2016)^10^. The sample was further filtered for the availability of high-quality genome-wide genotyping data (Illumina Infinium PyschArray-24 BeadChip) to examine relatedness within the sample, and relatives with pi-hat > 0.125 were excluded. After filtering the sample for the eligibility criteria of the antidepressant exposure phenotype defined above, 764, 556 CpGs and 658 individuals remained for analysis. M values were extracted for downstream analysis using the *‘getM()’* function implemented in *minfi* and were then standardised using Z-score normalisation using the *‘scale()’* function in R. Cell proportions (CD4T, CD8T, NK cells, B cells, Granulocytes and Monocytes) were estimated using the Houseman approach as implemented in *minfi*^5^. Lymphocyte cell proportions were then calculated as the sum of the estimated CD4T, CD8T, NK and B cell proportions.

##### Methylation Profile Score

After adhering to the quality control protocols, all 212 CpGs were available for the calculation of the MPS, which was derived as a weighted sum of the CpGs. There was no missingness in any of the CpGs included in the score.

##### Antidepressant exposure ~ MPS association model

The association between the antidepressant exposure and antidepressant exposure MPS was tested using a generalised linear model with a logistic link function, using the *‘glm()’* function from the *stats* R package^30^, with antidepressant exposure as the outcome. The model included the following predictors with a fixed effect: antidepressant exposure MPS, age at blood sampling, sex, estimated lymphocyte cell proportions (aggregate of CD4T, CD8T, B cells and NK cells), estimated monocyte cell proportions, M values at the AHRR probe (‘cg05575921’) to proxy for smoking status, and the top 10 genetic principal components.

```
glm(formula = as.factor(antidep) ~ scale(AD_MRS) + scale(age) +
    scale(Mono) + scale(lymphocytes) + scale(cg05575921) + as.factor(sex_coded) +
    scale(C1) + scale(C2) + scale(C3) + scale(C4) + scale(C5) +
    scale(C6) + scale(C7) + scale(C8) + scale(C9) + scale(C10),
    family = "binomial", data = MRS_covs_pheno)
```

##### Funding and Acknowledgement

The German multicenter consortium “Neurobiology of Affective Disorders. A translational perspective on brain structure and function” is funded by the German Research Foundation (Research Unit FOR2107). Principal investigators are Tilo Kircher (speaker FOR2107, DFG grant numbers KI588/14-1, KI588/14-2, KI588/20-1, KI588/22-1, KI 588/15-1, KI 588/17-1), Udo Dannlowski (co-speaker FOR2107; DA1151/5-1, DA1151/5-2, DA1151/6-1), Axel Krug (KR3822/5-1, KR3822/7-2), Igor Nenadic (NE2254/1-2, NE2254/2-1, NE2254/3-1, NE2254/4-1), Carsten Konrad (KO4291/3-1), Marcella Rietschel (RI 908/11-1, RI 908/11-2), Markus Nöthen (NO 246/10-1, NO 246/10-2), Stephanie Witt (WI 3439/3-1, WI 3439/3-2). Tilo Kircher received unrestricted educational grants from Servier, Janssen, Recordati, Aristo, Otsuka, neuraxpharm. We are deeply indebted to all study participants and staff. A list of acknowledgments can be found here: www.for2107.de/acknowledgements.

#### Netherlands Twin Register

Protocol paper: <https://doi.org/10.1017/thg.2019.93>^31^

##### Study population

The Netherlands Twin Register is a population-based cohort of over 200,000 people from across the Netherlands. It consists of twin-families, i.e. twins, their parents, spouses and siblings aged between 0 and 99 years at recruitment, and started around 1987 with new–born twins and adolescent and adult twins. Full details have been reported previously^31^. DNA was collected from buccal cells and whole blood as part of multiple projects. For the current paper, we analysed DNA methylation measured in whole blood collected in the NTR-Biobank study^34^. Good quality whole blood DNA methylation data were available for 3087 samples from 3055 individuals, including monozygotic and dizygotic twins, parents of twins, siblings of twins and spouses of twins. For 32 individuals, longitudinal methylation data were available (two time points, mean range=5.2 year). The current analysis included individuals with information on antidepressant medication use and covariates (N = 3, 004 samples). **Ethical Approval** Informed consent was obtained from all participants. The study was approved by the Central Ethics Committee on Research Involving Human Subjects of the VU university Medical Centre, Amsterdam, an Institutional Review Board certified by the U.S Office of Human Research Protections (IRB number IRB00002991 under Federal-wide Assurance-FWA00017598; IRB/institute codes, NTR 03-180).

##### Antidepressant exposure phenotype

At blood draw, participants were asked about all current medication use and for all medicines the dose, brand and chemical names were recorded directly from the medication packaging. Following the Anatomical Therapeutic Chemical (ATC) classification system, subclasses of N06A were considered anti-depressants. Antidepressant medication use was coded as 1 (antidepressant-exposed) / 0 (antidepressant-unexposed). There were 87 recorded exposed and 2,917 unexposed with quality DNAm data.

##### DNAm preprocessing

DNA methylation was assessed with the Infinium HumanMethylation450 BeadChip Kit (Illumina, San Diego, CA, USA) by the Human Genotyping facility (HugeF) of ErasmusMC, the Netherlands (<http://www.glimdna.org/>) as part of the Biobank-based Integrative Omics Study (BIOS) consortium^35^. DNA methylation measurements have been described previously^35^. Genomic DNA (500ng) from whole blood was bisulfite treated using the Zymo EZ DNA Methylation kit (Zymo Research Corp, Irvine, CA, USA), and 4 µl of bisulfite-converted DNA was measured on the Illumina 450k array following the manufacturer’s protocol. A number of sample- and probe-level quality checks and sample identity checks were performed, as described in detail previously^34^. In short, sample-level QC was performed using R pacakge, *‘MethylAid’*^36^. Probes were set to missing in a sample if they had an intensity value of exactly zero, or a detection p > .01, or a bead count of<3. After these steps, probes that failed based on the above criteria in >5% of the samples were excluded from all samples (only probes with a success rate ≥ 0.95 were retained). The following probes were also removed: sex chromosomes, probes with a single nucleotide polymorphism (SNP) within the CpG site (at the C or G position) irrespective of minor allele frequency in the Genome of the Netherlands (GoNL) population, irrespective of minor allele frequency^37^, and ambiguous mapping probes reported by Chen et al with an overlap of at least 47 bases per probe^21^. The methylation data were normalized with functional normalization^38^.

##### Methylation Profile Score

After adhering to quality control protocols, all 212 CpGs were available for the calculation of the MPS, which was derived as a weighted sum of the CpGs. The rate of missingness per CpG ranged from 0% to 3.2% (average 0.4%). 71 CpGs had no missing values. In the case that an individual had a missing value for a CpG(s), the probe(s) were excluded from the MPS (i.e., given a weight of 0).

##### Antidepressant exposure ~ MPS association model

The association between antidepressant exposure and the MPS was assessed using a generalised linear mixed model, using the *‘glmer()’* function from the *lme4* R package^27^, with antidepressant exposure as the outcome. The model included the following predictors, antidepressant exposure MPS, sex, age at blood sampling, percentages of monocytes, eosinophils and neutrophils, HM450K array row, 96-wells bisulfite sample plate (dummy-coding), M values at the AHRR probe (‘cg05575921’) to proxy for smoking status and family ID as a random effect. The model was specified with the logit link function and 20 quadrature points in the adaptive Gaussian quadrature approximation for integrating over the random effect. The optimisation algorithm used was the Bound Optimisation by Quadratic Approximation (BOBYQA) with a maximum of 100,000 function evaluations.

assoc_mod_glmr2 <- glmer(as.factor(antidep) ~ scale(AD_MRS) + scale(age) + as.factor(sex) + scale(Mono_Perc) + scale(Eos_Perc) + scale(Neut_Perc) + as.numeric(Array_rownum) + as.factor(Sample_Plate_collapsed_fix) + scale(cg05575921) + (1|familynumber), data=MRS_covs_pheno_ahhr, family=binomial(link = "logit"), nAGQ = 20,
 control = glmerControl(optimizer = "bobyqa", optCtrl = list(maxfun = 100000)))

##### Funding and Acknowledgements

We warmly thank all twin families of the Netherlands Twin Register who make this research possible. This work was supported by the Royal Dutch Academy for Arts and Science (KNAW) Academy Professor Award (PAH/6635) to DIB; the Netherlands Organization for Scientific Research (NWO 480-15-001/674) and Biobanking and Biomolecular Resources Research Infrastructure (BBMRI-NL: 184.021.007; 184.033.111).

#### Munich Antidepressant Response Study / Unipolar Depression Study

MARS protocol paper: <https://doi.org/10.1016/j.jpsychires.2008.05.002>^39^

UniDep protocol paper: <https://doi.org/10.1093/hmg/ddl166>^40^

##### Study population(s)

The Munich Antidepressant Response Signature (MARS) project^39^ is a naturalistic longitudinal clinical study providing a sample of adult Caucasian inpatients (aged between 18 and 75 years) admitted to psychiatric hospitals in an acute episode of major depression (according to ICD-10) in southern Germany. Data was collected between 1995 and 2005. Further characterisation of the cohort and information on study protocol has been described elsewhere^39^.

**Ethical Approval**: The study was approved by the local Ethics Committee of the Ludwig Maximilians University, Munich, Germany, and carried out in accordance with the latest version of the Declaration of Helsinki. All participants provided written consent after the study protocol and potential risks were explained.

The Unipolar Depression study (UniDep)^40^ is a cross-sectional case-control study in Germany. It consists of German in and outpatients (n = 1000, 67.4% female) with recurrent MDD from the Max-Planck-Institute of Psychiatry in Munich and psychiatric hospitals in Augsburg and Ingolstadt (located near Munich), with each hospital contributing a third of the patients. Patients were diagnosed according to the DSM-IV using the schedule for Clinical Assessment in Neuropsychiatry. Only Caucasian patients over 18 years of age with at least two moderate/severe depressive episodes were included. Those with the presence of manic or hypomanic episodes, mood incongruent psychotic symptoms, the presence of a lifetime diagnosis of intravenous drug abuse, and depressive symptoms which are secondary to a substance abuse disorder or due to medical illness or medication were excluded from the study. The mean age was 49.35 (±14.09 years). A control sample (n = 1029) was randomly selected from a Munich-based community sample, screened for the presence of anxiety or affective disorders using the Composite International Diagnostic Screener (CIDI), and matched for ethnicity, age (to 5-year intervals) and sex to those in the cases sample. Baseline data was collected between 2002 and 2004. Further characterisation of the cohort is described by Lucae et al (2006)^40^.

**Ethical Approval**: The study was approved by the Ethics committee of the Ludwig Maximillans University in Munich, Germany and written informed consent was obtained from all subjects.

##### Antidepressant exposure phenotype

For the MARS Depression cohort, baseline antidepressant exposure was assessed by asking patients about the antidepressants they had taken before admission to the hospital. During their stay at the hospital, information on the use of antidepressants was taken weekly from the patients clinical record. Assignment of drugs to the antidepressant group largely aligns with the ATC coding and BNF coding (Supplementary Table 1). The antidepressant exposure phenotype was taken from the antidepressant measure closest to the blood draw date per individual within 14 days of the blood draw. The mean/median time between the self-report rating and the blood draw was a day and 0 days respectively.

In the UniDep cohort, which was designed as a case-control study for the identification of genes contributing to unipolar depression, antidepressant exposure was not assessed among patients. Therefore, from the UniDep study only controls who were screened for mental disorders and any psychopharmacological medication use, could be included in the present analysis. Those who named any single antidepressant medication were defined as antidepressant-exposed (MARS only), and those who did either not name an antidepressant medication (MARS) or were recruited as controls (without MDD) were defined as antidepressant-unexposed (MARS + UniDep). In total, there were 135 antidepressant-exposed (MARS) and 177 antidepressant unexposed (35 MARS, 142 UniDep) with high quality DNAm data.

##### DNAm preprocessing

For both cohorts, genomic DNA was extracted from whole blood samples using the Gentra Puregene Genomic DNA and run together on the same arrays. Samples were randomised with respect to case-control status, sex and age. Genomic DNA was bisulfite converted using the Zymo EZ-96 DNA Methylation Kit (Zymo Research) and DNA methylation levels were assessed for > 480, 000 CpG sits using the Infinium HumanMethylation450 BeadChip Kit (Illumina, San Diego, CA, USA). Quality control of DNAm data, including intensity read outs, normalisation, cell type composition estimation, Beta-value and M value calculation was performed using the Bioconductor R package *minfi* (version 1.10.2)^5^. Samples with outlying mean intensities and those with mismatch between DNAm-predicted and reported sex were excluded from the analysis. Probes present on the X and Y chromosomes and those with non-specific binding^21^ were removed. Additionally, probes with a detection P-value > 0.01 in > 50% of the samples and those with a SNP located either at the interval at which the probe was design to hybridise or within 10bp from a SNP with a minor allele frequency > 0.01 (as reported in the 1000 Genomes Project)^41^ were removed. Following these steps there was ~ 425, 000 CpGs available for further analysis. The data were then normalized with functional normalisation, using the *‘FunNorm()’* function, an extension of quantile normalisation including in *minfi*. Visual inspection of the methylation principal components analysis plots was conducted to identify batch effects using the Bioconductor R package *shinyMethyl* (version 0.99.3)^3^. Identified batch effects (i.e., bisulfite conversion plate and plate position) were removed using the Empirical Bayes’ method *‘ComBat’*^42^. Batch-corrected M-values after *ComBat* were used for all further statistical analyses.

##### Methylation Profile Score

After adhering to the quality control protocols, 210 CpGs were available for the calculation of the MPS, which was derived as a weighted sum of the CpGs. There was no missingness in any of the CpGs included in the score.

##### Antidepressant exposure ~ MPS association model

The association between the antidepressant exposure and antidepressant exposure MPS was tested using a generalised linear model with a logistic link function, using the *‘glm()’* function from the *stats* R package^30^, with antidepressant exposure as the outcome. The model included the following predictors with a fixed effect: antidepressant exposure MPS, age at blood sampling, sex, estimated lymphocyte cell proportions (aggregate of CD4T, CD8T, B cells and NK cells), estimated monocyte cell proportions, M values at the AHRR probe (‘cg05575921’) to proxy for smoking status, and the top 10 genetic principal components.

```
glm(formula = as.factor(antidep) ~ scale(AD_MRS) + scale(age) +
    scale(Mono) + scale(lymphocytes) + scale(cg05575921) + as.factor(sex_coded) +
    scale(C1) + scale(C2) + scale(C3) + scale(C4) + scale(C5) +
    scale(C6) + scale(C7) + scale(C8) + scale(C9) + scale(C10),
    family = "binomial", data = MRS_covs_pheno)
```

##### Funding and Acknowledgement

The MARS cohort was sponsored by the Max Planck Society. The UniDep cohort was funded by the Bavarian Ministry of Commerce and by the Federal Ministry of Education and Research in the framework of the National Genome Research Network, Foerderkennzeichen 01GS0481 and the Bavarian Ministry of Commerce. DNA methylation analysis of a subset of both cohorts was financed by ERA-NET NEURON.

We would like to thank all contributors to the research project including physicians, psychologists, study nurses, researchers and research assistants, and of course patients of the hospital of the Max Planck Institute of Psychiatry in Munich and psychiatric hospitals in Augsburg and Ingolstadt.

#### Lothian Birth Cohort 1936

Protocol paper: <https://doi.org/10.1186/s13073-018-0585-7>^43^

##### Cohort Description

The Lothian Birth Cohort 1936 (N = 1, 091) is a follow-up study of the Scottish Mental Surveys distributed to 11 year olds residing in Scotland in 1947, and is designed to examine the distribution and causes of cognitive aging across the human life course^44^. Participants took the survey at 11 years old and were recruited to LBC1936 at the mean age of 70 years. LBC1936 has conducted various waves of assessment, including a series of cognitive, clinical, physical, and social data alongside blood donations which have been used for multi-omics measurement. Initial baseline assessment was conducted in a single visit to the Wellcome Trust Clinical Research Facility at the Western General Hospital in Edinburgh between November 2006 and May 2007. Participants provided written consent before any assessment and/or sampling took place

**Ethical Approval**: Ethical approval was obtained from the Multicentre Research Ethics Committee for Scotland (baseline, MREC/01/0/56), the Lothian Research Ethics Committee (age 70, LREC/2003/2/29), and the Scotland A Research Ethics Committee (ages 73, 76, 79, 07/MRE00/58). All participants provided written informed consent.

##### Antidepressant exposure phenotype

At each wave of assessment, medication currently being taken by participants was recorded in a structured interview. Drugs were assigned to medication classes following the British National Formulary (BNF) paragraph code, with antidepressant codes beginning with ‘04030’ (eTable 1). Those who named any single antidepressant medication were defined as antidepressant-exposed and those who did not name a single antidepressant medication were defined as antidepressant-unexposed. At wave 1, there were 46 exposed and 843 unexposed individuals with high quality DNAm data.

##### DNAm preprocessing

DNAm was profiled from whole blood samples at baseline (N = 1, 004) using HumanMethylation450 BeadChip Kit, Illumina, San Diego, CA, USA. Details on sample preparation and quality check have been reported previously^45^. Firstly, raw intensity data were background corrected and normalised using internal controls. Manual inspection of the array control probe signals was used to identify and remove low quality samples (e.g., samples with inadequate hybridization, bisulfite conversion, nucleotide extension or staining signal). Probes with a low detection rate (p > 0.01 for more than 5% of samples), low call rate (p < 0.01 for < 95% of probes) were removed. Samples whose predicted sex, based on XY probes, did not match reported sex were removed, along with those for whom SNP array genotypes were inconsistent with predicted genotypes derived from the methylation array’s 65 ‘rs’ control probes. After these quality control steps, 457, 047 autosomal probes remained from 895 individuals. M-values were calculated using the “beta2m” function from the *lumi* R package^11^. All M values were normalised using beta-mixture quantile normalisation^46^.

##### Methylation Profile Score

After adhering to the quality control protocols, 206 CpGs were available for the calculation of the MPS, which was derived as a weighted sum of the CpGs. There was no missingness in any of the CpGs included in the score.

##### Antidepressant exposure ~ MPS association model

The association between the antidepressant exposure and the antidepressant exposure MPS was tested using a generalised linear mixed effects model with a logistic link function, using the *‘glmer()’* function from the *lme4* R package^27^, with antidepressant exposure as the outcome. The model included the following predictors with a fixed effect: antidepressant exposure MPS, age at blood sampling, sex, neutrophil cell count, lymphocyte cell count, eosinophil cell count, basophil cell count, M values at the AHRR probe (cg05575921) to proxy for smoking status and the top 4 genetic principal components. Additionally, array processing batch was included as a random effect, to account for additional technical factors.

```
Generalized linear mixed model fit by maximum likelihood (Laplace
  Approximation) [glmerMod]
 Family: binomial  ( logit )
Formula: as.factor(antidep) ~ scale(AD_MRS) + scale(age) + scale(neut) +
    scale(lymph) + scale(cg05575921) + scale(mono) + scale(eosin) +
    scale(baso) + as.factor(sex_coded) + scale(C1) + scale(C2) +
    scale(C3) + scale(C4) + (1 | array)
   Data: MRS_covs_pheno
```

##### Funding and Acknowledgement

The LBC1936 is supported by the Biotechnology and Biological Sciences Research Council, and the Economic and Social Research Council [BB/W008793/1] (which supports SEH), Age UK (Disconnected Mind project), the Medical Research Council [G0701120, G1001245, MR/M013111/1, MR/R024065/1], the Milton Damerel Trust, and the University of Edinburgh. SRC is supported by a Sir Henry Dale Fellowship jointly funded by the Wellcome Trust and the Royal Society (221890/Z/20/Z). Methylation typing of was supported by Centre for Cognitive Ageing and Cognitive Epidemiology (Pilot Fund award), Age UK, The Wellcome Trust Institutional Strategic Support Fund, The University of Edinburgh, and The University of Queensland.

#### Avon Longitudinal Study of Parents and Children

ALSPAC cohort protocol paper(s): <https://doi.org/10.1093/ije/dys064>^47^, <https://doi.org/10.1093/ije/dys066>^48^ DNAm protocol paper (ARIES): <https://doi.org/10.1093/ije/dyv072>^49^

##### Cohort Description

Pregnant women resident in Avon, UK with expected dates of delivery between 1st April 1991 and 31st December 1992 were invited to take part in the study. 20,248 pregnancies have been identified as being eligible and the initial number of pregnancies enrolled was 14,541. Of the initial pregnancies, there was a total of 14,676 foetuses, resulting in 14,062 live births and 13,988 children who were alive at 1 year of age^48^. When the oldest children were approximately 7 years of age, an attempt was made to bolster the initial sample with eligible cases who had failed to join the study originally. As a result, when considering variables collected from the age of seven onwards (and potentially abstracted from obstetric notes) there are data available for more than the 14,541 pregnancies mentioned above: The number of new pregnancies not in the initial sample (known as Phase I enrolment) that are currently represented in the released data and reflecting enrolment status at the age of 24 is 906, resulting in an additional 913 children being enrolled (456, 262 and 195 recruited during Phases II, III and IV respectively. The phases of enrolment are described in more detail in the cohort profile paper and its update^50^. The total sample size for analyses using any data collected after the age of seven is therefore 15,447 pregnancies, resulting in 15,658 foetuses. Of these 14,901 children were alive at 1 year of age. As part of Accessible Resource for Integrated Epigenomic Studies (ARIES), a sub-sample ALSPAC children, mothers and partners had DNAm assayed using the Illumina Infinium HumanMethylation450 or MethylationEPIC Beadchip array from blood samples collected from ALSPAC children and their parents at multiple time points from birth to middle age^49^. Study data were collected and managed using REDCap electronic data capture tools hosted at the University of Bristol. REDCap (Research Electronic Data Capture) is a secure, web-based software platform designed to support data capture for research studies^51^. Please note that the study website contains details of all the data that is available through a fully searchable data dictionary and variable search tool (<http://www.bristol.ac.uk/alspac/researchers/our-data/>).

**Ethical Approval**: Ethical approval for the study was obtained from the ALSPAC Ethics and Law Committee and the Local Research Ethics Committees. Consent for biological samples has been collected in accordance with the Human Tissue Act (2004).

##### Antidepressant exposure phenotype

Medications, including antidepressants, were recorded for children of ALSPAC at age 24 at the clinic appointment for the ‘Focus @ 24’ timepoint (‘’), when blood sampling also took place. Additionally, the ‘Life @ 28’ questionnaire, collected between December 2020 and April 2021, had a Mental Health Treatments section which included in-depth questions regarding antidepressant use, including the duration of adherence^52^.

Those who reported antidepressant use at the clinic (FieldID: FCKO1103) and/or those who reported antidepressant use for over 4 years at the questionnaire, that also overlapped with the clinic (FieldIDs: YPH7000, YPH7017 and YPH7047) were defined as antidepressant exposed. Those who reported no antidepressant use at the clinic assessment and/or no antidepressant use at the questionnaire or were not taking antidepressants at the time of the blood sample were classed as antidepressant-unexposed. In total, there were 43 exposed and 758 unexposed individuals with quality DNAm data for analysis.

##### DNAm preprocessing

Illumina Infinium HumanMethylation450 and MethylationEPIC Beadchip arrays were used to assess genome-wide DNAm patterns in peripheral blood. Samples across different time-points were distributed in a semi-random manner across slides to mitigate batch effects. Data quality assessment, pre-processing and normalization was performed using the R package *meffil* as previously described^7^. Briefly, probes undetected (detection p-value > 0.01) or with low bead count (<3) in >10% of samples were removed. Samples were removed if there was a mismatch between their predicted sex and recorded sex or if >10% of probes were undetected. Raw probe intensities were adjusted for potential dye bias, background corrected using the *‘noob’* method^8^ and normalized using functional normalization^38^. Cell proportions (CD4T, CD8T, NK cells, B cells, Granulocytes and Monocytes) were estimated using the Houseman mode in the *minfi* R package^5^. Lymphocyte cell proportions were then calculated as the aggregate of the estimated CD4T, CD8T, NK and Bcell cell proportions. M-value transformation was conducted using the *‘beta2m()’* function in the *lumi* R package^11^. The M-values were then standardised using Z-score normalisation using the *‘scale()’* function in R. In total, 857,809 CpG probes remained for 801 individuals.

##### Methylation Profile Score

After adhering to the quality control protocols, all 212 CpGs were available for the calculation of the MPS, which was derived as a weighted sum of the CpGs. The rate of missingness per CpG ranged from 0-6%. In the case that an individual had a missing value for a CpG(s), the probe(s) were excluded from the MPS (i.e., given a weight of 0).

##### Antidepressant exposure ~ MPS association model

The association between the antidepressant exposure and MPS was tested using a generalised linear model, using the *‘glm()’* function from the *stats* R package^30^, with antidepressant exposure as the outcome. The model included the following predictors: antidepressant exposure MPS, age at sampling, sex, monocyte cell proportions, lymphocyte cell proportions and AHRR M values (‘cg05575921’) to proxy for smoking status.

```
glm(formula = as.factor(antidep) ~ scale(AD_MRS) + scale(age) +
    scale(Mono) + scale(lymphocytes) + scale(cg05575921) + as.factor(sex_coded),
    family = "binomial", data = MRS_covs_pheno)
```

##### Funding and Acknowledgements

The UK Medical Research Council and Wellcome (Grant Ref: 217065/Z/19/Z) and the University of Bristol provide core support for ALSPAC. ASFK is funded by a Wellcome Early Career Award (Grant ref: 227063/Z/23/Z). A comprehensive list of grants funding is available on the ALSPAC website (<http://www.bristol.ac.uk/alspac/external/documents/grant-acknowledgements.pdf>). This publication is the work of the authors and ED, PY & ASFK will serve as guarantors for the ALSPAC contents of this paper. We thank all the families who took part in this study, the midwives for their help in recruiting them, and the whole ALSPAC team, which includes interviewers, computer and laboratory technicians, clerical workers, research scientists, volunteers, managers, receptionists and nurses.

#### Environmental risk (E-Risk) Longitudinal Twin Study

Protocol paper: <https://doi.org/10.1111/1469-7610.00082>^53^

##### Cohort Description

The Environmental Risk (E-Risk) Longitudinal Twin Study is a representative birth cohort study of 2,232 twins born in England and Wales between 1994-1995. Baseline data collection occurred between 1999-2000 when 1,116 families (93% of those eligible) with same-sex 5-year-old twins participated in home-visit assessments. Full details about the sample are reported elsewhere^53^. This sample comprised 56% monozygotic and 44% dizygotic twin pairs; sex was evenly distributed within zygosity (49% male). This is a nationally representative sample - E-Risk families’ addresses are currently a near-perfect match to the deciles of the UK’s 2015 Lower-layer Super Output Area Index of Multiple Deprivation (IMD). Follow-up home-visits were conducted when children were aged 7, 10, 12, and 18 (participation rates were 98%, 96%, 96%, and 93%, respectively). At age 18, 2066 participants were assessed. Average age at time of assessment was 18.4 years (SD=0.36); all interviews were conducted after the 18th birthday. There were no differences between those who did and did not take part at age 18 in terms of socio-economic status (SES) assessed when the cohort was initially defined (χ2=0.86, p=0.65), age-5 IQ scores (t=0.98, p=0.33), and age-5 behavioural (t=0.40, p=0.69) or emotional (t=0.41, p=0.68) problems.

**Ethical Approval** The Joint South London and Maudsley and Institute of Psychiatry Research Ethics Committee approved each study phase. Parents gave informed consent, and twins gave assent between 5 and 12 years and then informed consent at age 18.

##### Antidepressant exposure phenotype

Antidepressant exposure was assessed at the age- 18 home visit, where participants were asked: “Have you taken any medicines in the last two weeks?” and they were then shown a card listing common medications including antidepressants. If they reported antidepressant use then they were asked to provide the name of the specific drug(s) they had taken, what condition it was being used to treat, and the duration of use. Medicines were classed by a consultant psychiatrist as antidepressants following BNF criteria. Those who reported using one or more antidepressant medications in the previous 2 weeks were defined as antidepressant-exposed, and those who did not report any use of antidepressant medication in this period were defined as antidepressant-unexposed. In total, there were 36 exposed and 1,622 unexposed with high quality DNAm data.

##### DNAm preprocessing

DNAm was profiled using 1,700 whole blood samples collected at the age-18 home visit, using the Illumina Infinium HumanMethylation450 BeadChip. Of the available samples, 31 samples were excluded due to a low DNA concentration. Approximately 500ug of DNA from each sample was treated with sodium bisulfite using EZ-96 DNA Methylation Kit (Zymo Research, CA, USA). DNAm was then quantified using the Illumina Infinium HumanMethylation450 BeadChip (Illumina Inc, CA, USA) run on an Illumina iScan System (Illumina, CA, USA) using the manufacturers’ standard protocol. Twin pairs were randomly assigned to bisulfite-coversion plates and Illumina 450K arrays, with siblings positioned adjacently to minimise batch effects.

The following quality control (QC) pipeline was applied to all 1,669 unique individuals and was performed in the R statistical programming environment. Data was imported using the *‘methylumIDAT()’* function from the *methylumiII* package^54^. Samples were excluded if median methylation (‘M’) and unmethylation (‘U’) signal intensities were < 2,500 (n = 10). The fully methylated control samples were first identified using their intensity profiles (signal characteristic of being fully methylated) and used to confirm the absence of any plate rotations or plate mislabelling had occurred before being removed from the dataset. The efficacy of the sodium bisulfite conversion reaction was assessed using ten control probes included on the 450K array, and probes with a “conversion score” < 80 were removed. Multidimensional scaling was performed for DNAm probes on each of the sex chromosomes and compared to reported sex, finding a discrepancy for 2 samples from 2 different monozygotic twin pairs. Further investigation into these twin pairs concluded that discordant samples had been mistakenly switched in the lab, and therefore paired with the twin from other twin pair. Correction of ID identifiers led to a 100% match. The genotype concordance between SNP probes on the 450K array and data generated using Illumina OmniExpress24v1.2 genotyping BeadChips was assessed to confirm genetic identity of DNA samples. Data were available for 35 of the 65 SNP probes from both platforms for 1,638 (98.7%) of the samples. A total of 1,658 samples passed the stringent QC pipeline. The *‘pfilter()’* function from the *wateRmelon* package^4^ in R was used to process the data and exclude samples which had > 1% of probes having a detection p value > 0.05 (n = 0). Probes which had a beadcount < 3 in 5% of the samples (n = 567) and those with a detection p value > 0.05 in >1% of the samples were removed from the dataset. The data was then normalised using the dasen method from the *wateRmelon* package^4^. Prior to any analyses, probes with common (> 5% MAF) SNPs within 10bp of the single base extension and probes with sequences previously identified as potentially hybridising to multiple genomic loci were excluded (n = 52,760)^21^. After QC and annotation of CpGs to the 450k array 430,802 sites remained for analysis. Methylation beta-values were transformed to M-values using logit transformation. These Illumina DNA methylation data are accessible from the Gene Expression Omnibus (accession code: GSE105018).

##### Methylation Profile Score

After adhering to the quality control protocols, 207 CpGs were available for the calculation of the MPS, which was derived as a weighted sum of the CpGs. There was no missingness in any of the CpGs included in the score.

##### Antidepressant exposure ~ MPS association model

The association between antidepressant exposure and the MPS was assessed using a generalised linear mixed model, using the ‘glmer()’ function from the ‘lme4’ R package1, with antidepressant exposure as the outcome. The model included the following predictors: antidepressant exposure MPS, sex, predicted monocyte cell proportions, predicted lymphocyte cell proportions, M values at the AHRR probe (cg05575921), and family ID as a random effect. The model was specified with a logit link function and 20 quadrature points in the adaptive Gaussian quadrature approximation for integrating over the random effect. The optimisation algorithm used was the Bound Optimisation by Quadratic Approximation (BOBYQA) with a maximum of 100,000 function evaluations.

assoc_mod<- glmer(as.factor(antidep) ~ scale(AD_MRS) +
 as.factor(sex_coded) + scale(Mono) + scale(lymphocytes)+ scale(as.numeric(Chip))+ scale(cg05575921)+ (1|FID), data=MRS_covs_pheno, family=binomial(link = "logit"), nAGQ = 20,
 control = glmerControl(optimizer = "bobyqa", optCtrl = list(maxfun = 100000)))

##### Funding and Acknowledgements

The E-Risk Study is funded by grants from the UK Medical Research Council [G1002190; MR/X010791/1]. Additional support was provided by the US National Institute of Child Health and Human Development [HD077482] and the Jacobs Foundation. Chloe C. Y. Wong was supported by the National Institute for Health and Care Research (NIHR) Maudsley Biomedical Research Centre at South London and Maudsley NHS Foundation Trust and King’s College London [NIHR203318]. Helen L. Fisher was supported by the Economic and Social Research Council (ESRC) Centre for Society and Mental Health at King’s College London [ES/S012567/1]. The views expressed are those of the authors and not necessarily those of the NIHR, the Department of Health and Social Care, the ESRC, or King’s College London.

We are grateful to the E-Risk study mothers and fathers, the twins, and the twins’ teachers for their participation. Our thanks to Professors Terrie Moffitt and Avshalom Caspi, the founders of the E-Risk study, and to the E-Risk team for their dedication, hard work, and insights.

### Prospective Generation Scotland Follow Up

#### Stratifying Resilience and Depression Longitudinally

Protocol paper: <https://doi.org/10.1093/ije/dyx115>^56^

##### Cohort Description

The Stratifying Resilience and Depression Longitudinally (STRADL) cohort^56^, is a follow up study of a subset of GS participants (n = 1, 168) on average five years after baseline assessment. STRADL was primarily aimed for the assessment of mental health, specifically MDD. Data collection included socio-economic and demographic profiling, laboratory samples, extensive psychological assessment and magnetic resonance imaging (MRI)^57^. Further details of the STRADL cohort and GS protocol are published elsewhere^1,57^.

**Ethical Approval**: All components of STRADL received formal, national ethical approval from the NHS Tayside committee on research ethics (reference 14/SS/0039).

##### Antidepressant exposure phenotype

Medication use in STRADL was assessed using a free-response questionnaire asking participants to list prescribed or bought medications, classifying antidepressants using the British National Formulary (BNF) framework. We took forward those who did not self-report antidepressant use (controls) at Generation Scotland baseline (n = 15, 028) and who were present in the STRADL subsample (n = 901). Those who did or did not list an antidepressant medication at the STRADL reassessment time point were defined as antidepressant-exposed and unexposed respectively. In total, there were 46 exposed and 617 unexposed individuals with high-quality DNAm data.

##### DNAm preprocessing

DNAm was profiled from whole-blood samples taken at the STRADL timepoint using the Illumina HumanMethylation EPIC array. The processing was completed in two sets and used the same methodology as those collected in the wider Generation Scotland cohort at baseline^1^. The R package *Meffil*^7^ was used to remove samples which: had a mismatch between genotyped and DNAm-predicted sex, had > 0.5% of CpGs with a detection P > 0.01, were outliers for bisulphite conversion control probes, had median signal intensity >3 standard deviations lower than expected and those with evidence of dye bias. The R package *shinyMethyl*^3^ was used to exclude outliers based on visual inspection of the log median intensity of methylated vs unmethylated signals per array. Following this, *Meffil* was again implemented to identify and exclude probes with a beadcount < 3 in > 5% of the samples and/or probes in which > 1% of the samples had a detection of P > 0.01. Multidimensional scaling (MDS) was performed to inspect for further outlier samples and identified 40 males outlying according to the X chromosome DNAm levels and were subsequently removed from the analysis. Data were renormalised and inspection of MDS confirmed no further outliers present across both sets. Cell proportions (CD4T, CD8T, NK cells, B cells, Granulocytes and Monocytes) were estimated using the Houseman mode in the *minfi* R package^5^. Lymphocyte cell proportions were then calculated as the aggregate of the estimated CD4T, CD8T, NK and Bcell cell proportions. Data were then normalised using the dasen method in the *wateRmelon* R package^4^ and converted to M values using the *‘beta2m()’* function in the *lumi* R package^11^. After quality control, there were 793, 706 (n = 503) and 775, 284 (n = 372) CpGs available from set 1 and 2 respectively. The sets were filtered to CpGs common to both and joined together, resulting in 774, 073 CpGs for 875 individuals.

##### Methylation Profile Score

After adhering to the quality control protocols, all 212 CpGs were available for the calculation of the MPS, which was derived as a weighted sum of the CpGs. There was no missingness in any of the CpGs included in the score.

##### Antidepressant exposure ~ MPS association mode

The association between the antidepressant exposure and MPS was tested using a generalised linear mixed effect linear model, using the *‘glmer()’* function from the *lme4* R package^27^. Directly measured fixed effect covariates included age, sex, AHRR probe M-values (‘cg05575921’) to proxy for smoking status, and estimated white blood cell proportions (monocytes and lymphocytes). The array processing batch and family ID was included as a random effect to account for additional technical factors and relatedness within the cohort.

```
Generalized linear mixed model fit by maximum likelihood (Laplace
  Approximation) [glmerMod]
 Family: binomial  ( logit )
Formula: as.factor(antidep) ~ scale(AD_MRS) + scale(age) + scale(Mono) +
    scale(lymphocytes) + scale(cg05575921) + as.factor(sex_coded) +
    (1 | batch) + (1 | famid)
```

##### Funding and Acknowledgement

STRADL is supported by the Wellcome Trust through a Strategic Award (reference 104036/Z/14/Z). The Chief Scientist Office of the Scottish Government Health Department (CZD/16/6) and the Scottish Funding Council (HR03006) provided core support for Generation Scotland. A.M.M. is supported by the Dr Mortimer and Theresa Sackler Foundation. Funding from the Medical Research Council and Biotechnology and Biological Sciences Research Council is gratefully acknowledged (MR/K026992/1). We would like to express gratitude to all individuals who have taken part in both GS:SFHS and STRADL, and the entire project team including academic researchers, administrative staff, research managers and statisticians.

### Results

#### Methylome wide association study

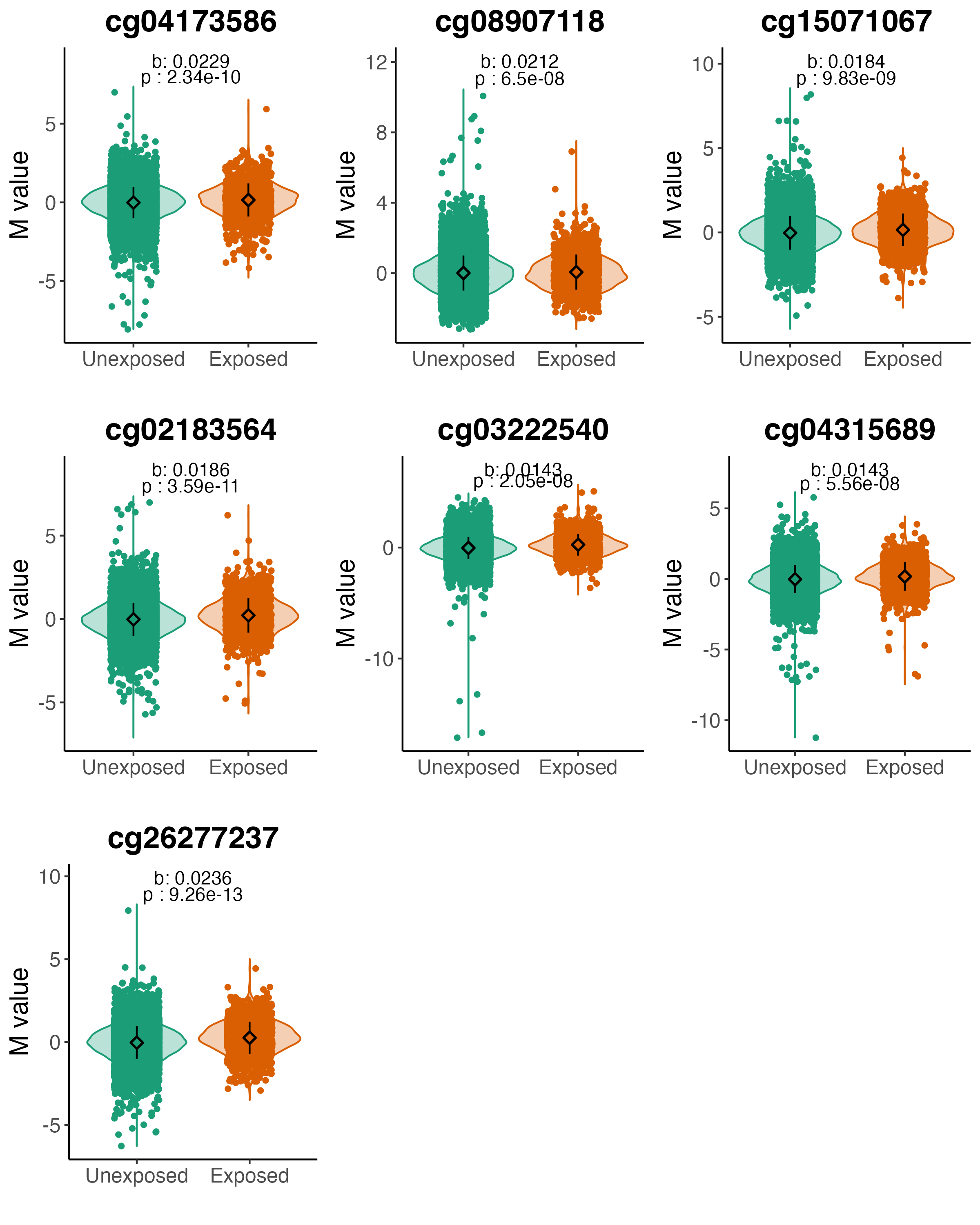

**Figure** **16:** Methylation in antidepressant-exposed and antidepressant-unexposed individuals for significant probes in the self-reported AD use (B = Beta calculated in the self-report MWAS, p = p value calculated from the self-report MWAS).

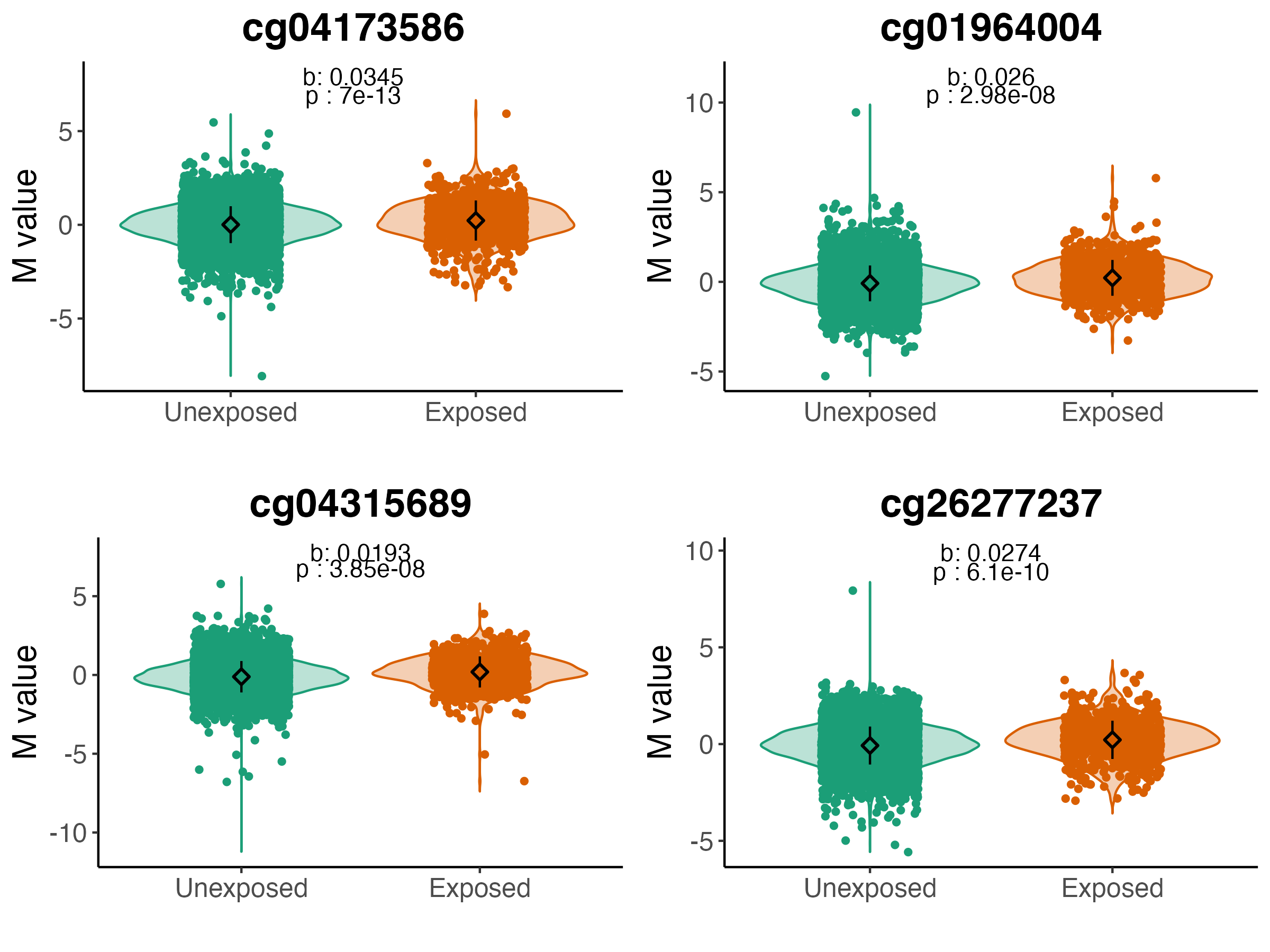

**Figure** **17:** Methylation in antidepressant-exposed and antidepressant-unexposed for significant probes in the prescription-derived AD use (B = Beta calculated in the prescription-derived MWAS, p = p value calculated from the prescription-derived MWAS).

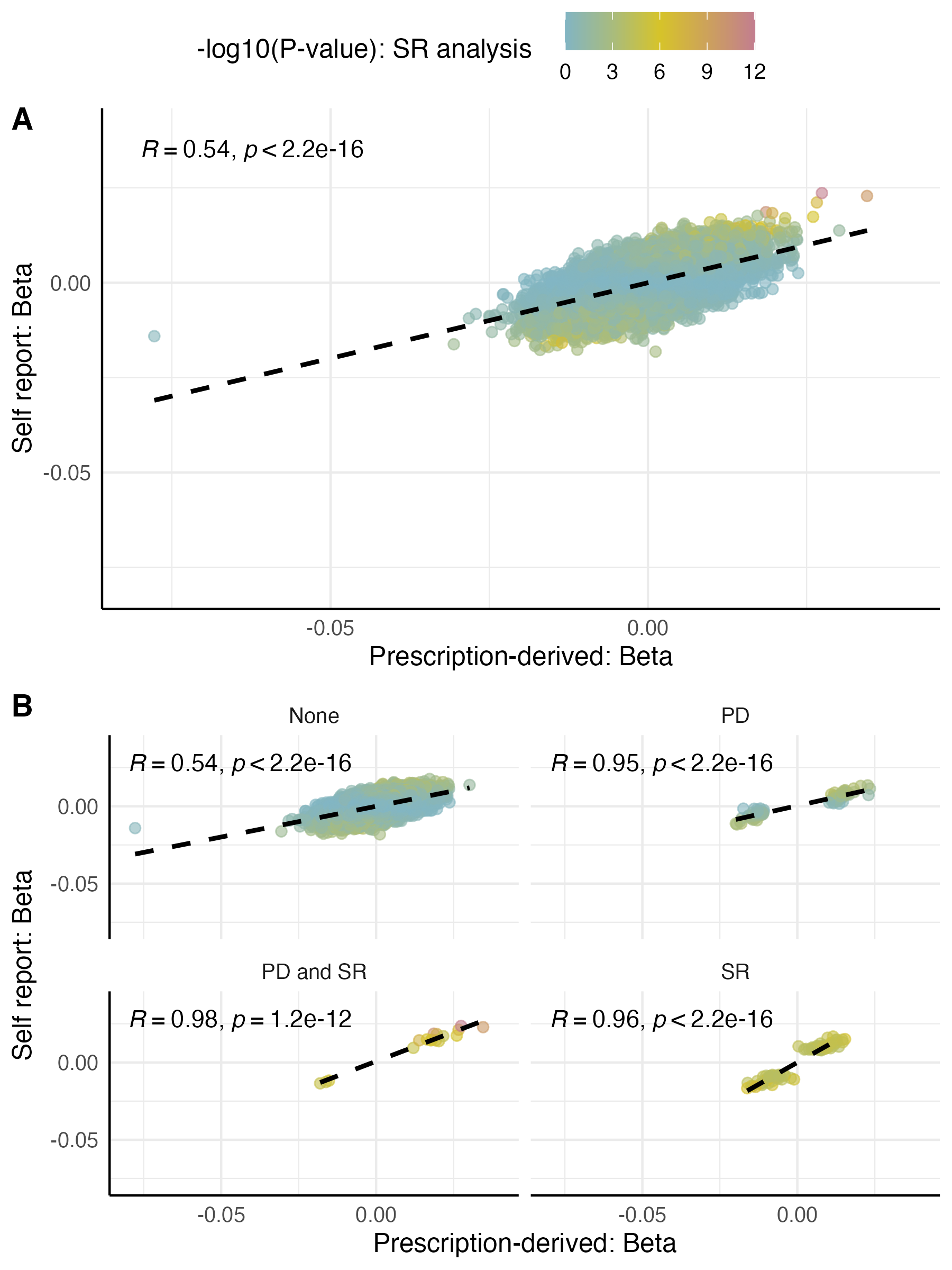

**Figure** **18:** Beta effect estimates for CpGs from the self-report MWAS and the prescription-derived MWAS. A) All CpG probes effect sizes, colored by the p-value in the self-report analysis. B) Subplots of the probes stratified into whether probes are present in the top 100 CpGs in both analyses (bottom left), just the self-report (top right) or just the prescription-derived analysis (bottom right), or not in the top 100 CpGs for either analysis (Top left). PD = Prescription-derived, SR = Self-report.

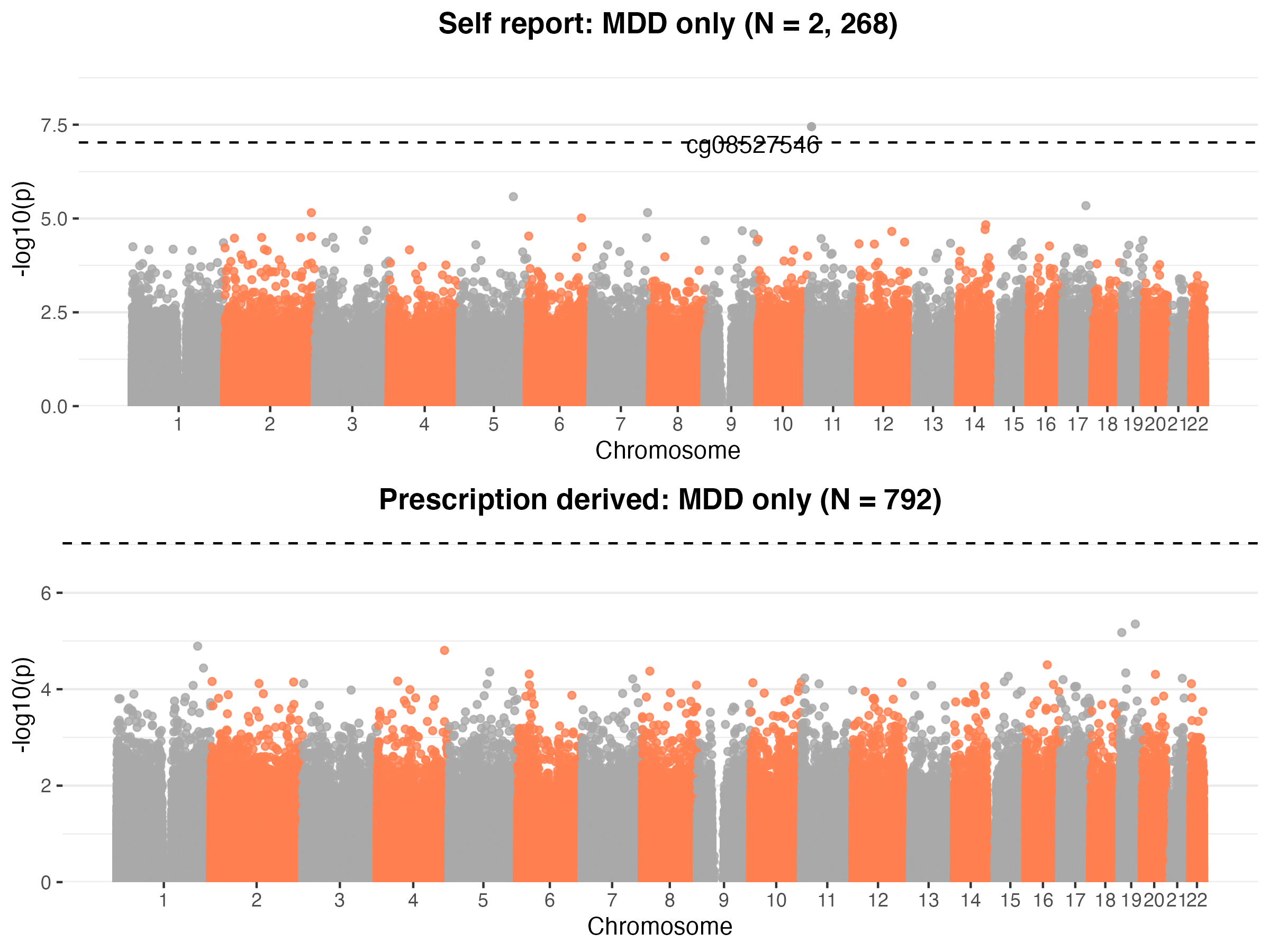

**Figure** **19:** Methylome-wide association study of the self-report (top) and prescription-derived (bottom) phenotypes, when both antidepressant-exposed and antidepressant-unexposed groups are are filtered to those with a lifetime MDD status.

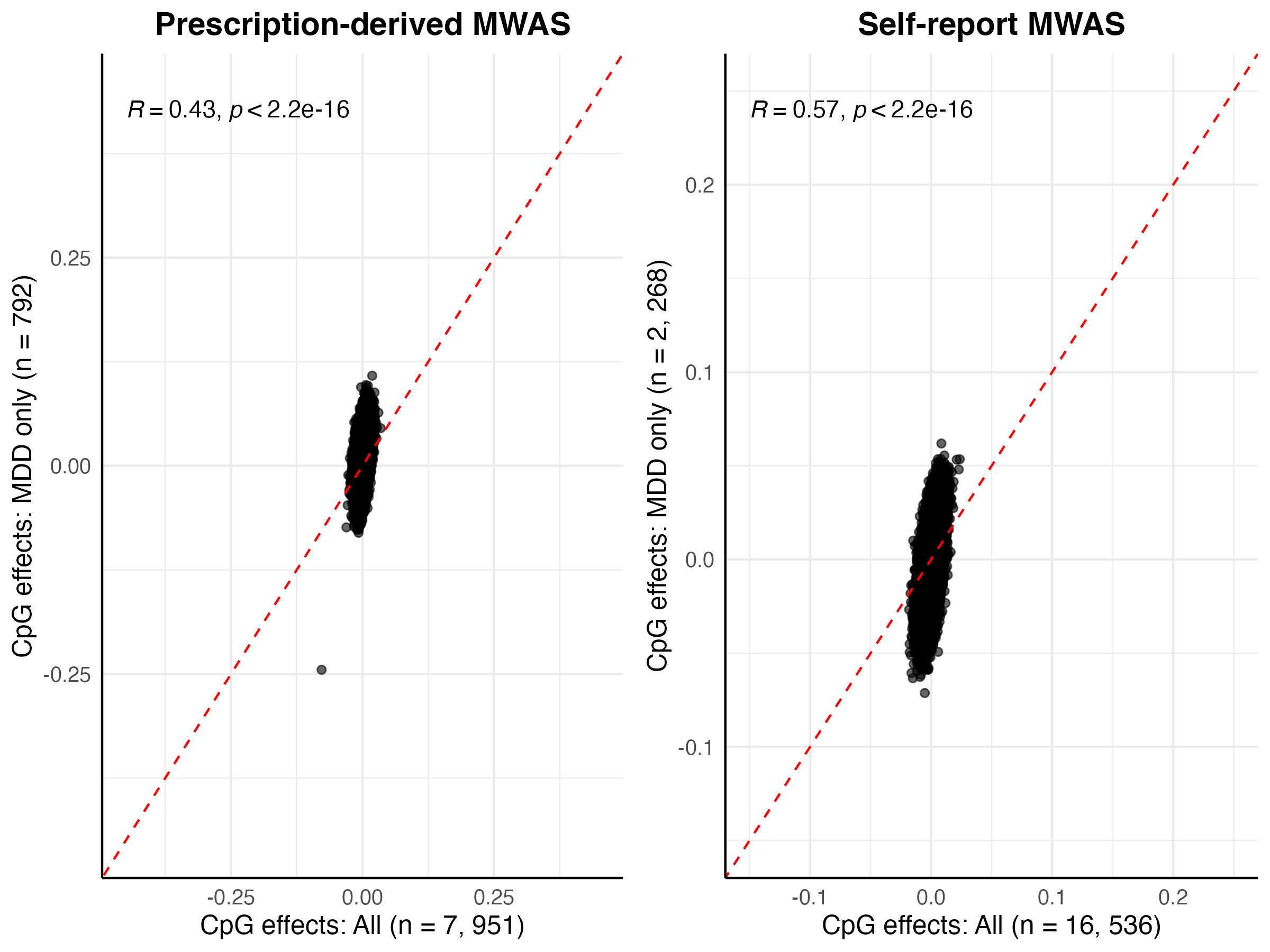

**Figure** **20:** The correlation of probe effect sizes for the self-report (right) and prescription-derived (left) MWAS conducted on all individuals (x-axis) and on a subset of individuals with lifetime MDD status (y-axis). R = Pearson Correlation Coefficient.

#### Differentially Methylated regions

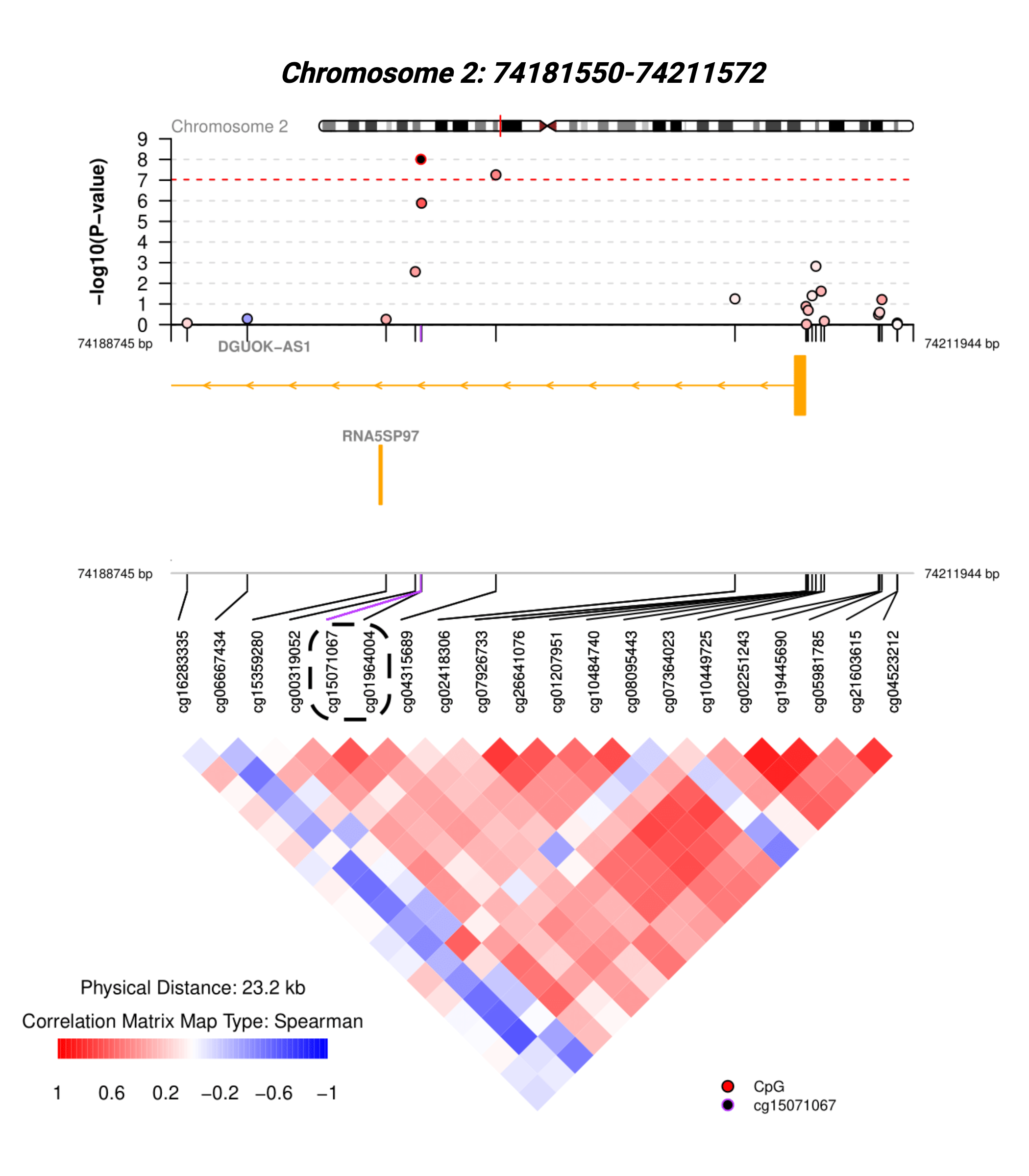

**Figure** **21:** The significant DMR from the self-report MWAS. The top panel presents the CpGs within and near the DMR, coloured by their correlation with cg15071067. The orange bars show gene-tracks, and the bottom heatmap presents the correlation matrix of the CpGs. Plot was created using the coMET R package^58^. DMR = Differentially methylated region.

#### Overlap of PD and SR CpGs and their annotated gene lists

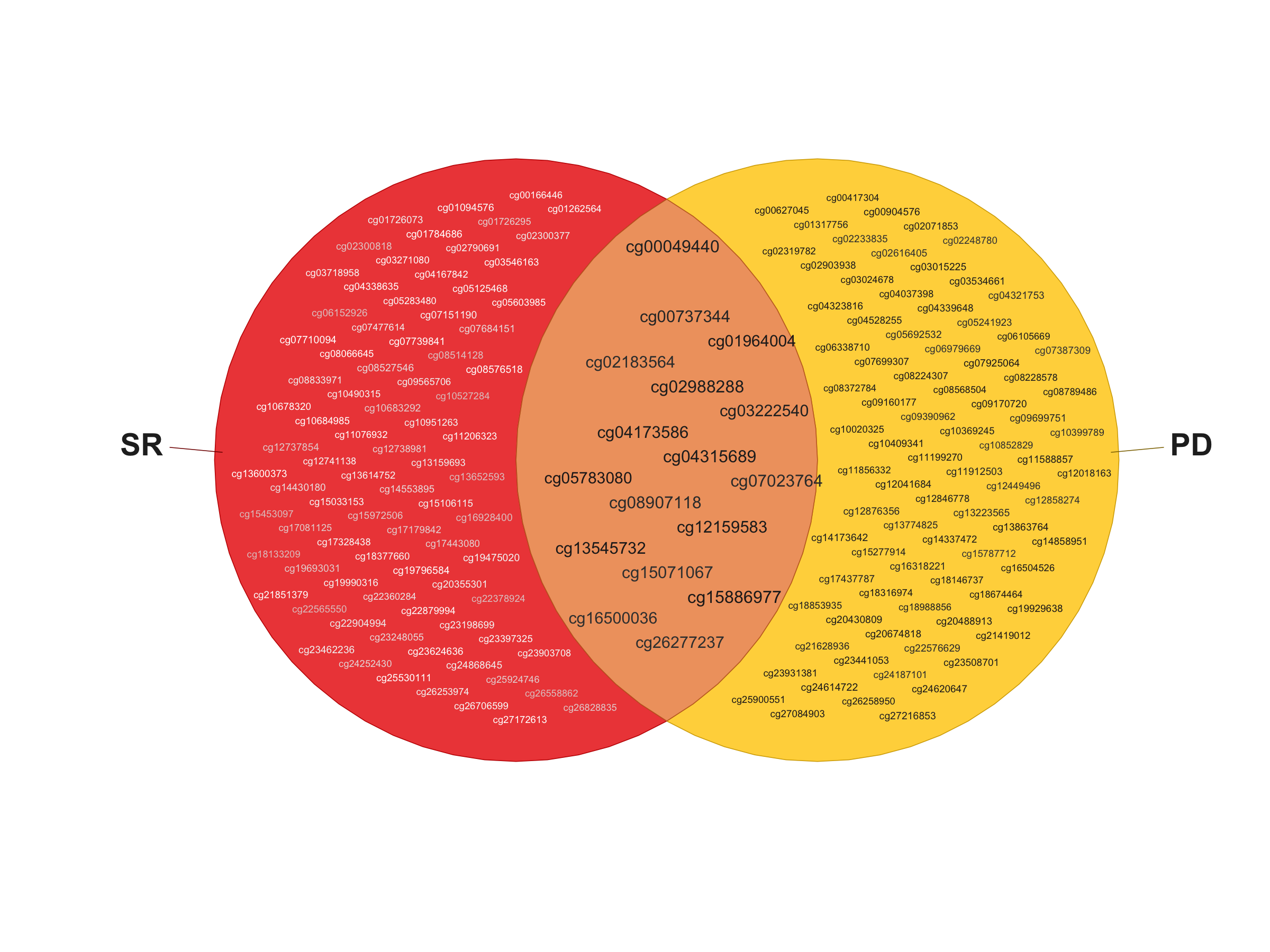

**Figure** **22:** Top 100 CpGs from the self-report and prescription-derived MWAS. There are 17 overlapping CpGs, which is significantly more than expected by chance (p = 1.95x10^-48^).

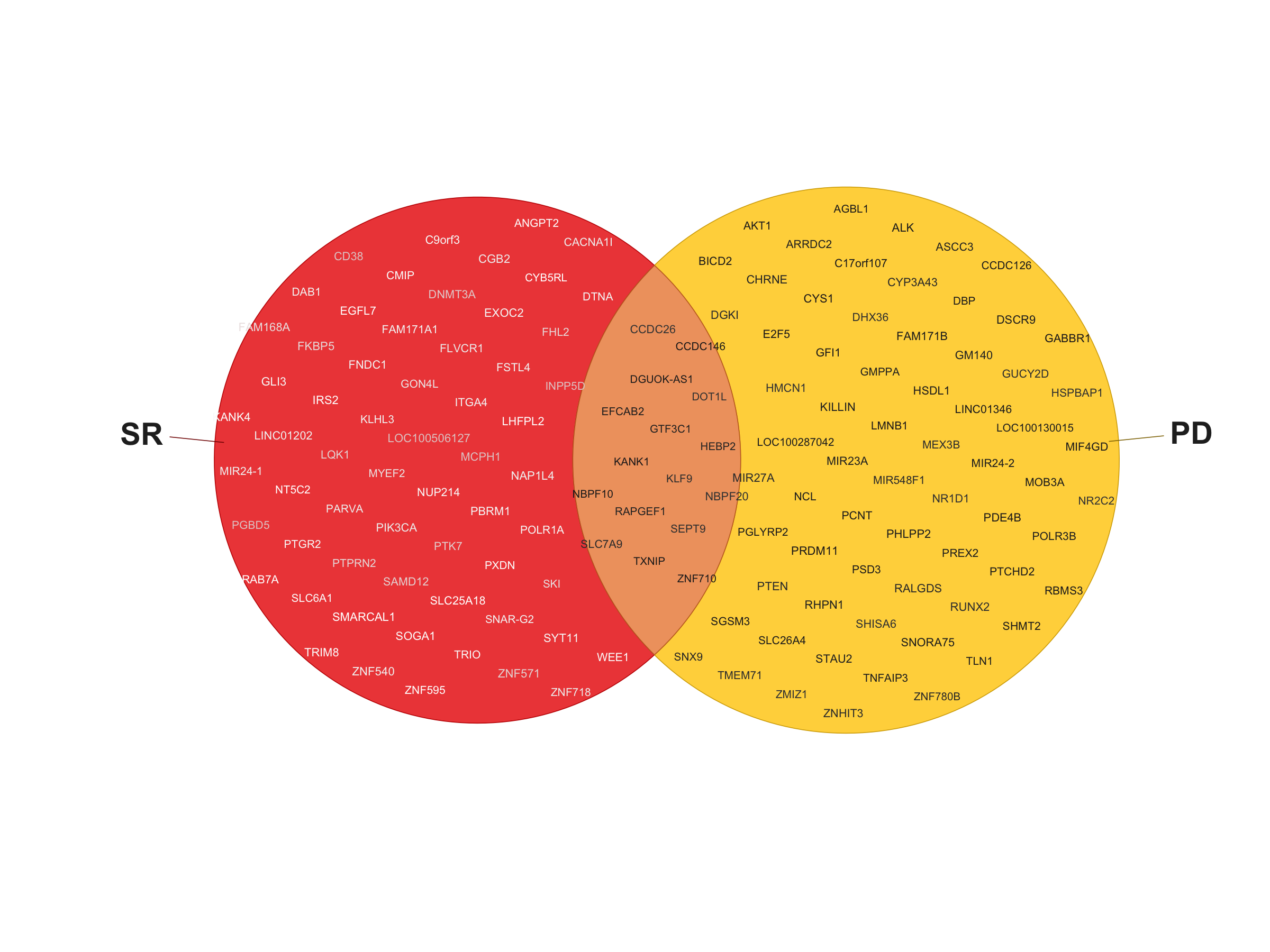

**Figure** **23:** The genes which map to the top 100 CpGs from the self-report (n = 77) and prescription-derived (n = 83) MWAS. There were 16 overlapping genes from each gene list, which was significantly more than expected by chance (p = 1.3x10^-25^).

#### Tissue Specificity

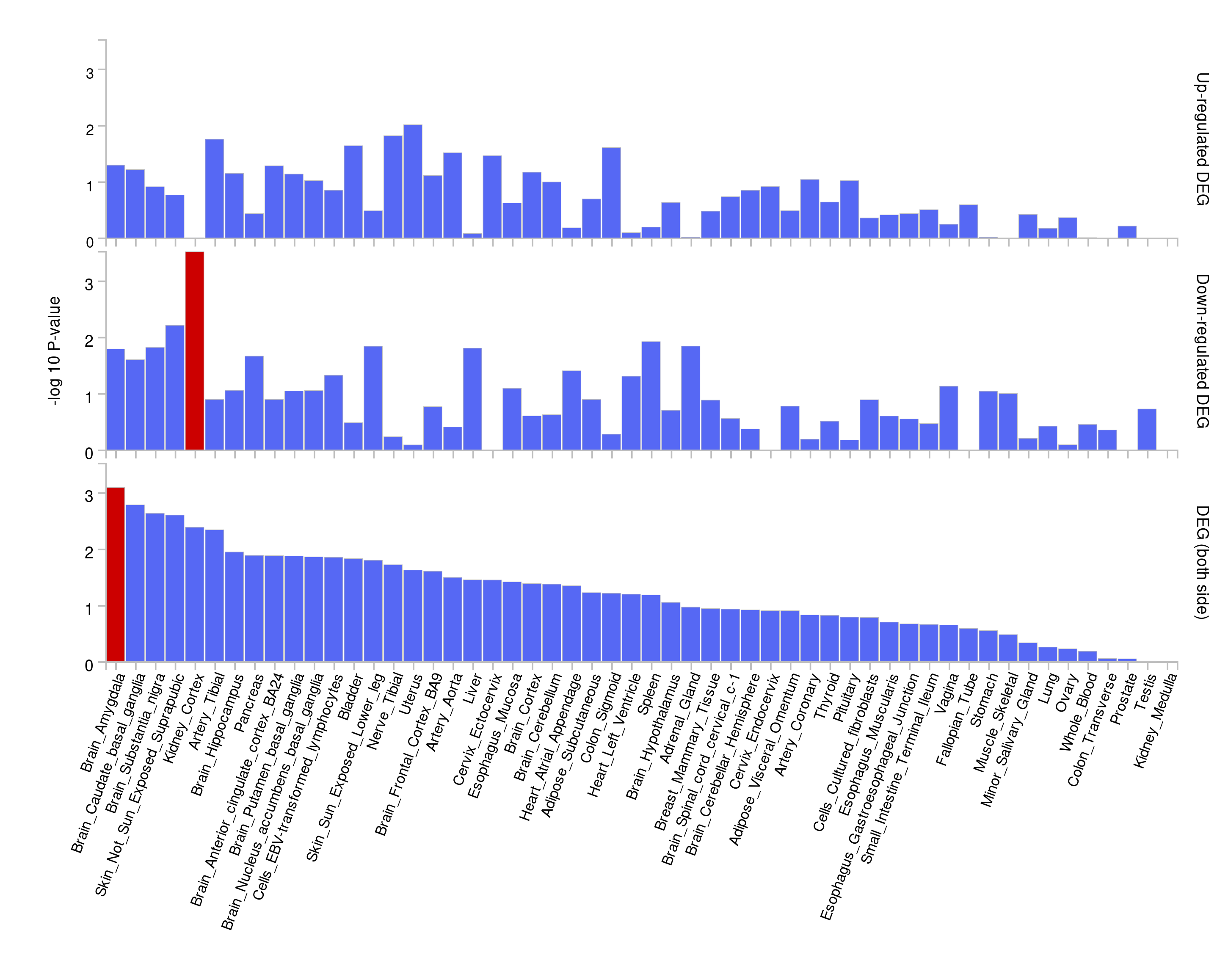

**Figure** **24:** Enrichment of the self-report gene set and 54 specific tissues from GTEx v8 (returned from the GENE2FUNC tool in FUMA). The background gene set was all genes which could be annotated to the Illumina EPIC array, derived using the Infinium MethylationEPIC BeadChip database.

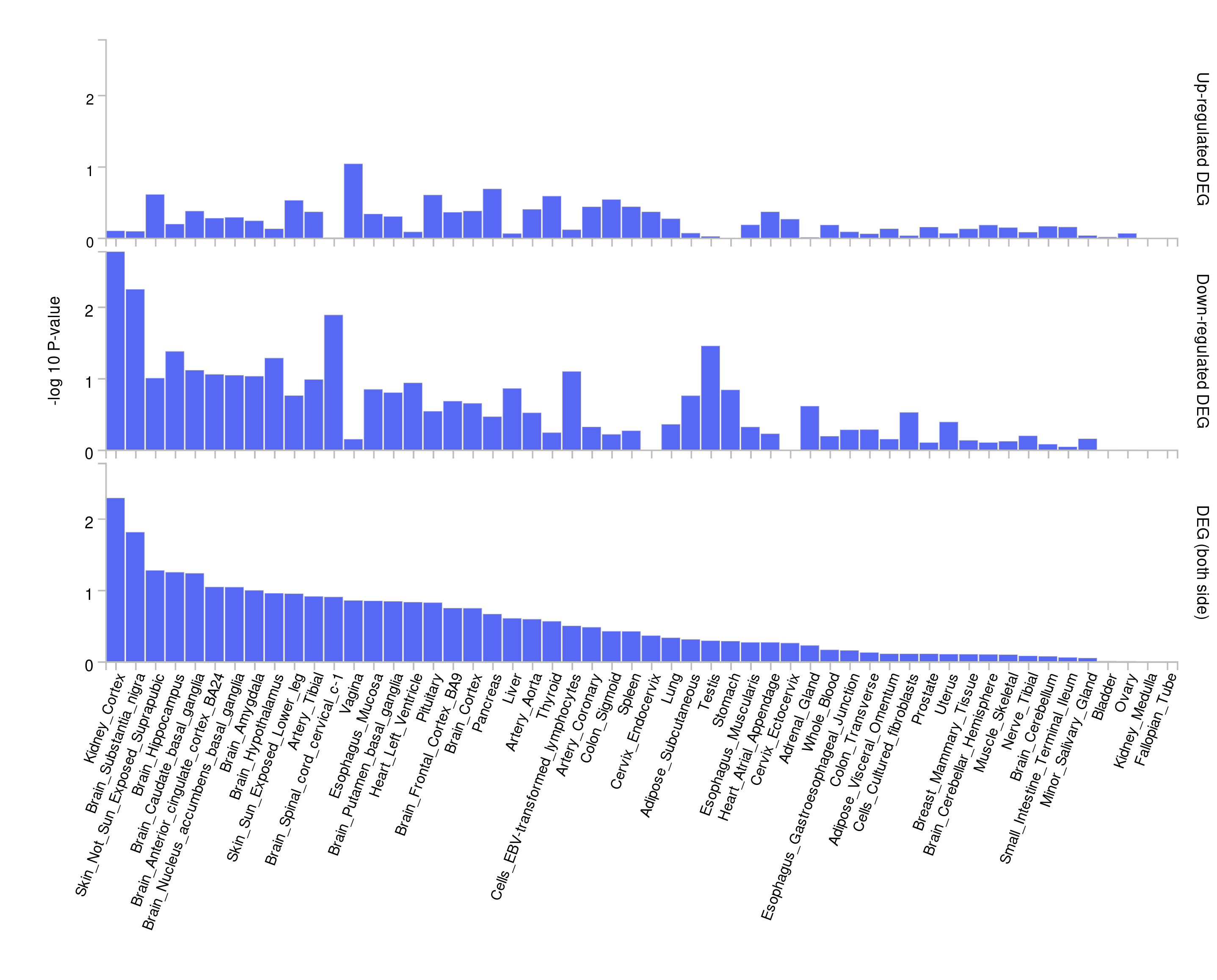

**Figure** **25:** Enrichment of the prescription-derived gene set and 54 specific tissues from GTEx v8 (returned from the GENE2FUNC tool in FUMA). The background gene set was all genes which could be annotated to the Illumina EPIC array, derived using the Infinium MethylationEPIC BeadChip database.

#### Gene Ontology analysis

##### SynGo

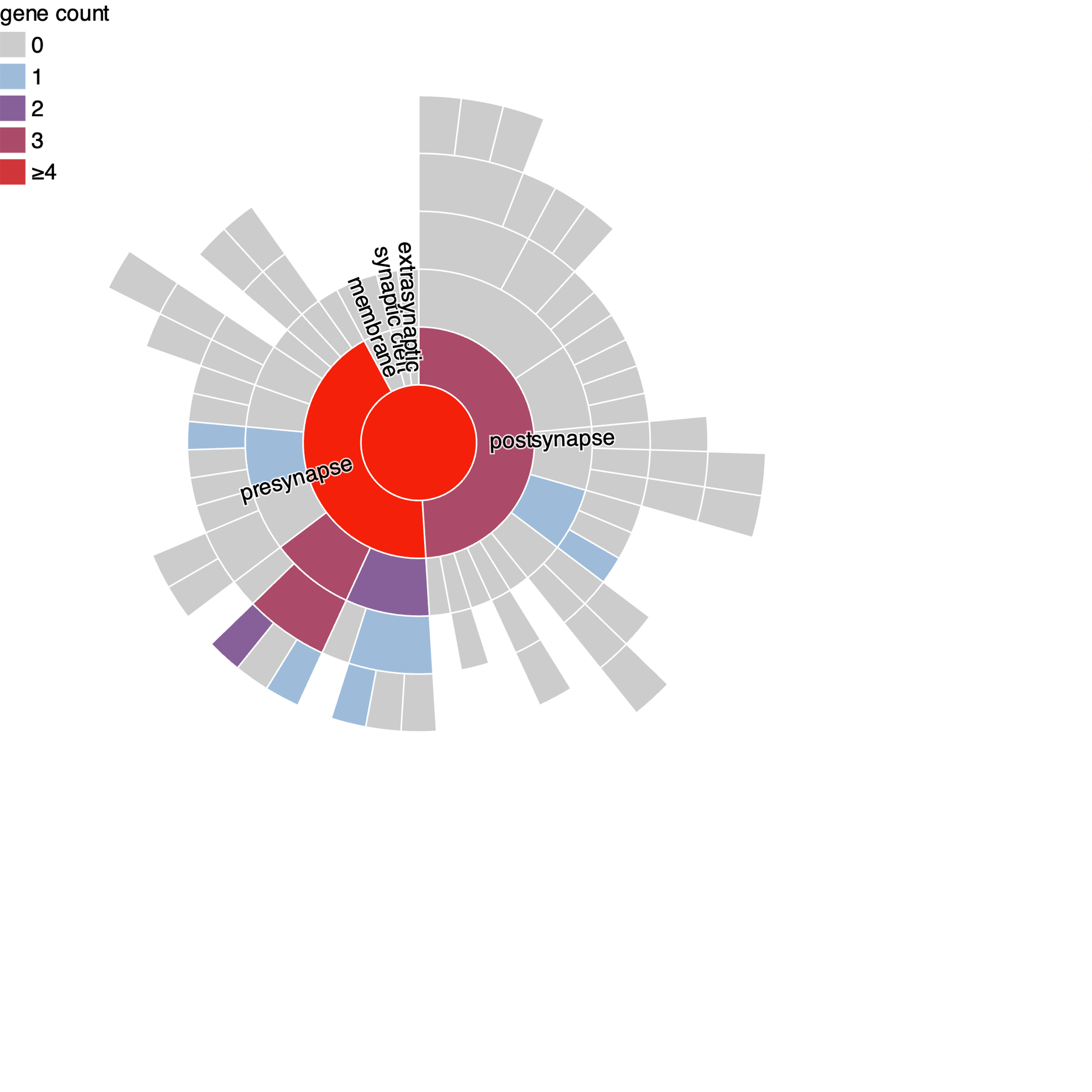

**Figure** **26:** Visualisation of the SynGO terms which are annotated by genes in the self-report gene set, separated by presynaptic (left) or post synaptic (right) location (returned from SynGO web portal).

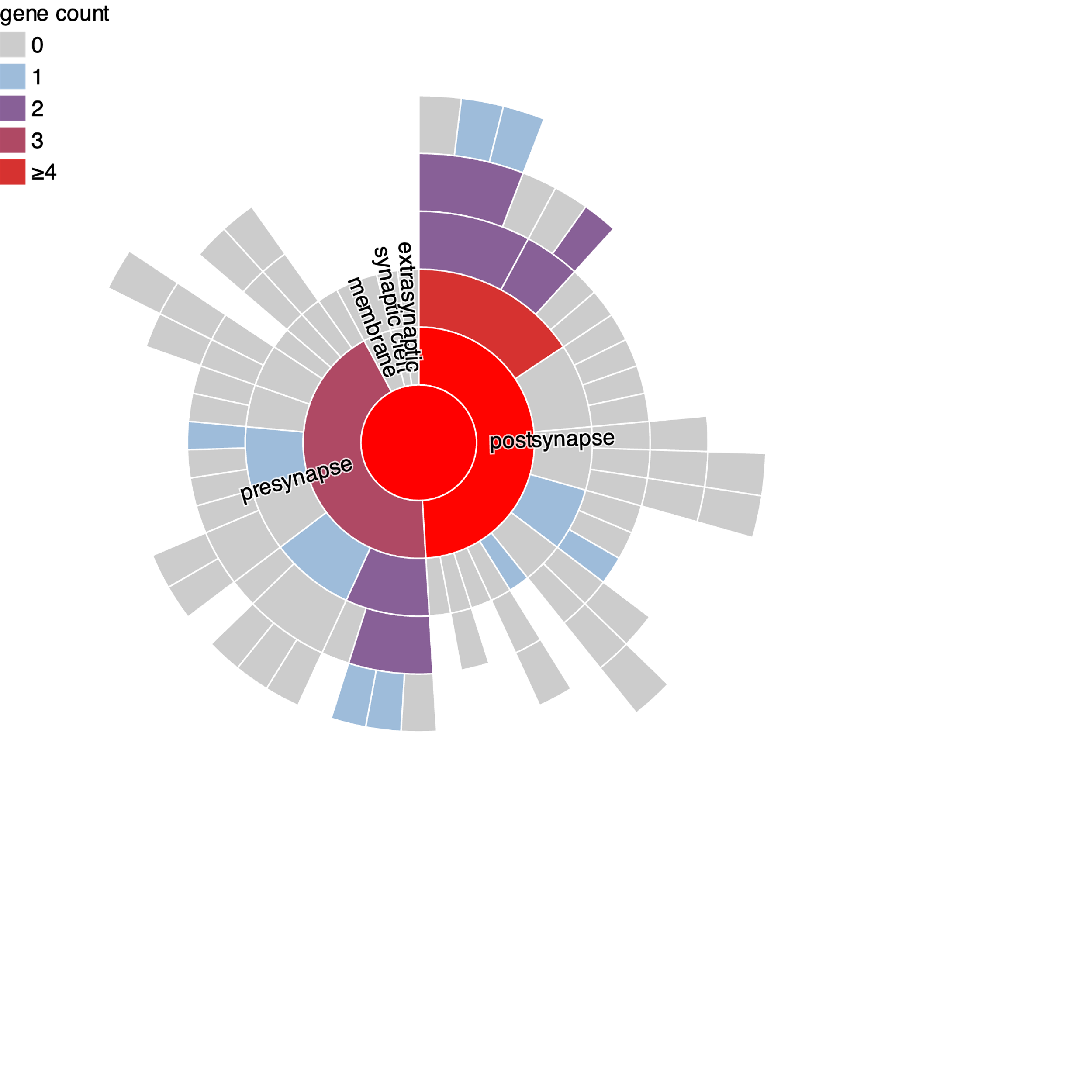

**Figure** **27:** Visualisation of the SynGO terms which are annotated by genes in the prescription-derived gene set, separated by presynaptic (left) or post synaptic (right) location (returned from SynGO web portal).

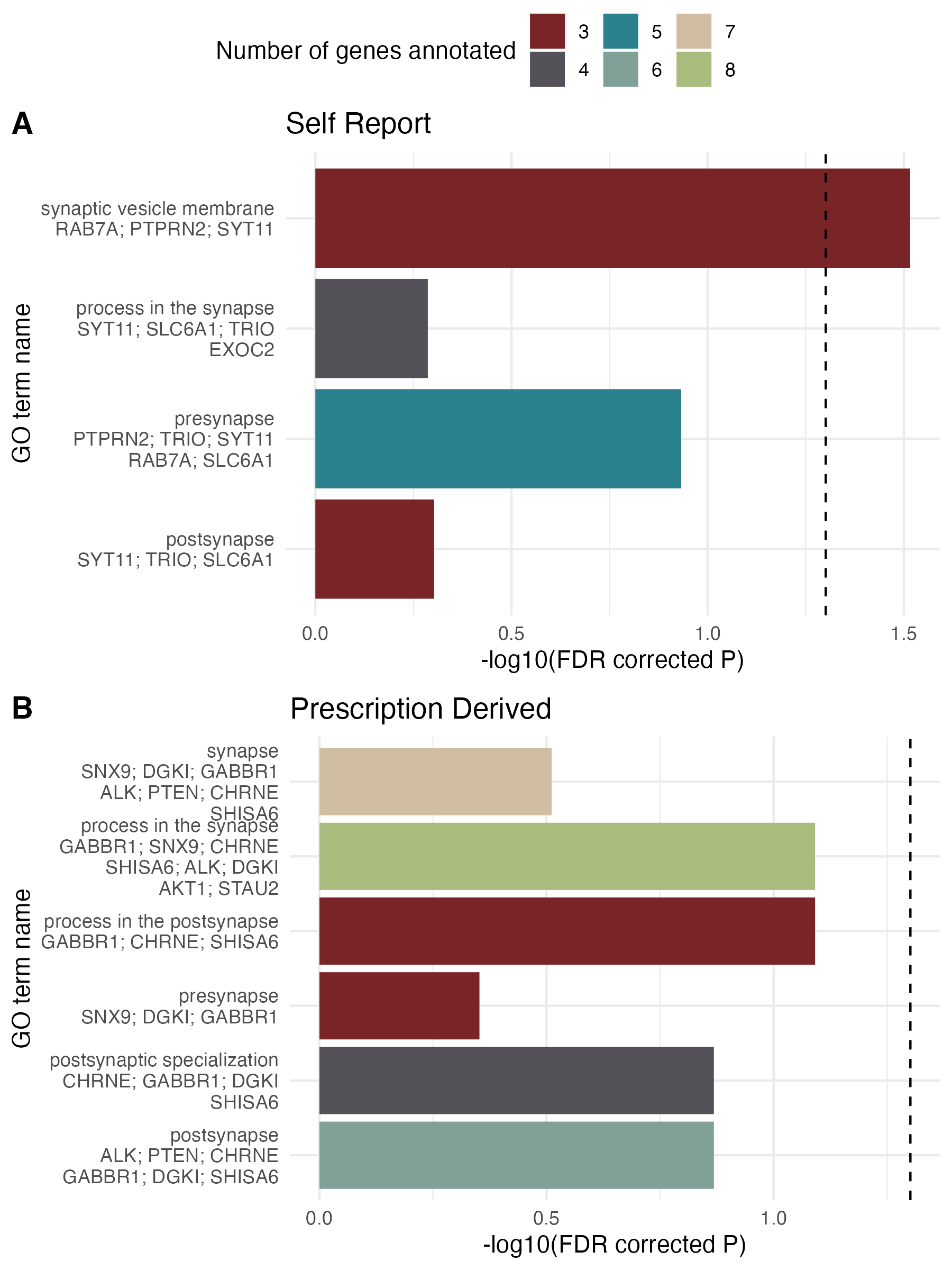

**Figure** **28:** Enrichment of SynGO terms in the self-report (top) and prescription-derived gene sets. The only term with significant enrichment was the synaptic vesicle membrane with the self-report gene set.

##### Msigdbr GO: Biological Processes

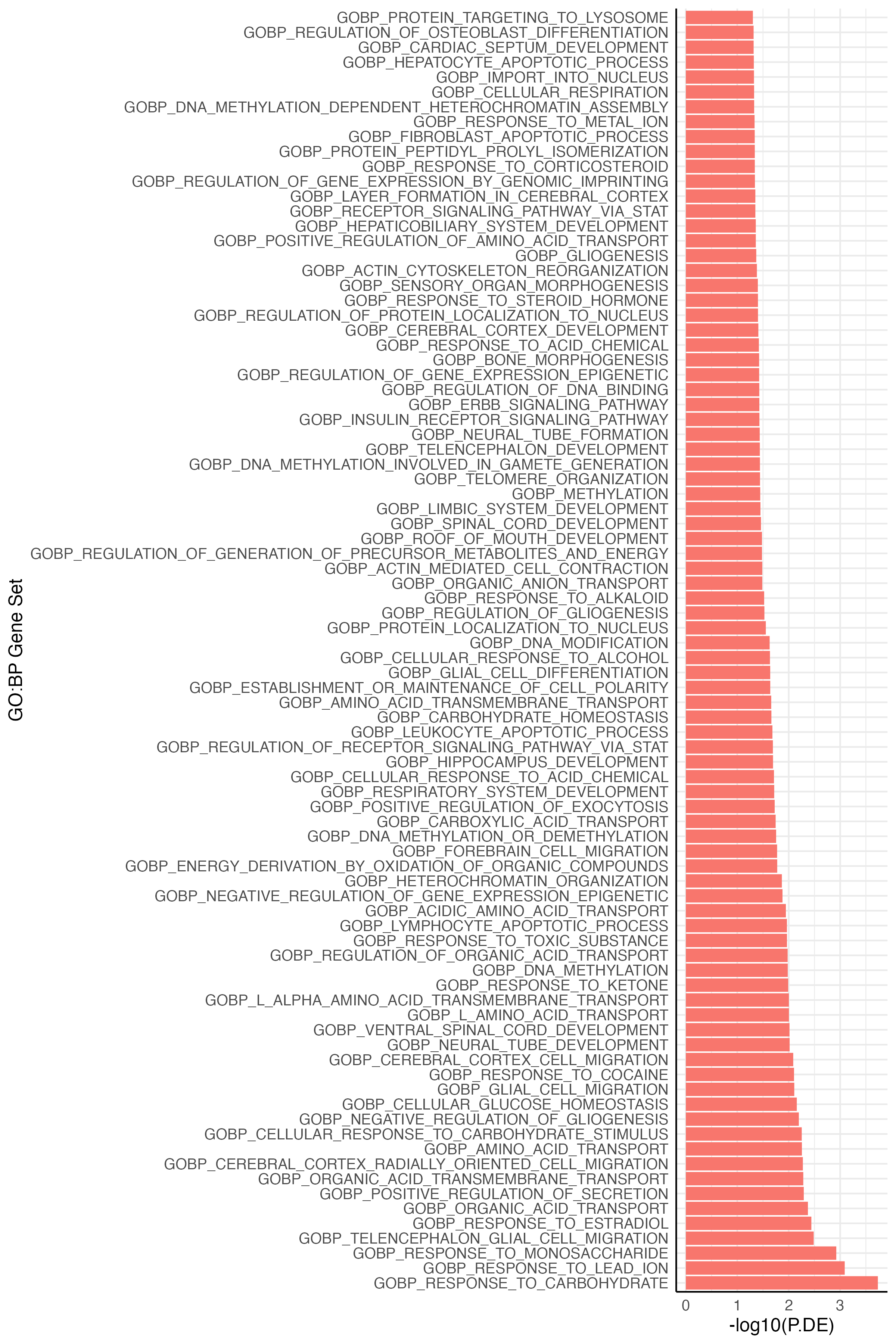

**Figure** **29:** GO: Biological Processes gene sets which were nominally enriched for the self-report gene set.

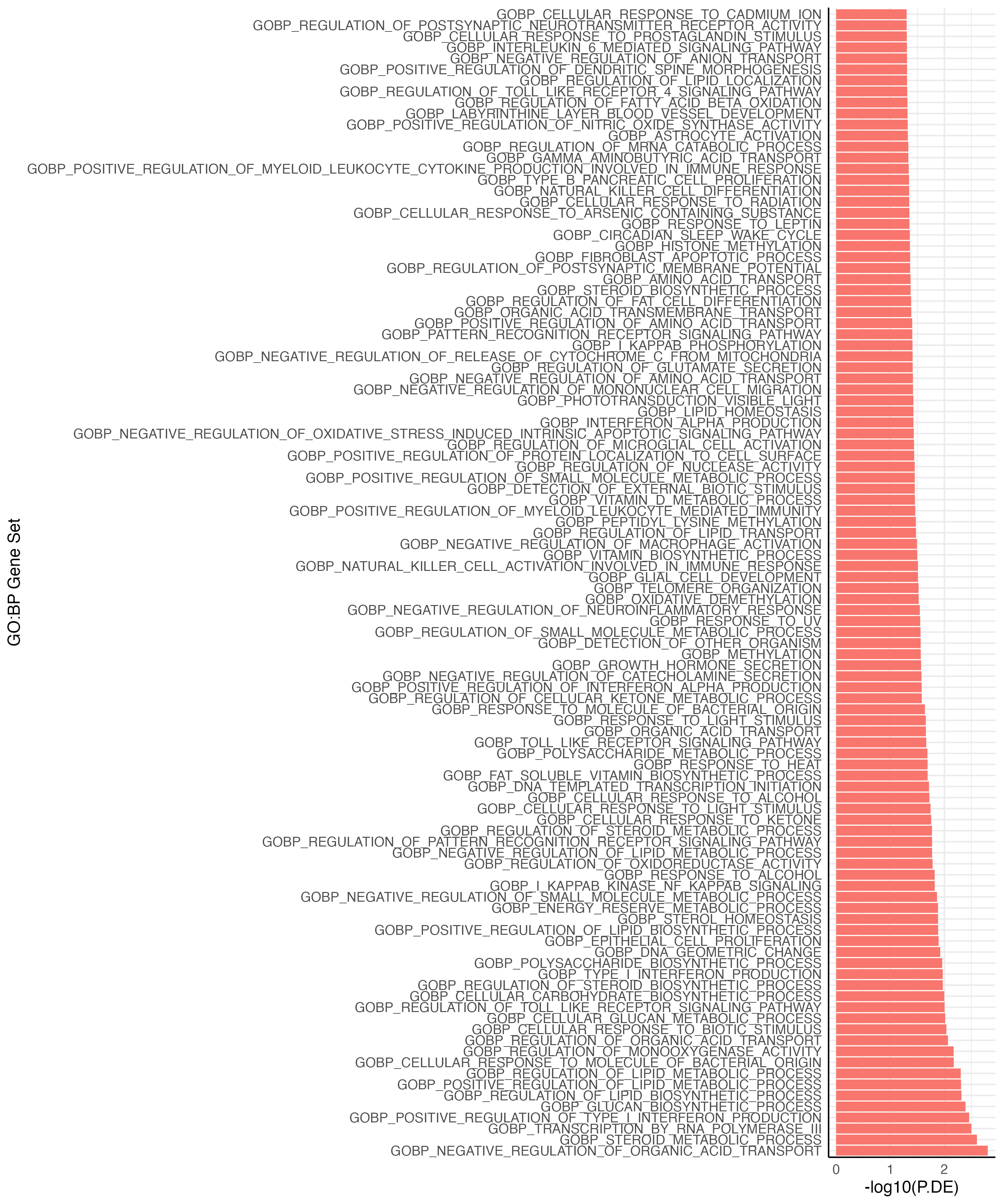

**Figure** **30:** GO: Biological Processes gene sets which were nominally enriched for the prescription-derived gene set.

#### Permutation-based enrichment analysis; Netherlands Study of Anxiety and Depression

Protocol paper: <https://doi.org/10.1002/mpr.256>^59^

##### Cohort Description

The Netherlands Study of Depression and Anxiety is a naturalistic ongoing longitudinal cohort study which aims to investigate the long-term course and consequences of depression and anxiety in the Netherlands. Recruitment took place in the general population across various health care settings (community, primary care and specialised mental health care) for individuals between 18 and 65 years of age. The recruited sample consists of 1,701 individuals with current diagnosis of depression and/or anxiety, 907 individuals with lifetime diagnoses or at risk due to family history of subthreshold symptoms and 373 healthy controls. A four-hour baseline assessment was conducted and included written questionnaires, interviews, a medical examination, cognitive tasks, blood and saliva sampling and intensive screening regarding mental health outcomes. Detailed assessments were then repeated after one, two, four and eight years of follow-up.

**Ethics Approval**: The study protocol was approved centrally by the Ethical Review Board of the VU University Medical Centre and subsequently by local review boards of each participating centre. After full verbal and written information about the study, written informed consent was obtained from all participants at the start of baseline assessment.

##### Depression status and antidepressant exposure phenotypes

Major Depressive Disorder (MDD) DSM-IV diagnoses were obtained using the Composite International Diagnostic Interview (CIDI). The present study only included persons who fulfilled the 6-month recency MDD diagnosis. During the assessment, participants were asked about current medication. The assignment of drugs to the antidepressant treatment group was based on drug container inspection of all medications used in the past month, classified according to the World Health Organization Anatomical Therapeutic Chemical (ATC) classification. We included three classes of antidepressants: selective serotonin reuptake inhibitors (SSR: ATC: N06AD), tricyclic antidepressants (TCA: ATC: N06AA) and other antidepressants (ATC: N05AX). Those who named any single medication were classed as antidepressant-exposed (n = 398), and those who did not use a single antidepressant medication were defined as antidepressant unexposed (n = 414).

##### DNA methylation: Methyl-CG binding domain sequencing (MBD-Seq) assay

DNA methylation was assayed at baseline from whole-blood samples from 812 MDD patients using an optimized protocol for methyl-CG binding domain sequencing (MBD-Seq). Elsewhere we summarized key features of optimized MBD-seq using empirical data^60^. Critically, comparisons with whole genome bisulfite sequencing (WGB-Seq) showed that our optimized MBD-Seq protocol achieves near complete coverage of all methylation sites in blood^62^. First, genomic DNA samples were sonicated to 150bp using a Covaris S2 ultra-sonicator. Next, DNA fragments with methylated CpGs (mCpG) were enriched using components of the MethylMinerTM Kit (Invitrogen) via affinity purification with the methyl-CG domain protein (MBD2). For each capture reaction, 15uL of prepared MBD-beads was incubated with 1.5ug of fragmented DNA for 1 hour at room temperature on an orbital shaker at 650rpm, and then washed three times with 1x Bind-Wash Buffer. The bound methylated fragments were recovered in three pooled elutions of 500mM NaCI buffer and purified by ethanol precipitation. The MBD enriched fractions were used to generate indexed libraries with the TruSeq Nano DNA HT Library Prep Kit (Illumina). Libraries were size-selected using SPRI beads to obtain a mean insert size of 150bp. The libraries were then pooled and sequenced on the NextSeq 500 using a 75-bp single-end configuration and High-Output v2 chemistry (Illumina). Reads were aligned (build hg19/GRCh37) with Bowtie2^63^ using a seed-and-extend approach combined with local alignment while allowing for gaps. Specifically, we used a 20bp seed with zero mismatches. Rather than considering the entire read, local alignment was used to improve sensitivity by finding the maximum similarity score between the reference sequence and a substring of the extension that may be “trimmed” at both ends. Gaps were allowed to account for small indels.

##### Quality control of MBD-Seq data

Data was further processed using *RaMWAS*, an R/Bioconductor pipeline for large-scale analyses of methylation data from enrichment platforms^64^. The code and software are available from Bioconductor (<https://bioconductor.org/packages/release/bioc/html/ramwas.html>) as well as GitHub (<https://github.com/andreyshabalin/>). Reads aligning to loci without CpGs (non-CpGs) represent “noise” caused by, for example, imperfect enrichment leading to non-methylated fragments being sequenced. We use a threshold of 0.05 for non-CpG/CpG coverage ratio to remove samples with high-noise levels, leaving an average of 0.010 (SD = 0.005) in the remaining samples. Samples were also excluded if the sequence variants called from the methylation data did not match the genotype information, as this indicates that perhaps a sample swap or sample contamination occurred. Multidimensional outliers were identified, using principal components of the methylation dataset using the function *“pcout”* from the *mvoutliers* R package^65^ and excluded. The mean number of reads per subject was 59,758,680 and the average alignment rate was 99.1%. Quality control steps were performed for both multi- and duplicate-reads. For multi-reads, i.e., when a read aligns to multiple locations of the genome, Bowtie2 selected the alignment with a highest alignment score. In the case where Bowtie2 encounters a set of equally good alignment it uses a pseudo-random number to select one primary alignment. Duplicate-reads, i.e., multiple identical reads with the same start and end location, are more likely to occur when sequencing enriched genomic fractions. Therefore, we allow up to three duplicate-reads at each genomic position. In the instance where there are > 3 duplicate reads, read count is reset to 1, implicitly assuming that excess reads are tagging a single clonal fragment. This results in an average of 48,287,403 reads per sample (81.6% of all reads).

##### Identifying CpGs and quantifying DNAm

We combined the reference genome sequence (hg19/GRCh37) with common SNPs calculated on the European super population from the 1000 Genomes (Phase 3). CpGs which were 1) created by SNPs with minor allele frequency < 1%, 2) in loci prone to alignment errors (identified using RaMWAS in-silico alignment experiment)^66^ and 3) with an average coverage of < 0.1 or having 0 coverage in > 90% of samples were removed. This left a total of 26,269,235 CpGs for analysis. CpGs methylation were quantified using RaMWAS in a three-step process. First, a non-parametric approach is used to estimate the fragment size distribution from the sequencing data using isolated CpGs^67^. This distribution is then used to calculate the probability that a sequenced fragment will cover the CpG under consideration. The CpG score is then calculated by taking the sum of probabilities for all fragments aligning within proximity of the CpG.

##### Enrichment analysis

We tested whether top findings from the case-control methylome-wide association studies (MWAS) in Generation Scotland (GS) were also more likely to be among the top findings of the MWAS of antidepressant exposure in NESDA MDD cases. For this purpose, we created for both MWASs a yes/no variable indicating whether sites were in the top X percentage, using three different thresholds (0.1, 0.5 and 1%). As CpG methylation tends to be highly correlated over short distances^60^, we allowed for a flank of 150bp on either site of the CpG in the more sparse GS methylation array data when mapping results from the two MWASs. Next, we cross-classified the two yes/no variables resulting in a 2 by 2 table. The tests for enrichment in overlap were performed using Cramér’s V (sometimes referred to as Cramér’s phi). We chose Cramér’s V over a variety of other association statistics for 2 by 2 table based on a simulations study showing it resulted in the most powerful test. For the analyses we used circular permutations^68^ to account for having correlated methylation levels between CpG sites. Previous studies have shown that circular permutations properly control the type I error even if correlations between methylation sites were > 0.99^69^. A total of 100,000 permutations were performed were performed with the help of our R package shiftR (<https://cran.r-project.org/web/packages/shiftR/index.html>) that performs permutations through fast bitwise operations. P values were calculated as the proportion of permutations that yielded a value equal of greater that Cramér’s V observed in the analysis of the observed data. Three thresholds (0.1, 0.5 and 1%) were used to define top findings in the MWASs. We corrected for this “multiple testing” by using the same thresholds in the permutations and then selected the most significant result for each permutation.

##### Acknowledgements

The NESDA study is supported by the Geestkracht program of the Netherlands Organization for Health Research and Development (Zon-Mw, grant number 10–000-1002) and the participating institutions (VU University Medical Center, Leiden University Medical Center, University Medical Center Groningen. The current methylation project was supported by grant R01MH099110 from the National Institute of Mental Health. The sponsors had no role in the design and conduct of the study; collection, management, analysis, and interpretation of the data; preparation, review, or approval of the manuscript; or decision to submit the manuscript for publication.

#### Correlation of methylation and time in treatment periods

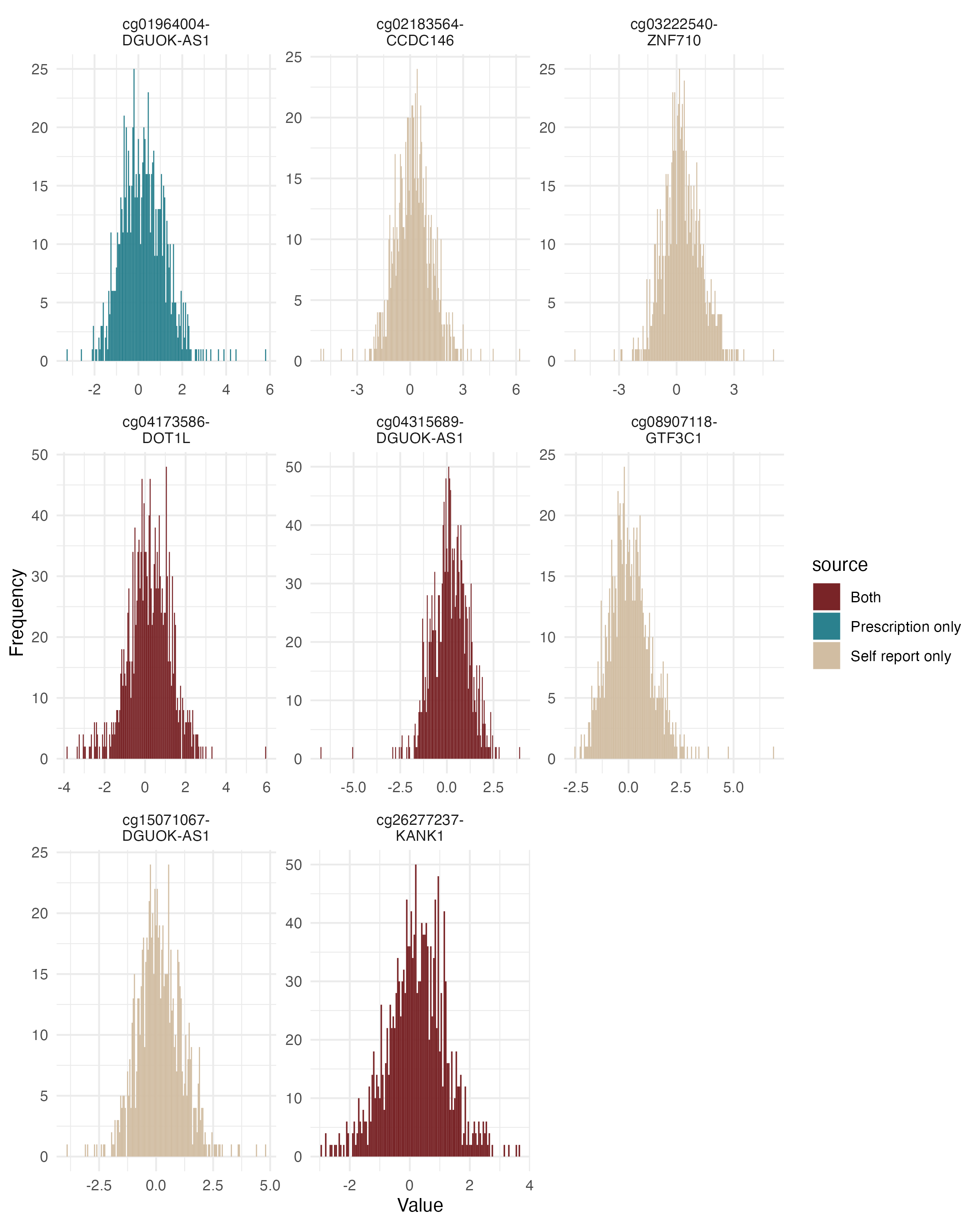

**Figure** **31:** Histograms of the methylation levels of probes significantly associated with antidepressant exposure (either self-report and/or prescription-derived measures), in those in a treatment period at the time of blood draw. Colour of the bars denotes whether the CpG was significant in the self-report MWAS, prescription-derived MWAS or both.

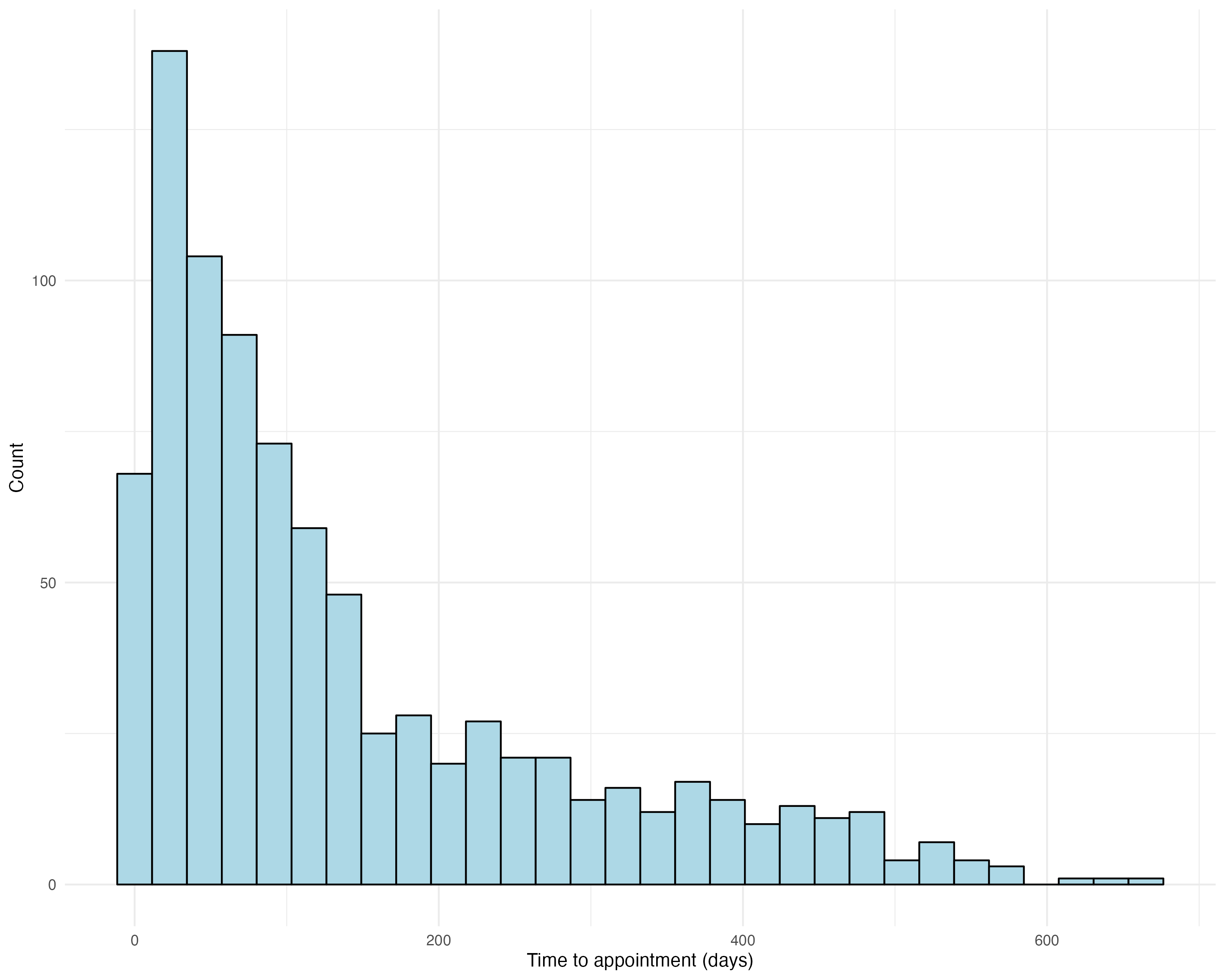

**Figure** **32:** Histogram of the time between the beginning of a current treatment period and the blood draw appointment (DNAm measurement), for those within a treatment period at the time of the blood draw.

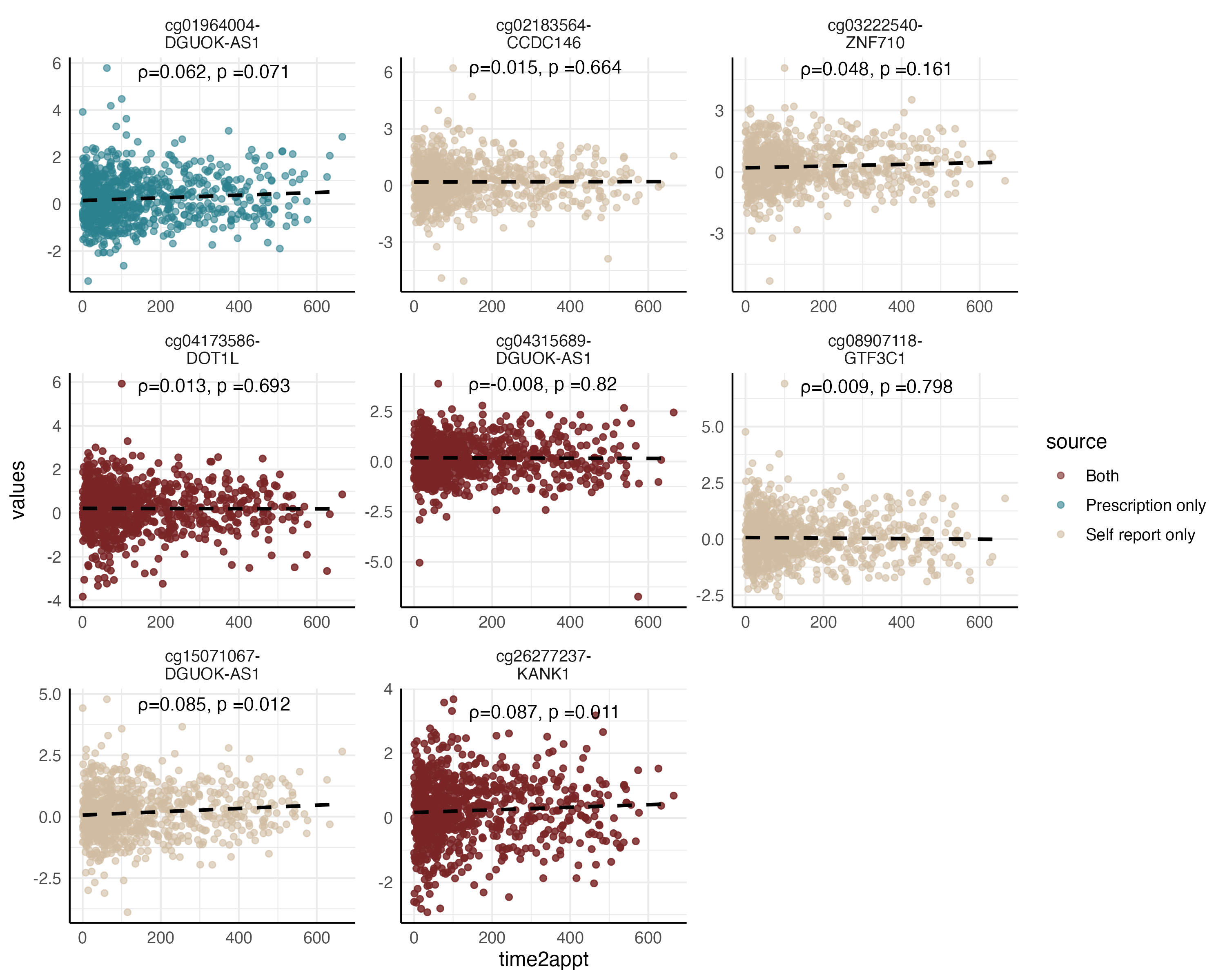

**Figure** **33:** Methylation at significant probes identified from self-report and prescription-derived AD MWAS and the time in treatment prior to the blood draw. Spearman correlation tests were performed, with rho and p-values printed on each graph. Colour of the dots denotes whether the CpG was significant in the self-report MWAS, prescription-derived MWAS or both.

#### Methylation Profile Score

##### LASSO training model

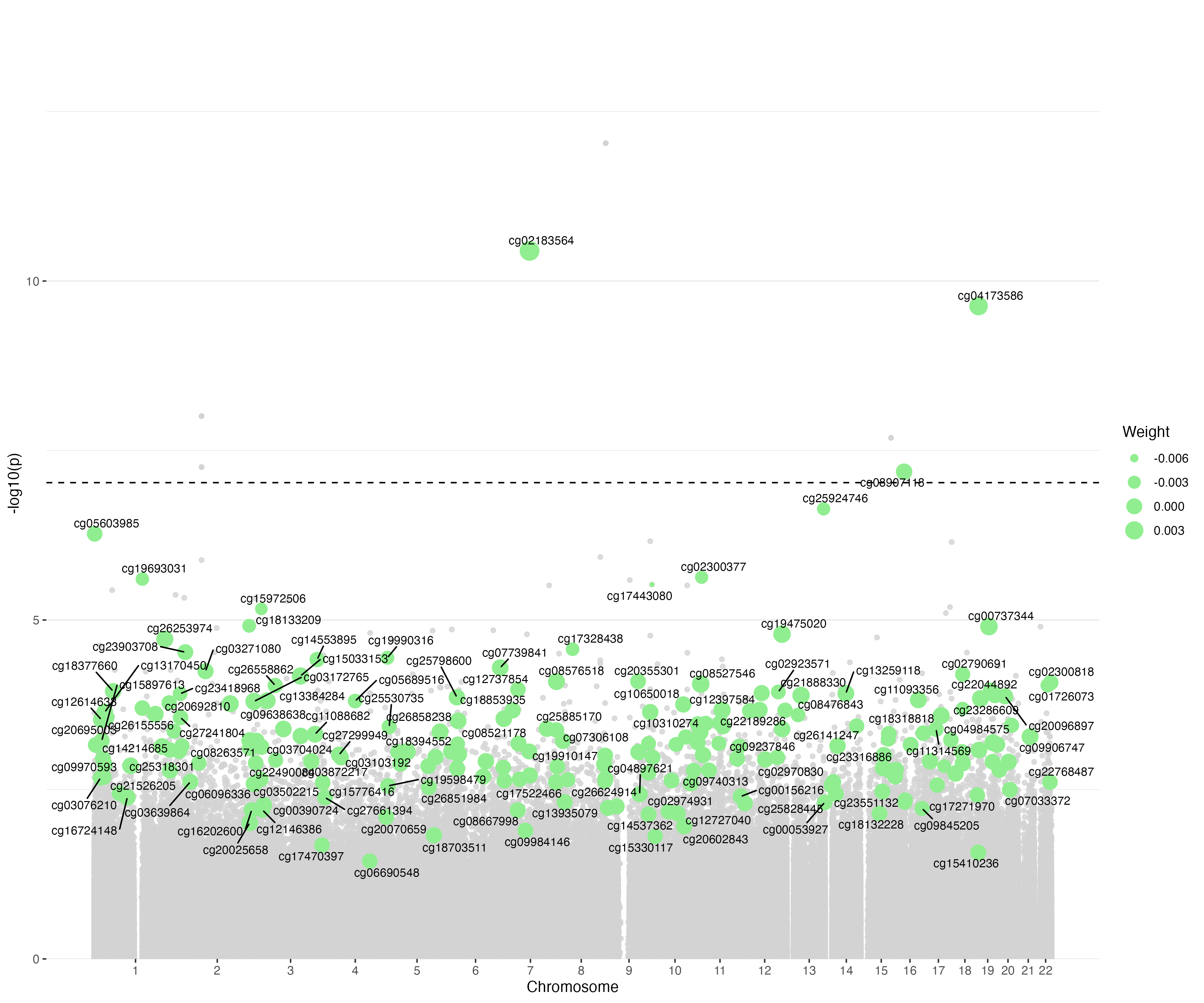

**Figure** **34:** The self-reported antidepressant use MWAS manhattan plot with the 212 CpGs with a non-zero weight from the antidepressant exposure ~ DNAm LASSO model in Generation Scotland highlighted in green and labelled.

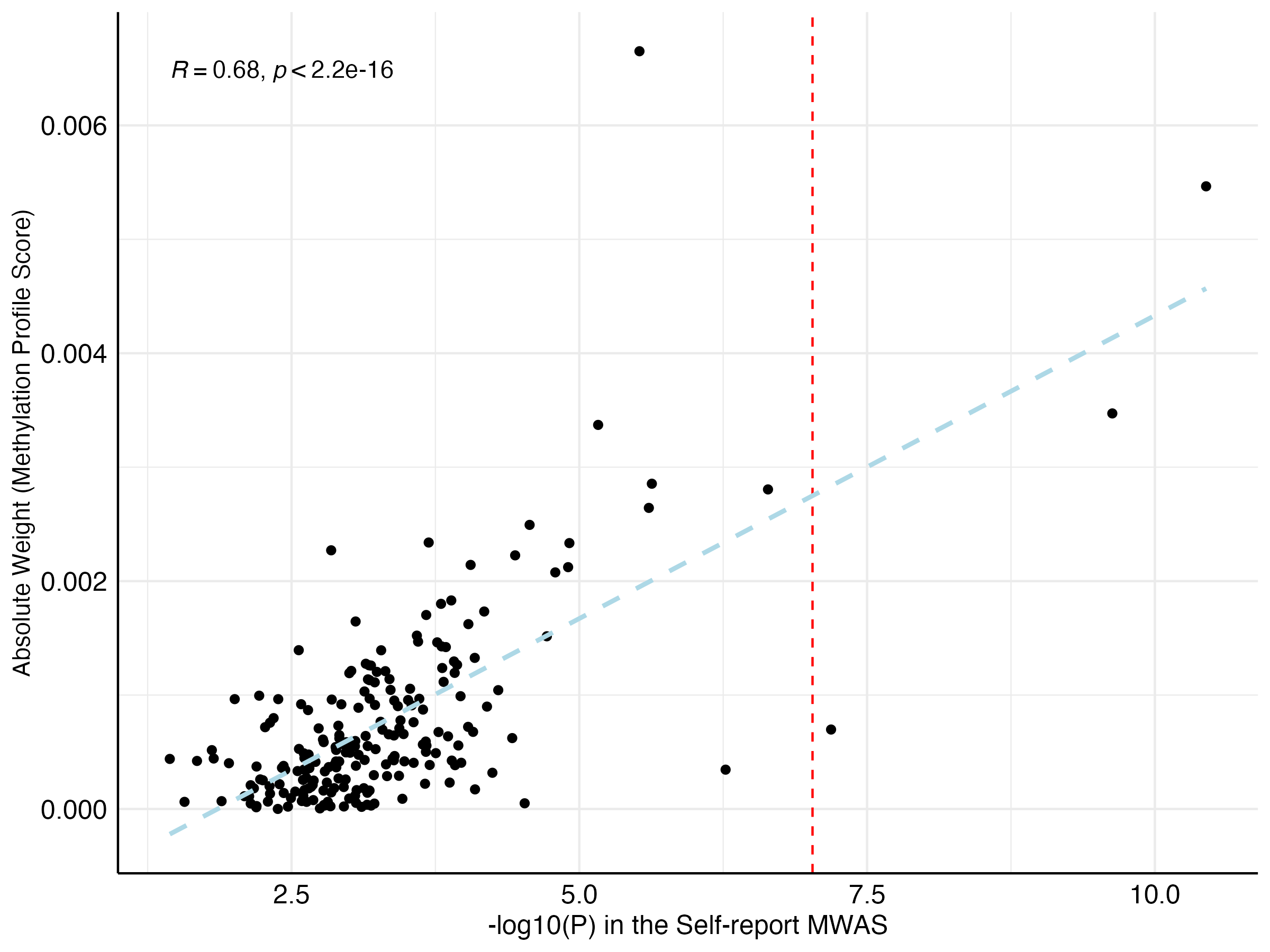

**Figure** **35:** The -log10(p) from the self-reported MWAS and the weights assigned to the CpG in the LASSO regression for the 212 CpGs included in the methylation profile score. The red dashed line denotes the significance threshold (9.42x10^-08^) in the MWAS, and the light blue dashed line is the regression line between the two variables.

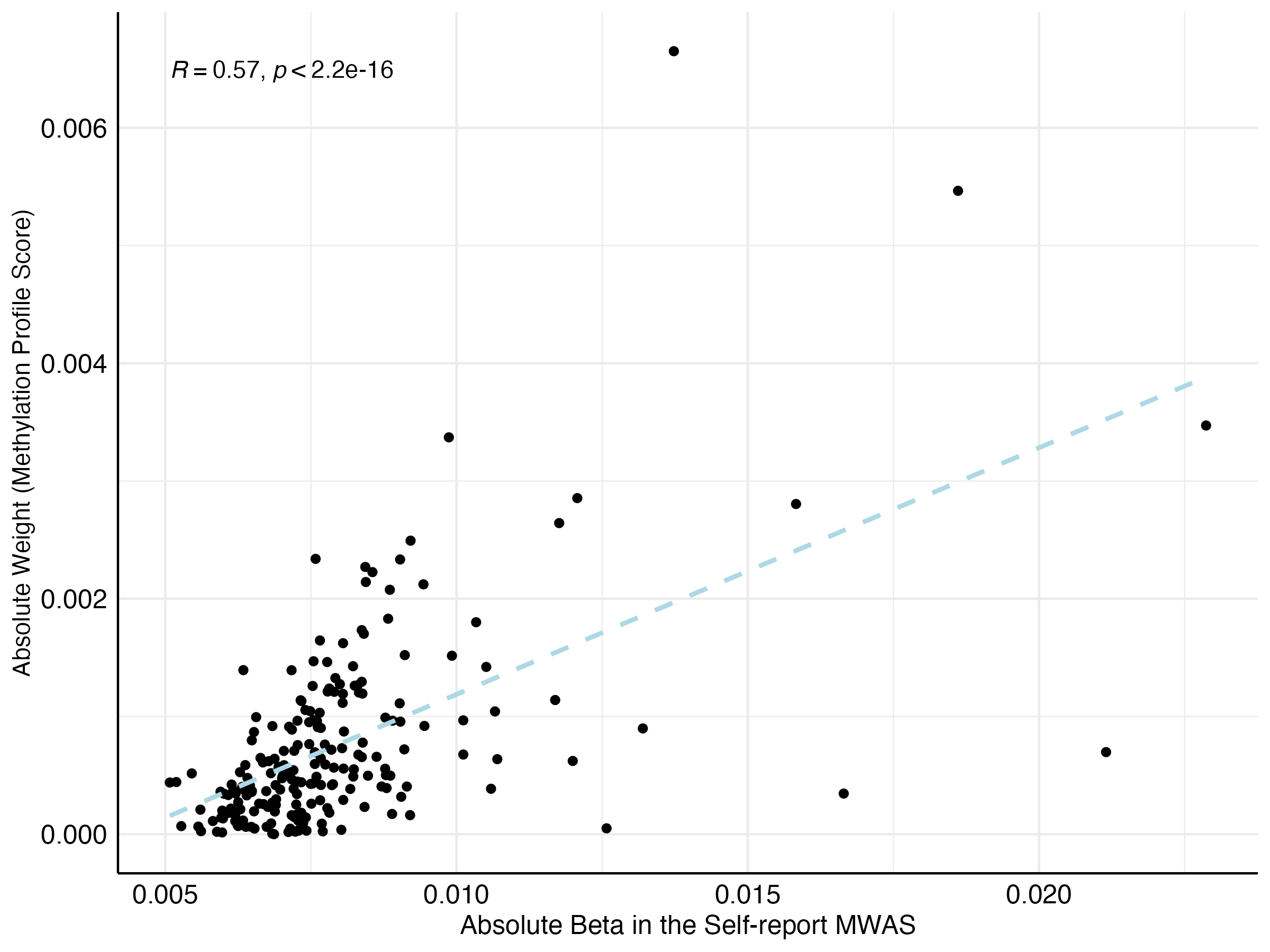

**Figure** **36:** The beta effect sizes from the self-report MWAS and the weights assigned to the CpG in the LASSO regression for the 212 CpGs included in the methylation profile score. The light blue dashed line denotes the regression line between the two variables.

#### MPS calculation in external cohorts

##### Finn Twin Cohort

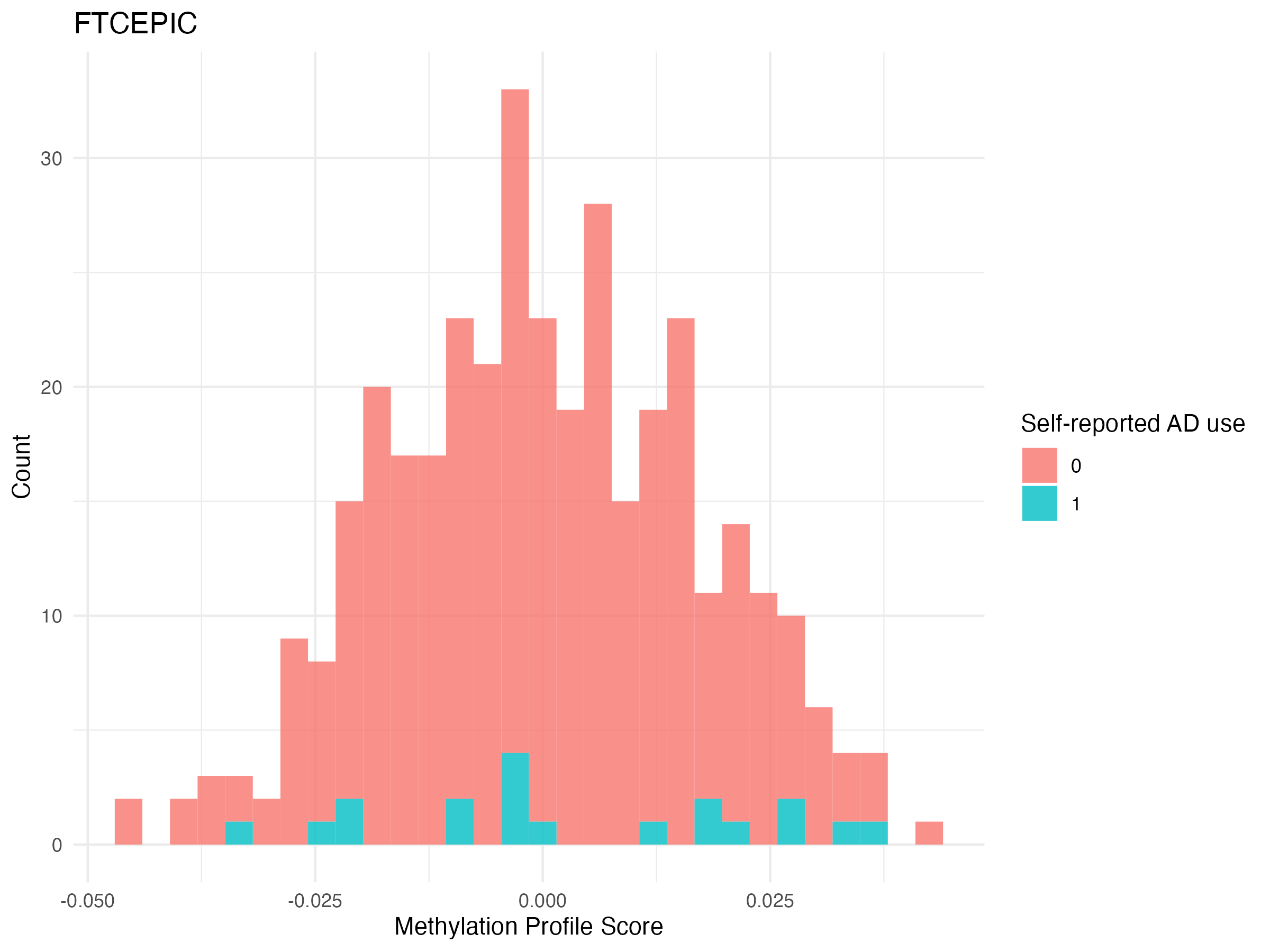

**Figure** **37:** The distribution of antidepressant exposure MPS in antidepressant-exposed and antidepressant-unexposed individuals in FinnTwin12 & FinnTwin16 (which used Illumina EPIC array) (n_exposed_ = 19, n_unexposed_ = 344)

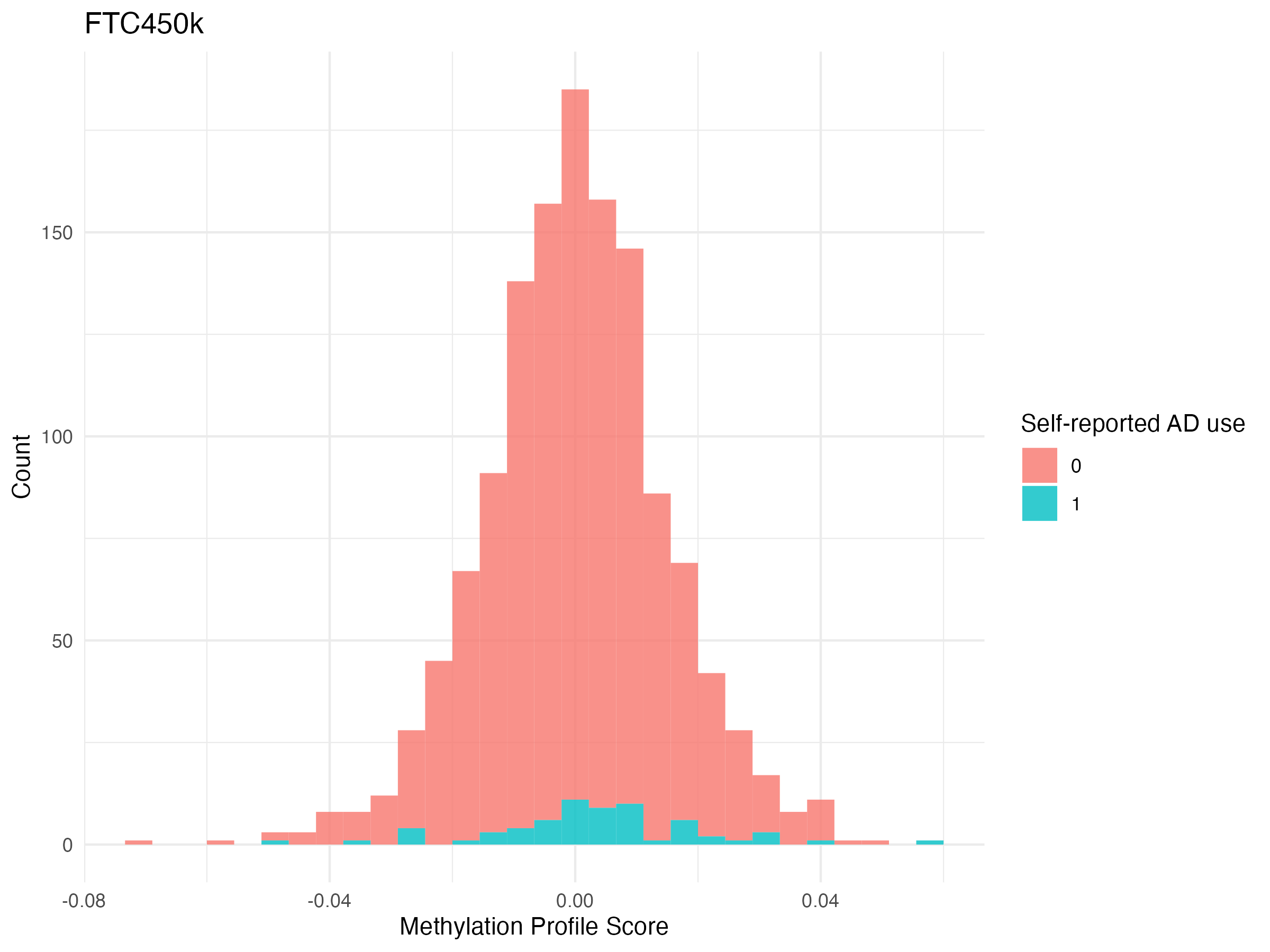

**Figure** **38:** The distribution of antidepressant exposure MPS in antidepressant-exposed and antidepressant-unexposed in the Older Twin Cohort (which used Illumina 450K array) (n_exposed_ = 65, n_unexposed_ = 1250)

##### SHIP-TREND

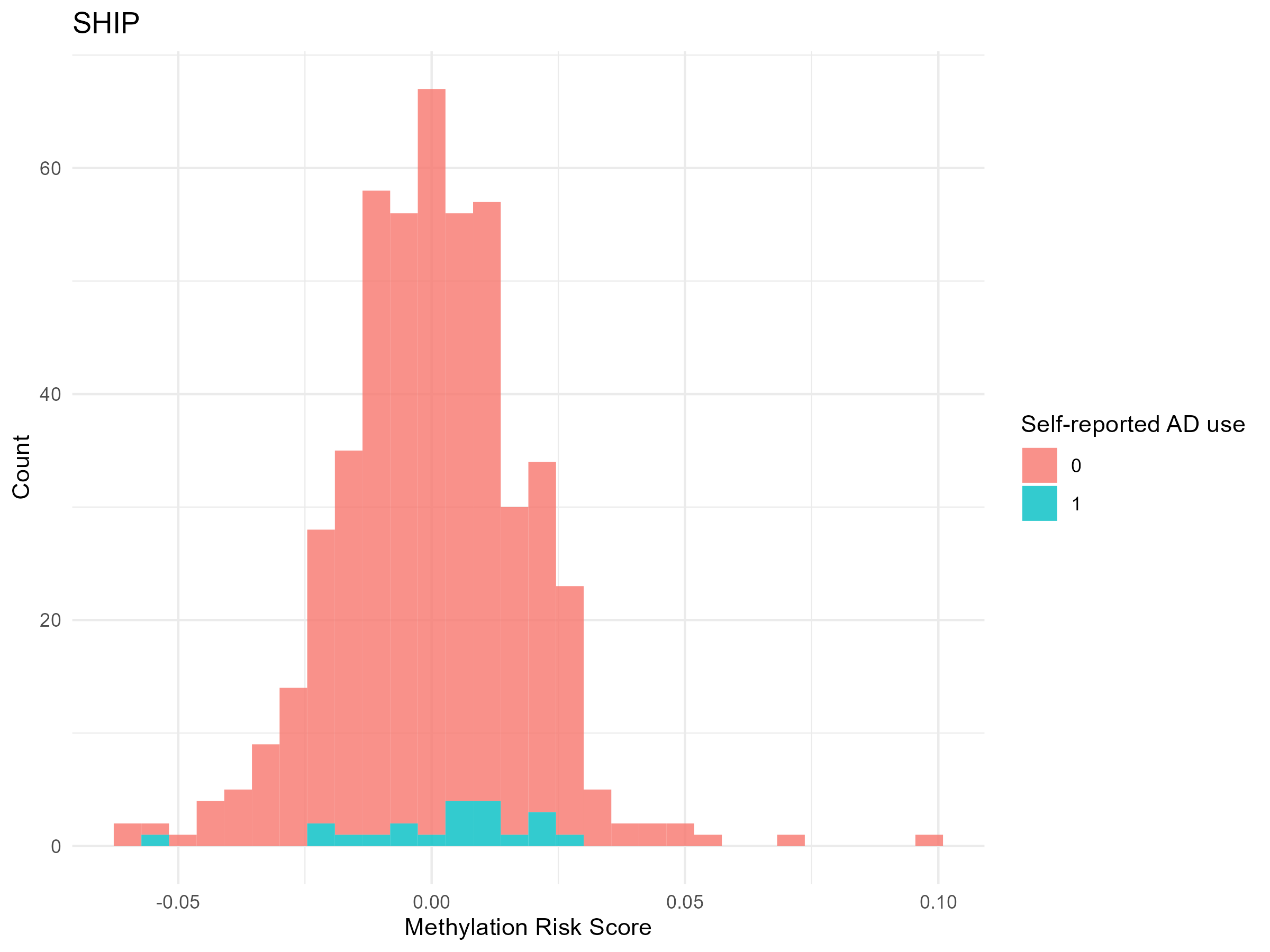

**Figure** **39:** The distribution of antidepressant exposure MPS in antidepressant-exposed and antidepressant-unexposed in the SHIP-Trend Cohort (n_exposed_ = 21, n_unexposed_ = 474).

##### FOR2107

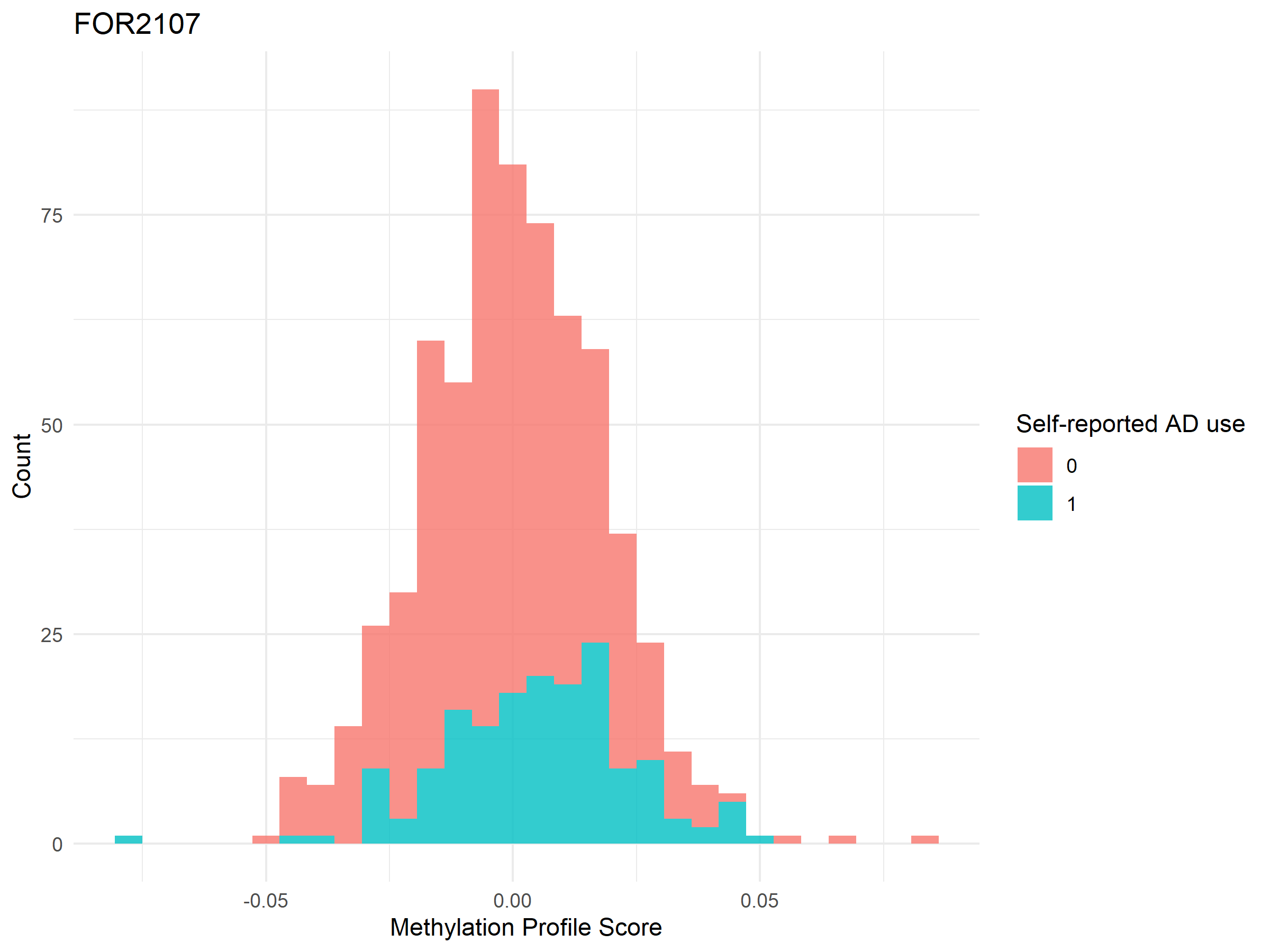

**Figure** **40:** The distribution of antidepressant exposure MPS in antidepressant-exposed and antidepressant-unexposed in the FOR2107 cohort (n_exposed_ = 165, n_unexposed_ = 493).

##### NTR

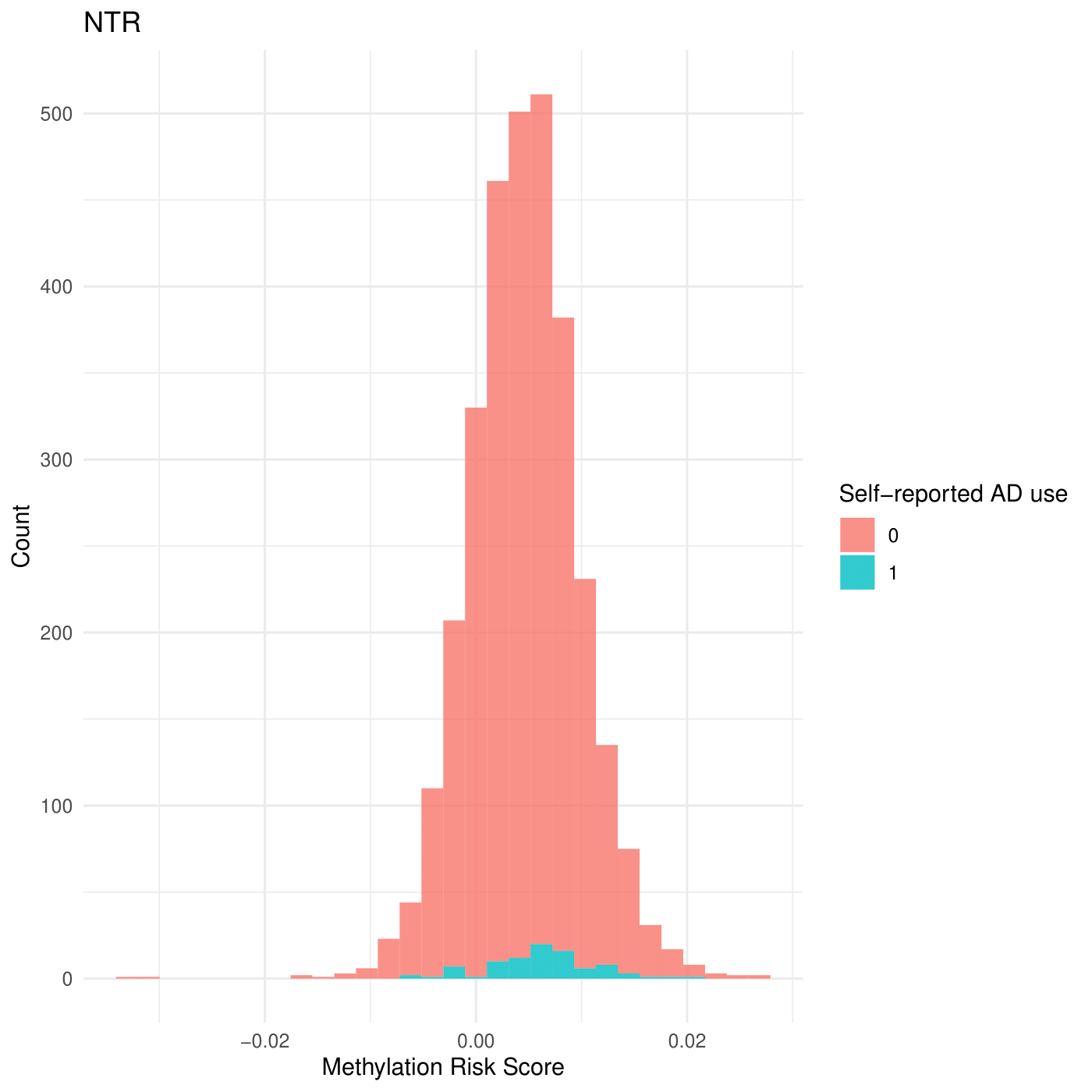

**Figure** **41:** The distribution of antidepressant exposure MPS in antidepressant-exposed and antidepressant-unexposed in the NTR cohort (n_exposed_ = 89, n_unexposed_ = 2998).

##### MARS-UniDep

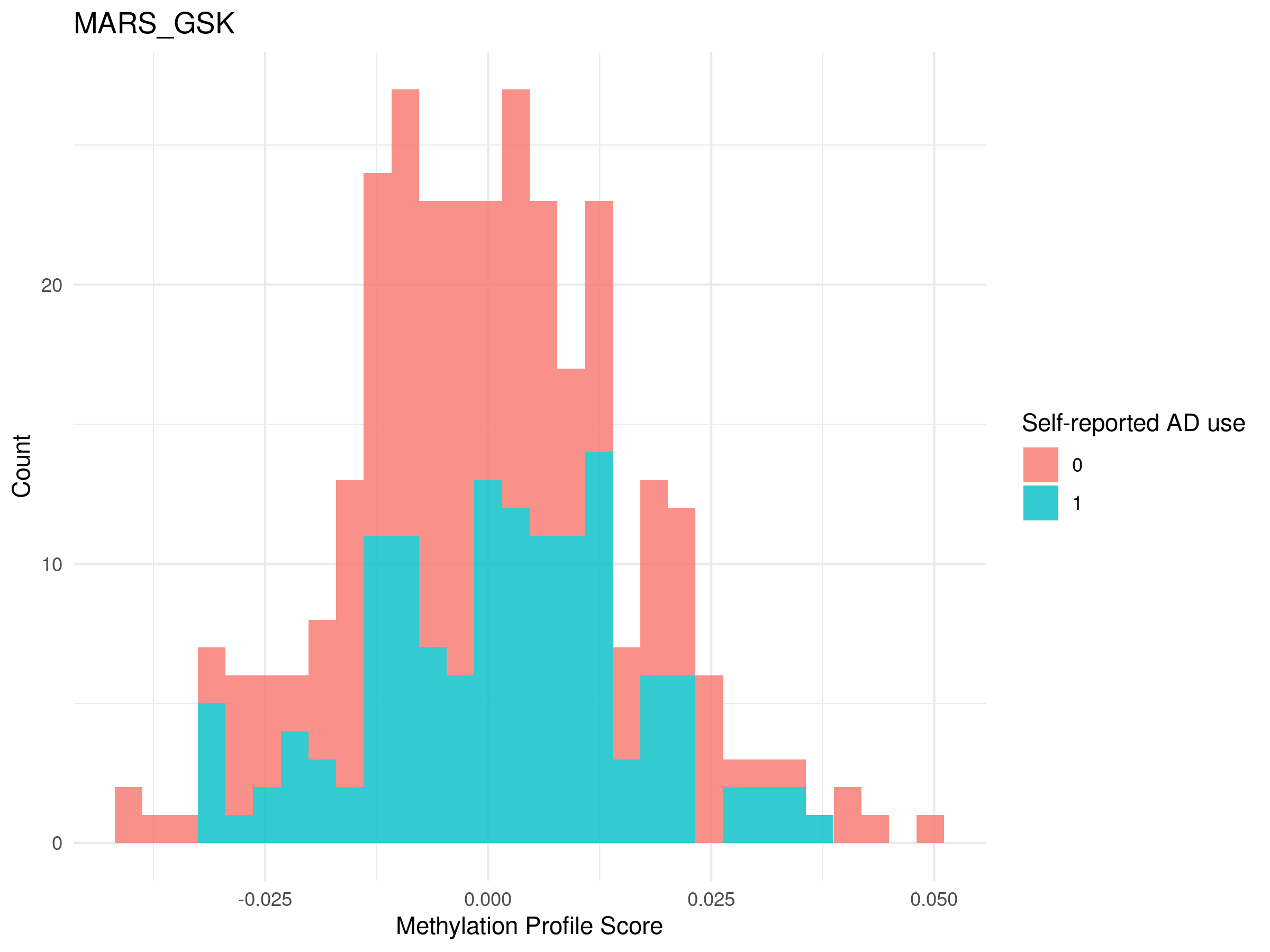

**Figure** **42:** The distribution of antidepressant exposure MPS in antidepressant-exposed and antidepressant-unexposed in the MARS-UniDep Cohort (n_exposed_ = 135, n_unexposed_ = 177). NB: The UniDep cohort is sometimes referred to as ‘GSK’.

##### LBC1936

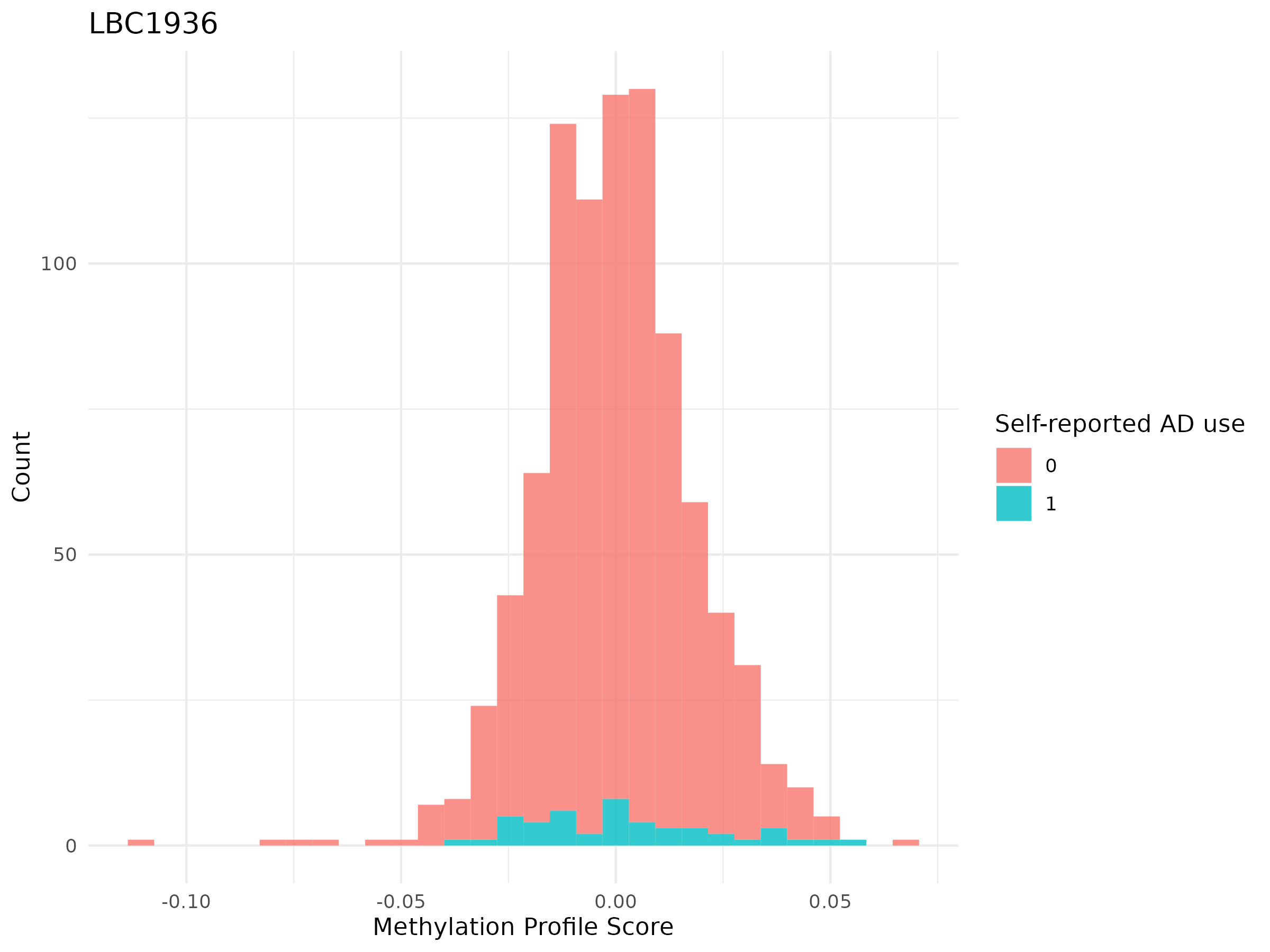

**Figure** **43:** The distribution of antidepressant exposure MPS in antidepressant-exposed and antidepressant-unexposed in the LBC1936 Cohort (n_exposed_ = 46, n_unexposed_ = 843).

##### ALSPAC

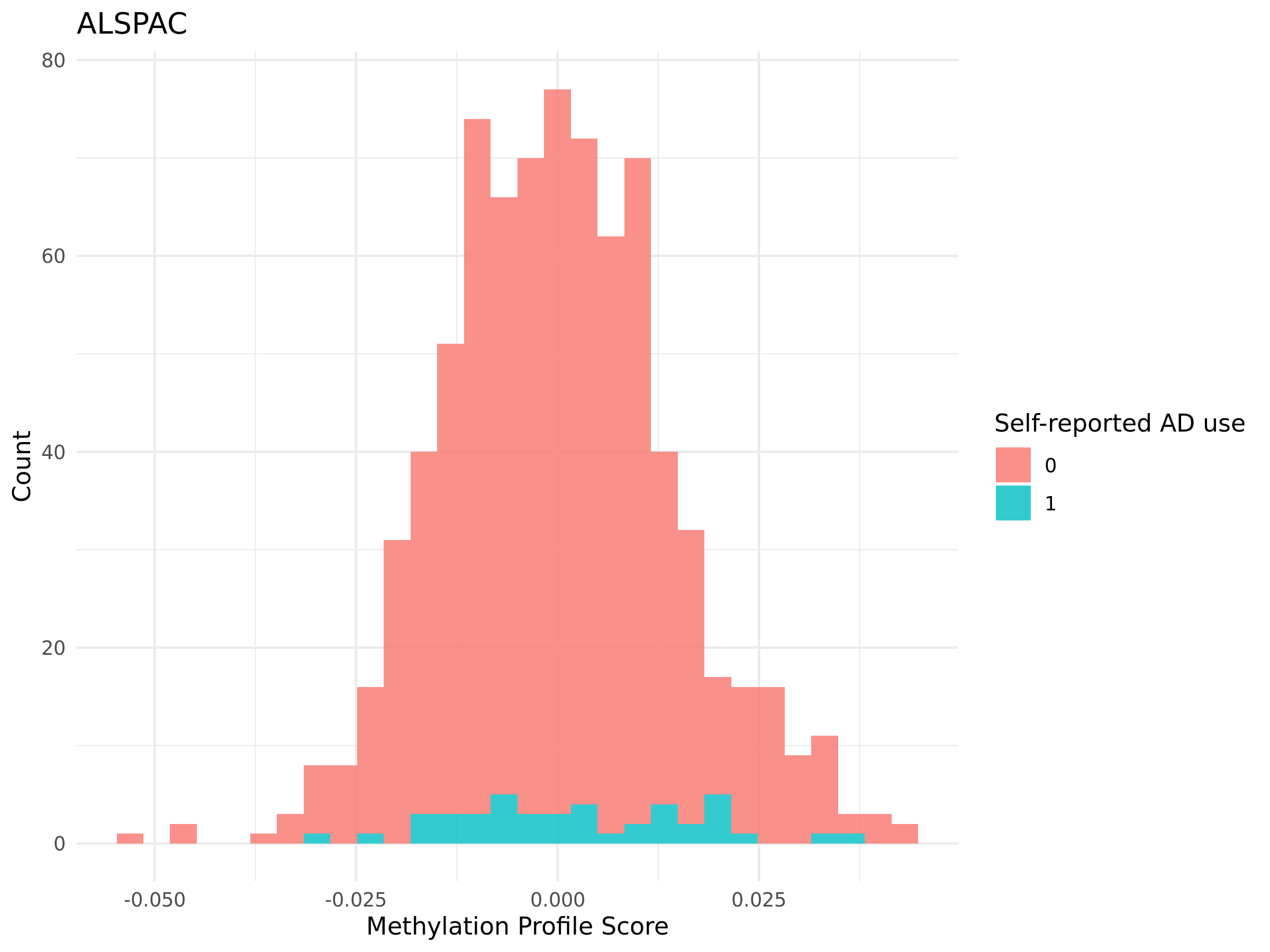

**Figure** **44:** The distribution of antidepressant exposure MPS in antidepressant-exposed and antidepressant-unexposed in the ALSPAC Cohort (n_exposed_ = 43, n_unexposed_ = 758).

##### ERISK

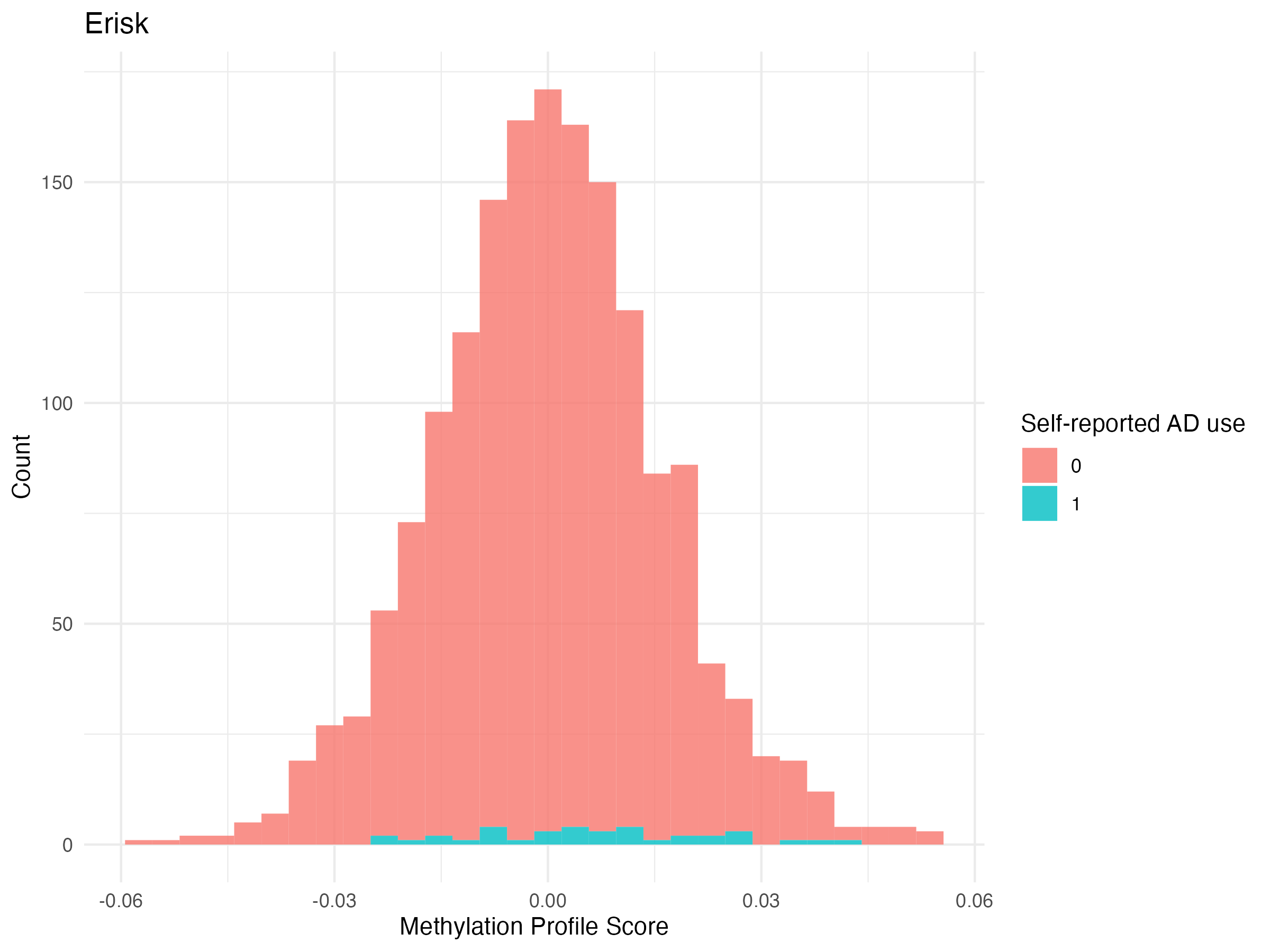

**Figure** **45:** The distribution of antidepressant exposure MPS in antidepressant-exposed and antidepressant-unexposed in the E-Risk Cohort (n_exposed_ = 36, n_unexposed_ = 1622).

#### MPS calculation in prospective Generation Scotland follow up

##### STRADL

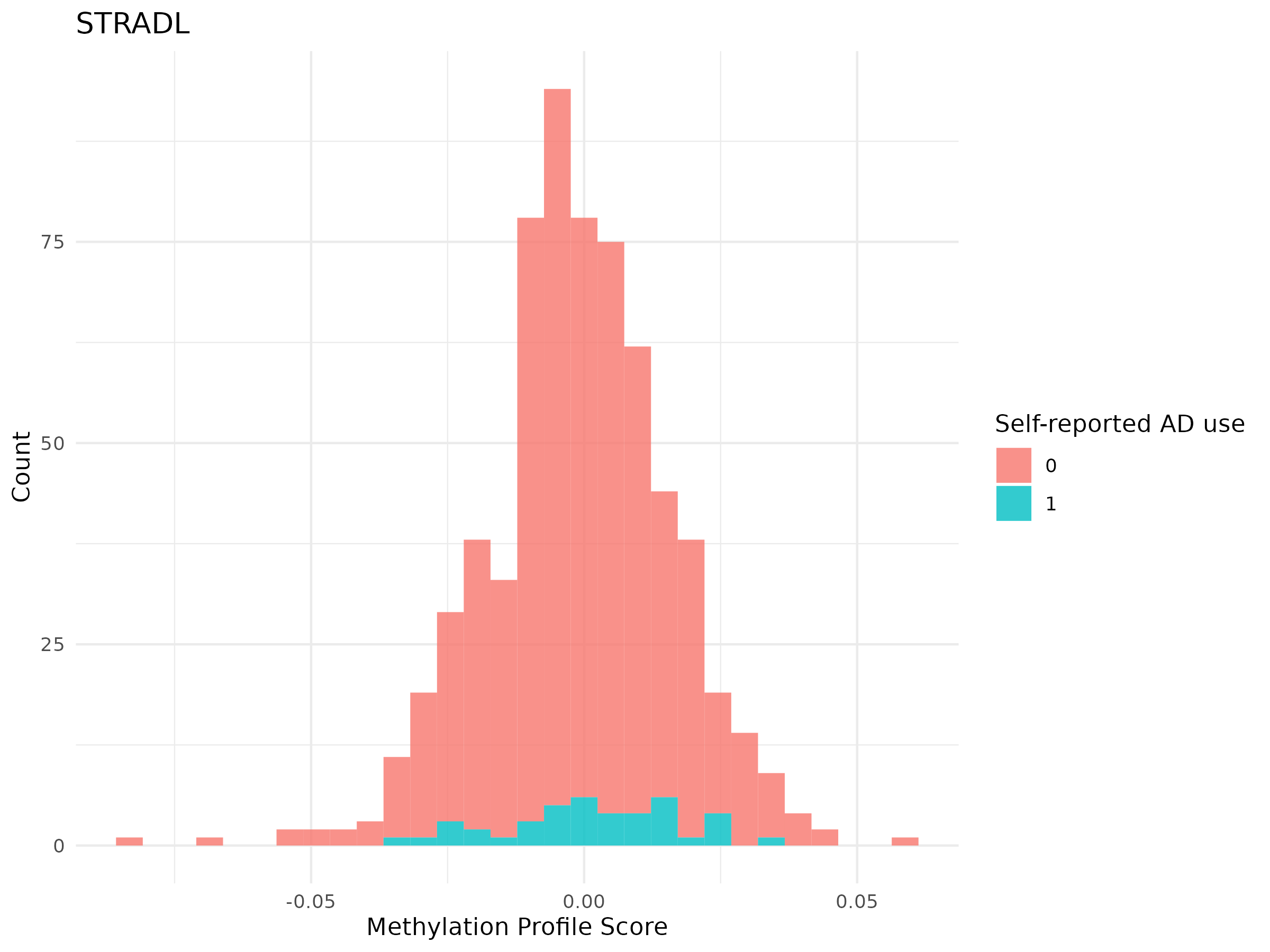

**Figure** **46:** The distribution of antidepressant exposure MPS in antidepressant-exposed and antidepressant-unexposed in the STRADL subcohort (n_exposed_ = 42, n_unexposed_ = 617). Note that this is a prospective subcohort of Generation Scotland (GS) at a 5-year follow-up. All individuals self-reported not taking antidepressants at baseline (GS) timepoint, and those who self-reported to taking antidepressants at the STRADL time point are classed as antidepressant-exposed and those who remain self-reporting no antidepressant exposure are classed as antidepressant-unexposed.

#### Antidepressant exposure ~ MPS associational models

##### Singular cohort results

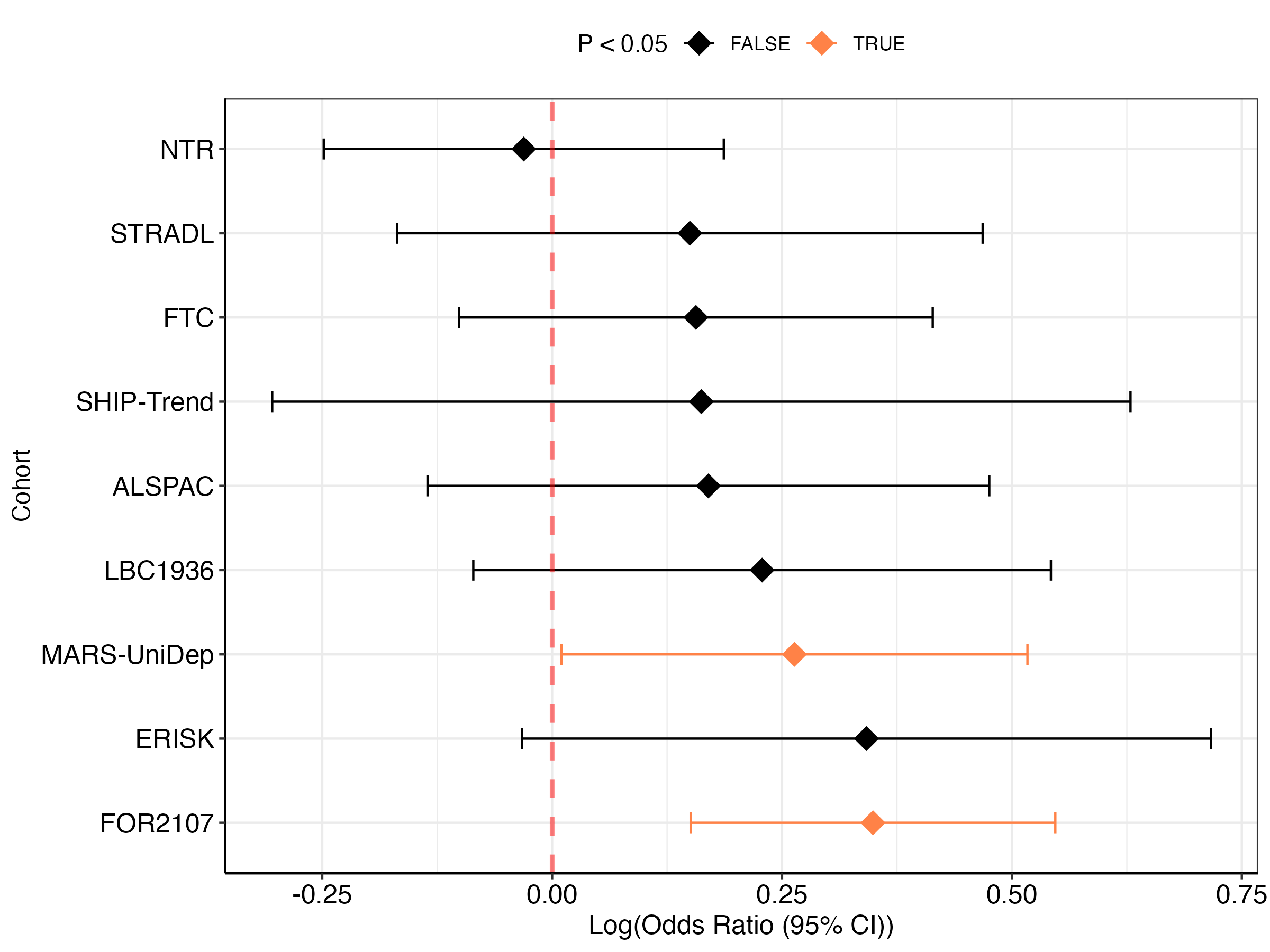

**Figure** **47:** The log(Odds Ratio) of antidepressant MPS association with antidepressant exposure in external cohorts.
