## Supplementary material for "Antidepressant Exposure and DNA Methylation: Insights from a Methylome-Wide Association Study": Tables

**Table 1 | Demographics of antidepressant exposed and unexposed individuals using the prescription-derived and self-reported antidepressant exposure phenotypes in Generation Scotland.** M = Mean, SD = Standard Deviation.

|  | Prescription-derived phenotype | | Self-report phenotype | |
| --- | --- | --- | --- | --- |
|  | Unexposed | Exposed | Unexposed | Exposed |
| N | 7090 | 861 | 15028 | 1508 |
| Age: M (SD) | 47.6 (15.2) | 51.1 (12.7) | 46.6 (14.9) | 50.2 (12.4) |
| BMI: M (SD) | 26.4 (4.78) | 28.5 (6.11) | 26.4 (5.01) | 28.2 (6.05) |
| Sex | | | | |
| Female: N (%) | 3746 (53) | 631 (73) | 8552 (57) | 1151 (76) |
| Male: N (%) | 3342 (47) | 230 (27) | 6464 (43) | 354 (23) |
| Smoking behaviours | | | | |
| Current: N (%) | 963 (14) | 214 (25) | 2448 (16) | 402 (27) |
| Former: N (%) | 1942 (27) | 273 (32) | 4183 (28) | 481 (32) |
| Never: N (%) | 3913 (55) | 328 (38) | 8114 (54) | 576 (38) |
| Pack years: M (SD) | 6.3 (13.1) | 12 (17.1) | 6.55 (13.3) | 11.6 (16.9) |
| Major Depressive Disorder (Lifetime status) | | | | |
| Cases: N (%) | 414 (5.8) | 382 (44) | 1502 (10) | 766 (51) |
| Controls: N (%) | 6514 (92) | 373 (43) | 12704 (85) | 556 (37) |

**Table 2 | Eight CpGs associated with self-reported and/or prescription-derived antidepressant use.** The EWAS catalog was searched using the *ewascatalog* R package^45^ for other studies (n > 1000) which report a significant CpG-trait association, accessed on

17/03/2024.

| Probe | Chr | BP | Gene | $\beta$_SR_ | SE_SR_ | P_SR_ | $\beta$_PD_ | SE_PD_ | P_PD_ | Other traits associated with the CpG (EWAS catalog) ^45^ |
| --- | --- | --- | --- | --- | --- | --- | --- | --- | --- | --- |
| cg26277237 | 9 | 631910 | *KANK1* | 0.024 | 0.0033 | 9.3x10^-13^ | 0.027 | 0.0044 | 6.1x10^-10^ | C-reactive protein levels^62^ (basic model), Chronic kidney disease (basic model)^13^, Type 2 diabetes (basic model)^59^, Estimated glomerular filtration rate^69^ |
| cg04173586 | 19 | 2167496 | DOT1L | 0.023 | 0.0036 | 2.35x10^-10^ | 0.034 | 0.0048 | 7x10^-13^ | Alcohol consumption^70^, COVID-19 severity (basic model)^59^, Sex^71^, Age^72^, Alzheimer’s disease Braak stage^73^, Estimated glomerular filtration rate^69^, Schizophrenia^70^ |
| cg08907118 | 16 | 27482516 | GTF3C1 | 0.021 | 0.0039 | 6.46x10^-8^ | 0.027 | 0.0052 | 3.0x10^-7^ | C-reactive protein levels (basic model)^62^, Chronic pain (basic model)^59^, Type 2 diabetes (basic model)^59^, Sex^71^, Age^72^ |
| cg02183564 | 7 | 76874892 | CCDC146 | 0.019 | 0.0028 | 3.59x10^-11^ | 0.019 | 0.0037 | 5.17x10^-7^ | Birthweight^72^, C-reactive protein levels (basic model)^62^, Gestational age, Chronic pain (basic model)^59^, Type 2 diabetes (basic model)^59^, Age^72^ |
| cg15071067 | 2 | 74196550 | DGUOK-AS1 | 0.018 | 0.0032 | 9.84x10^-9^ | 0.020 | 0.0042 | 3.3x10^-6^ | C-reactive protein levels (basic model)^62^, Incident chronic pain (basic model)^59^, Incident type 2 diabetes (basic & fully adjusted model)^59^, Chronic pain (self-report)^59^, Type 2 diabetes (basic model)^59^ |
| cg01964004 | 2 | 74196572 | DGUOK-AS1 | 0.017 | 0.0036 | 1.3X10^-6^ | 0.026 | 0.0047 | 3.0x10^-8^ | C-reactive protein levels (basic model)^62^, Incident type 2 diabetes (basic & fully adjusted model)^59^, Chronic pain (basic model)^59^, Ischemic heart disease (basic model)^59^, Type 2 diabetes (basic model)^59^ |
| cg03222540 | 15 | 90575080 | ZNF710 | 0.014 | 0.002 | 2.05x10^-8^ | 0.014 | 0.0034 | 3.7x10^-7^ | C-reactive protein levels (basic model)^62^, Chronic kidney disease (basic model)^59^, Type 2 diabetes (basic & fully adjusted model)^59^ |
| cg04315689 | 2 | 74198896 | DGUOK-AS1 | 0.014 | 0.0026 | 5.56x10^-8^ | 0.019 | 0.0035 | 3.85x10^-8^ | C-reactive protein levels (basic model)^62^, Incident type 2 diabetes (basic model)^59^, Chronic kidney disease (basic & fully adjusted model)^59^, Chronic pain (basic model) ^59^, Type 2 diabetes (basic & fully adjusted model)^59^ |
